# Critical vulnerabilities of nutrient content claims under U.S. FDA per serving size, CAC per 100 g or mL, CAC per serving size, and CAC per 100 kcal and the nutrient content of foods based on the proposed method

**DOI:** 10.1101/2020.07.26.20162099

**Authors:** Abed Forouzesh, Fatemeh Forouzesh, Sadegh Samadi Foroushani, Abolfazl Forouzesh

**Affiliations:** Alumni Office, University of Tehran, Tehran, Iran; Iranian Research Institute of Plant Protection, Agricultural Research Education and Extension Organization (AREEO), Tehran, Iran; Department of Medicine, Tehran Medical Branch, Islamic Azad University, Tehran, Iran

**Keywords:** nutrient-poor foods, nutrient-rich foods, nutrient profiling, food labeling, dietary guidance, food analysis

## Abstract

We revealed critical vulnerabilities of nutrient content claims under FDA per serving size (serving), CAC per 100 g or mL, CAC per serving, and CAC per 100 kcal and developed the proposed method to remove all vulnerabilities. We calculated the nutrient content of foods based on nutrient content claims under the proposed method. Then, we determined nutrient content claims for foods and food groups, and specified similarities between nutrient content claims in food groups. Also, we ranked foods and food groups based on met claims of the nutrient content. Nutrient content claims based on the proposed method, including free, very low, low, source, and high claims were determined in 8596 food items, 29 nutrients, and 25 food groups of the USDA National Nutrient Database for Standard Reference, release 28. Source and high claims are used for positive nutrients (including vitamins, minerals except sodium, protein, and dietary fiber). The very low claim is used for sodium, and free and low claims are used for cholesterol, energy, saturated fat, sodium, and total fat (also known as negative nutrients). In general, critical vulnerabilities of nutrient content claims under FDA and CAC can cause: (1) excessive energy intake based on FDA and CAC per serving and CAC per 100 g or mL; (2) exceeding the DV (NRV) for low nutrients under FDA per serving and CAC per 100 g or mL by consuming small amounts of foods per day; (3) the presence of nutrient free, but not low nutrient, foods based on FDA per serving and CAC per 100 g or mL; (4) the calculation of nutrient content in inappropriate amounts of foods based on FDA per serving, CAC per 100 g or mL, CAC per 100 kcal, and CAC per serving; and (5) determination of energy from total fat and saturated fat for relevant claims in inappropriate portions based on FDA per serving and CAC per 100 g or mL. Breakfast cereals, baby foods, pork products, lamb, veal, and game products, poultry products, and beef products had the highest average of scores for source and high nutrients. Restaurant foods, fast foods, and sausages and luncheon meats had the lowest average of scores for free, very low, and low nutrients. Nutrient source or high nutrient foods for all 24 positive nutrients were found in seven food groups (American Indian/Alaska Native foods; baby foods; beverages; dairy and egg products; legumes and legume products; meals, entrees, and side dishes; vegetables and vegetable products). There were very few source and high foods for potassium and vitamin D. Also, there were very few energy free foods.

## INTRODUCTION

Nutrition is coming to the fore as a major modifiable determinant of chronic disease, with scientific evidence increasingly supporting the view that alterations in diet have strong effects, both positive and negative, on health throughout life (WHO, 2003). It has been calculated that, in 2001, chronic diseases contributed approximately 60% of the 56.5 million total reported deaths in the world and approximately 46% of the global burden of disease (WHO, 2002). The proportion of the burden of noncommunicable diseases (NCDs) is expected to increase to 57% by 2020 (WHO, 2003). It is well known that nutritional deficiency increases the risk of common infectious diseases, notably those of childhood, and vice versa (WHO, 1968).

Providing information about the nutrient content of foods cause improved nutrition (Taylor and Wilkening, 2008). Nutrient means any substance normally consumed as a constituent of food: (a) which provides energy; or (b) which is needed for growth, development and maintenance of life; or (c) a deficit of which will cause characteristic bio-chemical or physiological changes to occur (CAC, 2017). Nutrient content claim is a nutrition claim that describes the level of a nutrient contained in a food (CAC, 2007). Nutrition facts label is the most important tool which informs consumers about the nutrient content of foods. Fast and easy understanding the nutrient content of foods can be achieved by nutrient content claims on nutrition facts labels. Nutrient content claims are established to help consumers make healthier food choices, thereby reducing nutritional deficiency and the incidence of diet-related chronic diseases.

Nutrient content claims were established by several authorities, and CAC and FDA are the most important among them. The CAC is an intergovernmental body with over 170 members, within the framework of the Joint FAO/WHO Food Standards Programme established by the Food and Agriculture Organization of the United Nations (FAO) and the World Health Organization (WHO), with the purpose of protecting the health of consumers and ensuring fair practices in the food trade (CAC, 2007). The *Codex Alimentarius* (Latin, meaning Food Code) is the result of the Commission’s work: a collection of internationally adopted food standards, guidelines, codes of practice and other recommendations (CAC, 2007). Nutrient content claims based on CAC are expressed in reference amounts of 100 g or mL, serving, and 100 kcal and nutrient content claims based on FDA are expressed in a reference amount of serving. According to source and high claims under FDA and CAC per serving, the serving is derived from RACC. In addition, according to free, very low, and low claims under FDA per serving, the serving can be derived from RACC, 100 g, or 50 g. Source and high claims are used for positive nutrients (including vitamins, minerals except sodium, protein, and dietary fiber). The very low claim is used for sodium, and free and low claims are used for cholesterol, energy, saturated fat, sodium, and total fat (also known as negative nutrients). Nutrient content claims, including free, very low, low, source, and high claims were addressed in this study.

Nutrient content claims are located in the closest place to consumers, so knowing the vulnerabilities of nutrient content claims, the nutrient content of foods based on nutrient content claims, food rankings based on nutrient content claims, similarities between nutrient content claims, and all issues surrounding them play an important role in improving the knowledge of pure and applied nutrition, and thus promoting the public health. Since this article has addressed the issues raised for the first time, it can double the importance of them. The proposed method was developed by evaluating the strengths and weaknesses of reference amounts (serving, 100 g or mL, and 100 kcal) and removing all vulnerabilities of FDA per serving for nutrient content claims.

## METHODS

Information on food and nutrient profiles were provided from the USDA National Nutrient Database for Standard Reference, release 28 (SR28) (USDA ARS, 2016). The USDA National Nutrient Database for Standard Reference provides the foundation for most food composition databases in the public and private sectors (USDA ARS, 2016). Twenty-nine nutrients derived from SR28, including calcium (*n* = 8,260), cholesterol (*n* = 8,068), choline (*n* = 4,691), copper (*n* = 7,379), dietary fiber (*n* = 8,027), energy (*n* = 8,596), folate (*n* = 6,621), iron (*n* = 8,463), magnesium (*n* = 7,887), manganese (*n* = 6,489), niacin (*n* = 7,987), pantothenic acid (*n* = 6,411), phosphorus (*n* = 8,037), potassium (*n* = 8,192), protein (*n* = 8,596), riboflavin (*n* = 8,008), saturated fat (*n* = 8,252), selenium (*n* = 6,961), sodium (*n* = 8,515), thiamin (*n* = 7,990), total fat (*n* = 8,596), vitamin A (*n* = 7,110), vitamin B_6_ (*n* = 7,725), vitamin B_12_ (*n* = 7,445), vitamin C (*n* = 7,808), vitamin D (*n* = 5,435), vitamin E (*n* = 5,784), vitamin K (*n* = 5,132), and zinc (*n* = 7,911), were used in this study.

Food groups were not provided in SR28 Excel data file. However, the preparation of results on the basis of food groups requires the allocation of food groups to food items. Thus, food groups were allocated to SR28 food items using the FoodData Central website (https://fdc.nal.usda.gov). The twenty-five food groups in alphabetical order are as follows: (1) American Indian/Alaska Native foods; (2) baby foods; (3) baked products; (4) beef products; (5) beverages; (6) breakfast cereals; (7) cereal grains and pasta; (8) dairy and egg products; (9) fast foods; (10) fats and oils; (11) finfish and shellfish products; (12) fruits and fruit juices; (13) lamb, veal, and game products; (14) legumes and legume products; (15) meals, entrees, and side dishes; (16) nut and seed products; (17) pork products; (18) poultry products; (19) restaurant foods; (20) sausages and luncheon meats; (21) snacks; (22) soups, sauces, and gravies; (23) spices and herbs; (24) sweets; and (25) vegetables and vegetable products.

RACCs were not provided in SR28 Excel data file. However, the preparation of results on the basis of nutrient content claims per serving requires the allocation of RACCs to food items. Thus, RACCs were allocated to SR28 food items using the guidance prepared by the Office of Nutrition and Food Labeling. The purpose of this guidance, FDA-2018-D-1459, is to provide examples of products that belong in each of the product categories included in the tables of RACCs per Eating Occasion established in 21 CFR 101.12(b) (FDA, 2018). These values represent the amount (edible portion) of food customarily consumed per eating occasion and were primarily derived from the 1977-1978 and the 1987-1988 Nationwide Food Consumption Surveys conducted by the U.S. Department of Agriculture and updated with data from the National Health and Nutrition Examination Survey, 2003-2004, 2005-2006 and 2007-2008 conducted by the Centers for Diseases Control and Prevention, in the Department of Health and Human Services (FDA, 2018).

Meals and main dishes were not specified in SR28 Excel data file. However, the nutrient content of meals and main dishes for low and very low claims is calculated per 100 g based on FDA per serving. Thus, meals and main dishes in SR28 food items were specified using the main dish product and meal product definitions established in 21CFR101.13 (revised as of April 1, 2018).

The criteria for nutrient content claims in FDA per serving were derived from IOM (2010). DVs for nutrients used in nutrient content claims under FDA per serving and the proposed method were provided from 21 CFR101.9 (revised as of April 1, 2018) based on 2,000 kcal for adults and children aged 4 years and older (in all foods excluding baby foods) and 1,000 kcal for children 1 through 3 years of age (in baby foods). DVs for total fat were considered 77.78 g for adults and children aged 4 years and older, and 38.9 g for children 1 through 3 years of age.

The criteria for nutrient content claims in CAC per 100 g or mL, CAC per serving, and CAC per 100 kcal were derived from CAC (2013). DVs for nutrients used in nutrient content claims under CAC per 100 g or mL, CAC per serving, and CAC per 100 kcal were provided from CAC (2017) and 21 CFR101.9 (revised as of April 1, 2018) based on 2,000 kcal for adults and children aged 4 years and older (in all foods excluding baby foods) and 1,000 kcal for children 1 through 3 years of age (in baby foods), respectively. The DV for total fat was not provided for adults and children aged 4 years and older in CAC (2017), so it was considered 66.667 g.

Establishing nutrient amounts of free, very low, and low claims based on the proposed method requires determining the number of daily servings. The number of daily servings at three energy levels (1,600 kcal, 2,200 kcal, and 2,800 kcal) was 15-26 servings: fruits, 2-4 servings; vegetables, 3-5 servings; dairy, 2-3 servings; grains, 6-11 servings; and protein foods, 5-7 ounces to provide a total of 2-3 servings (Bowman *et al*., 1998). According to the number of daily servings at three energy levels, the number of daily servings at the 2,000 kcal level is determined 17.8-20 servings. Since exceeding the number of daily servings may result in excess of the DV for foods low in cholesterol, energy, saturated fat, sodium, and total fat, the number of daily servings was determined 20 in this study. Since a typical consumer eats 20 or fewer servings of food per day (HHS, 1991; Kessler *et al*., 2003), the definition of “low” should enable that consumer to stay at or below 100 percent of the DV for a given nutrient (Kessler *et al*., 2003).

Nutrient content claims based on the proposed method, FDA per serving, CAC per 100 g or mL, CAC per serving, and CAC per 100 kcal are given in Tables 1 and 2.

**Table 1:**
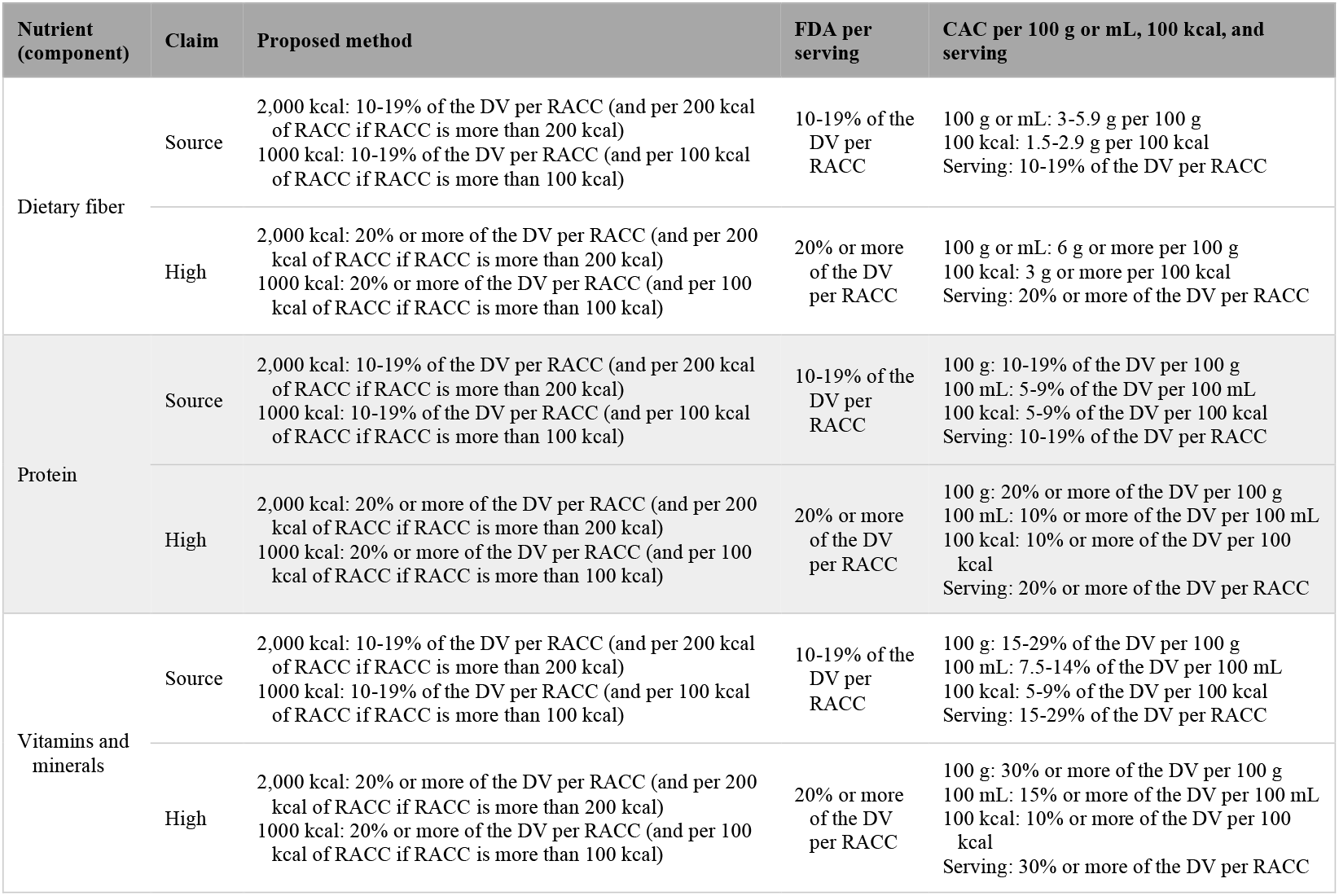
Source and high claims based on the proposed method, FDA per serving, CAC per 100 g or mL, CAC per 100 kcal, and CAC per serving for dietary fiber, protein, vitamins, and minerals.

| Nutrient (component) | Claim | Proposed method | FDA per serving | CAC per 100 g or mL, 100 kcal, and serving |
| --- | --- | --- | --- | --- |
| Dietary fiber | Source | 2,000 kcal: 10-19% of the DV per RACC (and per 200 kcal of RACC if RACC is more than 200 kcal)<br>1000 kcal: 10-19% of the DV per RACC (and per 100 kcal of RACC if RACC is more than 100 kcal) | 10-19% of the DV per RACC | 100 g or mL: 3-5.9 g per 100 g<br>100 kcal: 1.5-2.9 g per 100 kcal<br>Serving: 10-19% of the DV per RACC |
|  | High | 2,000 kcal: 20% or more of the DV per RACC (and per 200 kcal of RACC if RACC is more than 200 kcal)<br>1000 kcal: 20% or more of the DV per RACC (and per 100 kcal of RACC if RACC is more than 100 kcal) | 20% or more of the DV per RACC | 100 g or mL: 6 g or more per 100 g<br>100 kcal: 3 g or more per 100 kcal<br>Serving: 20% or more of the DV per RACC |
| Protein | Source | 2,000 kcal: 10-19% of the DV per RACC (and per 200 kcal of RACC if RACC is more than 200 kcal)<br>1000 kcal: 10-19% of the DV per RACC (and per 100 kcal of RACC if RACC is more than 100 kcal) | 10-19% of the DV per RACC | 100 g: 10-19% of the DV per 100 g<br>100 mL: 5-9% of the DV per 100 mL<br>100 kcal: 5-9% of the DV per 100 kcal<br>Serving: 10-19% of the DV per RACC |
|  | High | 2,000 kcal: 20% or more of the DV per RACC (and per 200 kcal of RACC if RACC is more than 200 kcal)<br>1000 kcal: 20% or more of the DV per RACC (and per 100 kcal of RACC if RACC is more than 100 kcal) | 20% or more of the DV per RACC | 100 g: 20% or more of the DV per 100 g<br>100 mL: 10% or more of the DV per 100 mL<br>100 kcal: 10% or more of the DV per 100 kcal<br>Serving: 20% or more of the DV per RACC |
| Vitamins and minerals | Source | 2,000 kcal: 10-19% of the DV per RACC (and per 200 kcal of RACC if RACC is more than 200 kcal)<br>1000 kcal: 10-19% of the DV per RACC (and per 100 kcal of RACC if RACC is more than 100 kcal) | 10-19% of the DV per RACC | 100 g: 15-29% of the DV per 100 g<br>100 mL: 7.5-14% of the DV per 100 mL<br>100 kcal: 5-9% of the DV per 100 kcal<br>Serving: 15-29% of the DV per RACC |
|  | High | 2,000 kcal: 20% or more of the DV per RACC (and per 200 kcal of RACC if RACC is more than 200 kcal)<br>1000 kcal: 20% or more of the DV per RACC (and per 100 kcal of RACC if RACC is more than 100 kcal) | 20% or more of the DV per RACC | 100 g: 30% or more of the DV per 100 g<br>100 mL: 15% or more of the DV per 100 mL<br>100 kcal: 10% or more of the DV per 100 kcal<br>Serving: 30% or more of the DV per RACC |

**Table 2:** Free, very low, and low claims based on the proposed method, FDA per serving, and CAC per 100 g or mL for cholesterol, energy, saturated fat, sodium, and total fat.

| Nutrient (component) | Claim | Proposed method | FDA per serving | CAC per 100 g or mL |
| --- | --- | --- | --- | --- |
| Cholesterol | Free | 2,000 kcal: 1.5 mg (0.5% of the DV) or less cholesterol and 1 g (5% of the DV) or less saturated fat per RACC (and per 30 g of food if RACC is less than 30 g)<br>1000 kcal: 1.5 mg (0.5% of the DV) or less cholesterol and 0.5 g (5% of the DV) or less saturated fat per RACC (and per 30 g of food if RACC is less than 30 g) | All foods excluding meals and main dishes: Less than 2 mg (0.667% of the DV) cholesterol and 2 g (10% of the DV) or less saturated fat per RACC and per labeled serving<br>Meals and main dishes: Less than 2 mg (0.667% of the DV) cholesterol and 2 g (10% of the DV) or less saturated fat per labeled serving | Solids: 5 mg (1.667% of the DV) or less cholesterol and 1.5 g (7.5% of the DV) or less saturated fat per 100 g and not more than 10% of energy from saturated fat<br>Liquids: 5 mg (1.667% of the DV) or less cholesterol and 0.75 g (3.75% of the DV) or less saturated fat per 100 mL and not more than 10% of energy from saturated fat |
|  | Low | 2,000 kcal: 15 mg (5% of the DV) or less cholesterol and 1 g (5% of the DV) or less saturated fat per RACC (and per 30 g of food if RACC is less than 30 g)<br>1000 kcal: 15 mg (5% of the DV) or less cholesterol and 0.5 g (5% of the DV) or less saturated fat per RACC (and per 30 g of food if RACC is less than 30 g) | All foods excluding meals and main dishes: 20 mg (6.67% of the DV) or less cholesterol (and per 50 g of food if RACC is 30 g or less or 2 tablespoons or less) and 2 g (10% of the DV) or less saturated fat per RACC<br>Meals and main dishes: 20 mg (6.67% of the DV) or less cholesterol and 2 g (10% of the DV) or less saturated fat per 100 g | Solids: 20 mg (6.67% of the DV) or less cholesterol and 1.5 g (7.5% of the DV) or less saturated fat per 100 g and not more than 10% of energy from saturated fat<br>Liquids: 10 mg (3.333% of the DV) or less cholesterol and 0.75 g (3.75% of the DV) or less saturated fat per 100 mL and not more than 10% of energy from saturated fat |
| Energy | Free | 2,000 kcal: 10 kcal (0.5% of the DV) or less energy per RACC (and per 30 g of food if RACC is less than 30 g)<br>1000 kcal: 5 kcal (0.5% of the DV) or less energy per RACC (and per 30 g of food if RACC is less than 30 g) | Less than 5 kcal (0.25% of the DV) energy per RACC and per labeled serving | Solids and liquids: 4 kcal (0.2% of the DV) or less energy per 100 g (solids) or 100 mL (liquids) |
|  | Low | 2,000 kcal: 100 kcal (5% of the DV) or less energy per RACC (and per 30 g of food if RACC is less than 30 g)<br>1000 kcal: 50 kcal (5% of the DV) or less energy per RACC (and per 30 g of food if RACC is less than 30 g) | All foods excluding meals and main dishes: 40 kcal (2% of the DV) energy or less per RACC (and per 50 g of food if RACC is 30 g or less or 2 tablespoons or less)<br>Meals and main dishes: 120 kcal (6% of the DV) or less energy per 100 g | Solids: 40 kcal (2% of the DV) or less energy per 100 g<br>Liquids: 20 kcal (1% of the DV) or less energy per 100 mL |
| Saturated fat | Free | 2,000 kcal: 0.1 g (0.5% of the DV) or less saturated fat per RACC (and per 30 g of food if RACC is less than 30 g) and 9% or less of energy from saturated fat<br>1000 kcal: 0.05 g (0.5% of the DV) or less saturated fat per RACC (and per 30 g of food if RACC is less than 30 g) and 9% or less of energy from saturated fat | All foods excluding meals and main dishes: Less than 0.5 g (2.5% of the DV) saturated fat and less than 0.5 g *trans* fat per RACC and per labeled serving<br>Meals and main dishes: Less than 0.5 g (2.5% of the DV) saturated fat and less than 0.5 g *trans* fat per labeled serving | Solids and liquids: 0.1 g (0.5% of the DV) or less saturated fat per 100 g (solids) or 100 mL (liquids) |
|  | Low | 2,000 kcal: 1 g (5% of the DV) or less saturated fat per RACC (and per 30 g of food if RACC is less than 30 g) and 9% or less of energy from saturated fat<br>1000 kcal: 0.5 g (5% of the DV) or less saturated fat per RACC (and per 30 g of food if RACC is less than 30 g) and 9% or less of energy from saturated fat | All foods excluding meals and main dishes: 1 g (5% of the DV) or less saturated fat per RACC and not more than 15% of energy from saturated fat<br>Meals and main dishes: 1 g (5% of the DV) or less saturated fat per 100 g and less than 10% of energy from saturated fat | Solids: 1.5 g (7.5% of the DV) or less saturated fat per 100 g and not more than 10% of energy from saturated fat<br>Liquids: 0.75 g (3.75% of the DV) or less saturated fat per 100 mL and not more than 10% of energy from saturated fat |
| Sodium | Free | 2,000 kcal: 11.5 mg (0.5% of the DV) or less sodium per RACC (and per 30 g of food if RACC is less than 30 g)<br>1000 kcal: 7.5 mg (0.5% of the DV) or less sodium per RACC (and per 30 g of food if RACC is less than 30 g) | All foods excluding meals and main dishes: Less than 5 mg (0.218% of the DV) sodium per RACC and per labeled serving<br>Meals and main dishes: Less than 5 mg (0.218% of the DV) sodium per labeled serving | Solids and liquids: 5 mg (0.25% of the DV) or less sodium per 100 g |
|  | Very low | 2,000 kcal: 57.5 mg (2.5% of the DV) or less sodium per RACC (and per 30 g of food if RACC is less than 30 g)<br>1000 kcal: 37.5 mg (2.5% of the DV) or less sodium per RACC (and per 30 g of food if RACC is less than 30 g) | All foods excluding meals and main dishes: 35 mg (1.522% of the DV) or less sodium per RACC (and per 50 g of food if RACC is 30 g or less or 2 tablespoons or less)<br>Meals and main dishes: 35 mg (1.522% of the DV) or less sodium per 100 g | Solids and liquids: 40 mg (2% of the DV) or less sodium per 100 g |
|  | Low | 2,000 kcal: 115 mg (5% of the DV) or less sodium per RACC (and per 30 g of food if RACC is less than 30 g)<br>1000 kcal: 75 mg (5% of the DV) or less sodium per RACC (and per 30 g of food if RACC is less than 30 g) | All foods excluding meals and main dishes: 140 mg (6.09% of the DV) or less sodium per RACC (and per 50 g of food if RACC is 30 g or less or 2 tablespoons or less)<br>Meals and main dishes: 140 mg (6.09% of the DV) or less sodium per 100 g | Solids and liquids: 120 mg (6% of the DV) or less sodium per 100 g |
| Total fat | Free | 2,000 kcal: 0.389 g (0.5% of the DV) or less total fat per RACC (and per 30 g of food if RACC is less than 30 g) and 35% or less of energy from total fat<br>1000 kcal: 0.195 g (0.5% of the DV) or less total fat per RACC (and per 30 g of food if RACC is less than 30 g) and 35% or less of energy from total fat | All foods excluding meals and main dishes: Less than 0.5 g (0.643% of the DV) total fat per RACC and per labeled serving<br>Meals and main dishes: Less than 0.5 g (0.643% of the DV) total fat per labeled serving | Solids and liquids: 0.5 g (0.75% of the DV) or less total fat per 100 g (solids) or 100 mL (liquids) |
|  | Low | 2,000 kcal: 3.889 g (5% of the DV) or less total fat per RACC (and per 30 g of food if RACC is less than 30 g) and 35% or less of energy from total fat<br>1000 kcal: 1.945 g (5% of the DV) or less total fat per RACC (and per 30 g of food if RACC is less than 30 g) and 35% or less of energy from total fat | All foods excluding meals and main dishes: 3 g (3.857% of the DV) or less total fat per RACC (and per 50 g of food if RACC is 30 g or less or 2 tablespoons or less)<br>Meals and main dishes: 3 g (3.857% of the DV) or less total fat per 100 g and not more than 30% of energy from total fat | Solids: 3 g (4.5% of the DV) or less total fat per 100 g<br>Liquids: 1.5 g (2.25% of the DV) or less total fat per 100 mL |

According to the proposed method, if energy per RACC is 200 kcal or less, source and high claims for adults and children aged 4 years and older are defined as 10-19% and 20% or more of the DV per RACC, respectively. Also, if energy per RACC is 100 kcal or less, source and high claims for children 1 through 3 years of age are defined as 10-19% and 20% or more of the DV per RACC, respectively. Energy per RACC for solid and liquid foods is calculated by formulas 1 and 2, respectively.

**Formula 1:** Energy _(kcal)_ per RACC (for solids) = (RACC _(g)_ ÷ 100) × energy _(kcal/100 g)_

**Formula 2:** Energy _(kcal)_ per RACC (for liquids) = (RACC _(mL)_ ÷ 100) × (density _(g/mL)_ × energy _(kcal/100 g)_)

If one food meets definitions of source claims for vitamins, minerals, protein, or dietary fiber by 10% of the DV per RACC, 10 eating occasions per day of that food meet the DV for vitamins, minerals, protein, or dietary fiber. If energy per RACC of one food is more than 200 kcal, 10 eating occasions per day of that food result in excess of the DV for energy in adults and children aged 4 years and older. Also, if energy per RACC of one baby food is more than 100 kcal, 10 eating occasions per day of that baby food result in excess of the DV for energy in children 1 through 3 years of age.

If energy per RACC is more than 200 kcal, source and high claims for adults and children aged 4 years and older are defined as 10-19% and 20% or more of the DV per 200 kcal of RACC, respectively. If energy per RACC is more than 200 kcal, 200 kcal of RACC for solid and liquid foods is calculated by formulas 3 and 4, respectively.

**Formula 3:** 200 kcal of RACC _(g)_ (for solids) = [(200 × RACC _(g)_)] ÷ [(RACC _(g)_ ÷ 100) × energy _(kcal/100 g)_]

**Formula 4:** 200 kcal of RACC _(mL)_ (for liquids) = [(200 × RACC _(mL)_] ÷ [(RACC _(mL)_ ÷ 100) × (density _(g/mL)_ × energy _(kcal/100 g)_)]

If energy per RACC is more than 100 kcal, source and high claims for children 1 through 3 years of age are defined as 10-19% and 20% or more of the DV per 100 kcal of RACC, respectively. If energy per RACC is more than 100 kcal, 100 kcal of RACC for solid and liquid baby foods is calculated by formulas 5 and 6, respectively.

**Formula 5:** 100 kcal of RACC _(g)_ (for solids) = [(100 × RACC _(g)_)] ÷ [(RACC _(g)_ ÷ 100) × energy _(kcal/100 g)_]

**Formula 6:** 100 kcal of RACC _(mL)_ (for liquids) = [(100 × RACC _(mL)_] ÷ [(RACC _(mL)_ ÷ 100) × (density _(g/mL)_ × energy _(kcal/100 g)_)]

For example, if a chocolate drink (milk and soy based, ready to drink) contains RACC of 240 mL, 101 kcal of energy per 100 g, and density of 1.00174 g/mL, calculate the serving of this chocolate drink for source and high claims under the proposed method. Energy per RACC of this chocolate drink is calculated by formula 2 as follows.

Energy per RACC: (240 ÷ 100) × (1.00174 × 101) = 242.822 _(kcal)_

Since energy per RACC is more than 200 kcal, the serving of this chocolate drink for source and high claims (200 kcal of RACC) is calculated by formula 4 as follows.

200 kcal of RACC: [200 × 240] ÷ [(240 ÷ 100) × (1.00174 × 101)] = 197.675 _(mL)_

Exceeding the DV for cholesterol, energy, saturated fat, sodium, and total fat may cause health problems. Thus, criteria for free, very low, and low claims were designed to minimize excess of the DV for these nutrients. Some points on free, very low, and low claims under the proposed method include: (1) energy does not play a role in determining the nutrient content and amount of foods for these claims; (2) the nutrient content of foods for these claims is calculated per RACC, but if RACC is small (small RACC means RACC of less than 30 g), it is calculated per 30 g (the 30 g criterion refers to the prepared form of the food); (3) nutrient amounts of free, very low, and low claims are defined as 0.5% or less, 2.5% or less, and 5% or less of the DV per serving, respectively; (4) on the basis of the portion of saturated fat in energy, saturated fat free and low saturated fat claims are defined as 9% or less of energy from saturated fat; (5) on the basis of the portion of total fat in energy, total fat free and low total fat claims are defined as 35% or less of energy from total fat; and (6) since saturated fat can raise blood cholesterol levels, the saturated fat content for cholesterol free and low cholesterol claims is defined as 5% or less of the DV per serving.

The score of source and high nutrients for food items was calculated by formula 7.

**Formula 7:** Score of source and high nutrients _(%)_ = [((100) ÷ (a × 2)) × (b)] + [((100) ÷ (a × 2)) × (c × 2)] a = number of high claims; b = number of source nutrients; c = number of high nutrients.

The score of free, very low, and low nutrients for food items was calculated by formula 8.

**Formula 8:** Score of free, very low, and low nutrients _(%)_ = [((100) ÷ (d × 2)) × (e × 2)] + [((100) ÷ (d × 2)) × (f × 1.5)] + [((100) ÷ (d × 2)) × (g)]

d = number of free claims; e = number of free nutrients; f = number of very low nutrients that are not free; g = number of low nutrients that are not free or very low.

The score of free, very low, low, source, and high nutrients for food items was calculated by formula 9.

**Formula 9:** Score of free, very low, low, source, and high nutrients _(%)_ = [((100) ÷ (i × 2)) × (j × 2)] + [((100) ÷ (i × 2)) × (f × 1.5)] + [((100) ÷ (i × 2)) × (b + g)]

b = number of source nutrients; f = number of very low nutrients that are not free; g = number of low nutrients that are not free or very low; i = number of free and high claims; j = number of free and high nutrients.

For example, the score of free, very low, low, source, and high nutrients under the proposed method for boiled spinach (NDB number 11458; number of source nutrients = 7; number of very low nutrients that are not free = 0; number of low nutrients that are not free or very low = 2; number of free and high claims = 29; number of free and high nutrients = 11) is calculated by formula 9 as follows.

Score of free, very low, low, source, and high nutrients: [((100) ÷ (29 × 2)) × (11 × 2)] + [((100) ÷ (29 × 2)) × (0 × 1.5)] + [((100) ÷ (29 × 2)) × (7 + 2)] = 53.45%

Percentages in this study were classified as excellent (90 ≤ percentage ≤ 100), almost excellent (80 ≤ percentage < 90), very good (70 ≤ percentage < 80), almost very good (60 ≤ percentage < 70), good (50 ≤ percentage < 60), almost good (40 ≤ percentage < 50), satisfactory (30 ≤ percentage < 40), acceptable (20 ≤ percentage < 30), almost insufficient (10 ≤ percentage < 20), insufficient (0 < percentage < 10), and absent (percentage = 0).

## RESULTS AND DISCUSSION

### Excessive energy intake based on source claims for vitamins, minerals, protein and dietary fiber under FDA per serving and source claims for protein and dietary fiber under CAC per serving

Source claims for vitamins, minerals, protein, and dietary fiber under FDA per serving and source claims for protein and dietary fiber under CAC per serving have caused excessive energy intake from some foods. If one food meets definitions of source claims for vitamins, minerals, protein, or dietary fiber by 10% of the DV per RACC, 10 eating occasions per day of that food meet the DV for vitamins, minerals, protein, or dietary fiber. If energy of one food per RACC is more than 200 kcal, 10 eating occasions per day of that food result in excess of the DV for energy in adults and children aged 4 years and older. Also, if energy of one baby food per RACC is more than 100 kcal, 10 eating occasions per day of that baby food result in excess of the DV for energy in children 1 through 3 years of age.

Exceeding the DV for energy based on source claims for vitamins, minerals, protein, and dietary fiber under FDA per serving and source claims for protein and dietary fiber under CAC per serving by consuming 10 RACCs in food groups was as follows: it was almost excellent to excellent in three food groups (meals, entrees, and side dishes; restaurant foods; fast foods); it was acceptable to satisfactory in seven food groups (baked products; poultry products; baby foods; pork products; breakfast cereals; beef products; lamb, veal, and game products); it was insufficient to almost insufficient in 13 food groups (vegetables and vegetable products; fruits and fruit juices; snacks; sweets; soups, sauces, and gravies; dairy and egg products; legumes and legume products; finfish and shellfish products; beverages; sausages and luncheon meats; nut and seed products; cereal grains and pasta; American Indian/Alaska Native foods); and it was absent in two food groups (fats and oils; spices and herbs) (Figure 1).

**Figure 1:**
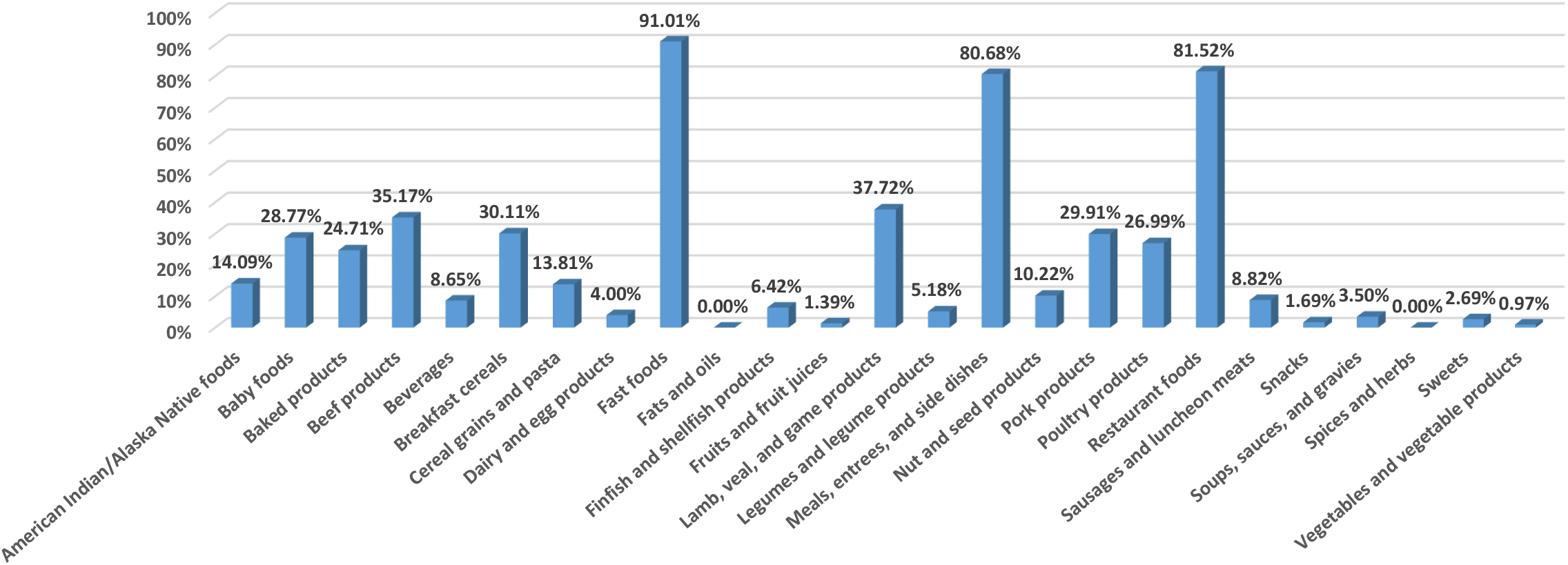
Exceeding the daily value for energy based on source claims for vitamins, minerals, protein, and dietary fiber under FDA per serving and source claims for protein and dietary fiber under CAC per serving by consuming 10 RACCs in food groups. All foods except the baby foods are based on the reference energy intake of 2,000 kcal for adults and children aged 4 years and older. The baby foods are based on the reference energy intake of 1,000 kcal for children 1 through 3 years of age.

### Excessive energy intake based on source claims for vitamins and minerals under CAC per serving

Source claims for vitamins and minerals under CAC per serving have caused excessive energy intake from some foods. If one food meets definitions of source claims for vitamins or minerals by 15% of the DV per RACC, 6.6667 eating occasions per day of that food meet the DV for vitamins or minerals. If energy of one food per RACC is more than 300 kcal, 6.6667 eating occasions per day of that food result in excess of the DV for energy in adults and children aged 4 years and older. Also, if energy of one baby food per RACC is more than 150 kcal, 6.6667 eating occasions per day of that baby food result in excess of the DV for energy in children 1 through 3 years of age.

Exceeding the DV for energy based on source claims for vitamins and minerals under CAC per serving by consuming 6.6667 RACCs in food groups was as follows: it was very good in one food group (fast foods); it was good in one food group (restaurant foods); it was satisfactory in one food group (meals, entrees, and side dishes); it was insufficient to almost insufficient in 14 food groups (vegetables and vegetable products; dairy and egg products; legumes and legume products; fruits and fruit juices; sweets; beverages; baby foods; nut and seed products; American Indian/Alaska Native foods; beef products; poultry products; pork products; lamb, veal, and game products; baked products); and it was absent in eight food groups (breakfast cereals; cereal grains and pasta; fats and oils; finfish and shellfish products; sausages and luncheon meats; snacks; soups, sauces, and gravies; spices and herbs) (Figure 2).

**Figure 2:**
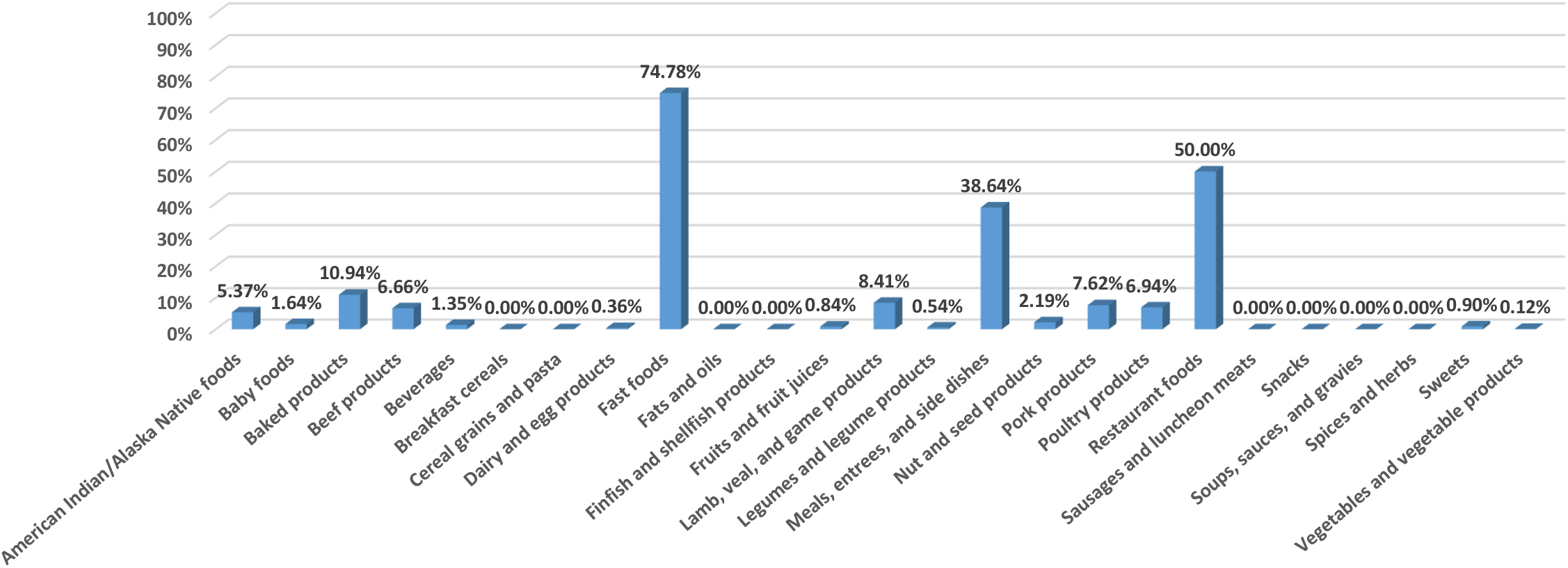
Exceeding the daily value for energy based on source claims for vitamins and minerals under CAC per serving by consuming 6.6667 RACCs in food groups. All foods except the baby foods are based on the reference energy intake of 2,000 kcal for adults and children aged 4 years and older. The baby foods are based on the reference energy intake of 1,000 kcal for children 1 through 3 years of age.

### Excessive energy intake based on the protein source claim under CAC per 100 g or mL

The protein source claim under CAC per 100 g or mL has caused excessive energy intake from some foods. If one solid food meets the definition of protein source claim by 10% of the DV per 100 g or if one liquid food meets the definition of protein source claim by 5% of the DV per 100 mL, 10 eating occasions per day of that solid food or 20 eating occasions per day of that liquid food meet the DV for protein. If energy of one solid food per 100 g is more than 200 kcal or if energy of one liquid food per 100 mL is more than 100 kcal, 10 eating occasions per day of that solid food or 20 eating occasions per day of that liquid food result in excess of the DV for energy in adults and children aged 4 years and older. Also, if energy of one solid baby food per 100 g is more than 100 kcal or if energy of one liquid baby food per 100 mL is more than 50 kcal, 10 eating occasions per day of that solid baby food or 20 eating occasions per day of that liquid baby food result in excess of the DV for energy in children 1 through 3 years of age.

Exceeding the DV for energy based on the protein source claim under CAC per 100 g or mL by consuming 1000 g of solid food or 2,000 mL of liquid food in food groups was as follows: it was almost excellent to excellent in six food groups (fast foods; fats and oils; breakfast cereals; nut and seed products; baked products; snacks); it was almost very good to very good in four food groups (cereal grains and pasta; sausages and luncheon meats; spices and herbs; sweets); it was almost good to good in six food groups (meals, entrees, and side dishes; baby foods; lamb, veal, and game products; beef products; dairy and egg products; restaurant foods); it was acceptable to satisfactory in five food groups (beverages; legumes and legume products; American Indian/Alaska Native foods; poultry products; pork products); and it was insufficient to almost insufficient in four food groups (vegetables and vegetable products; soups, sauces, and gravies; fruits and fruit juices; finfish and shellfish products) (Figure 3).

**Figure 3:**
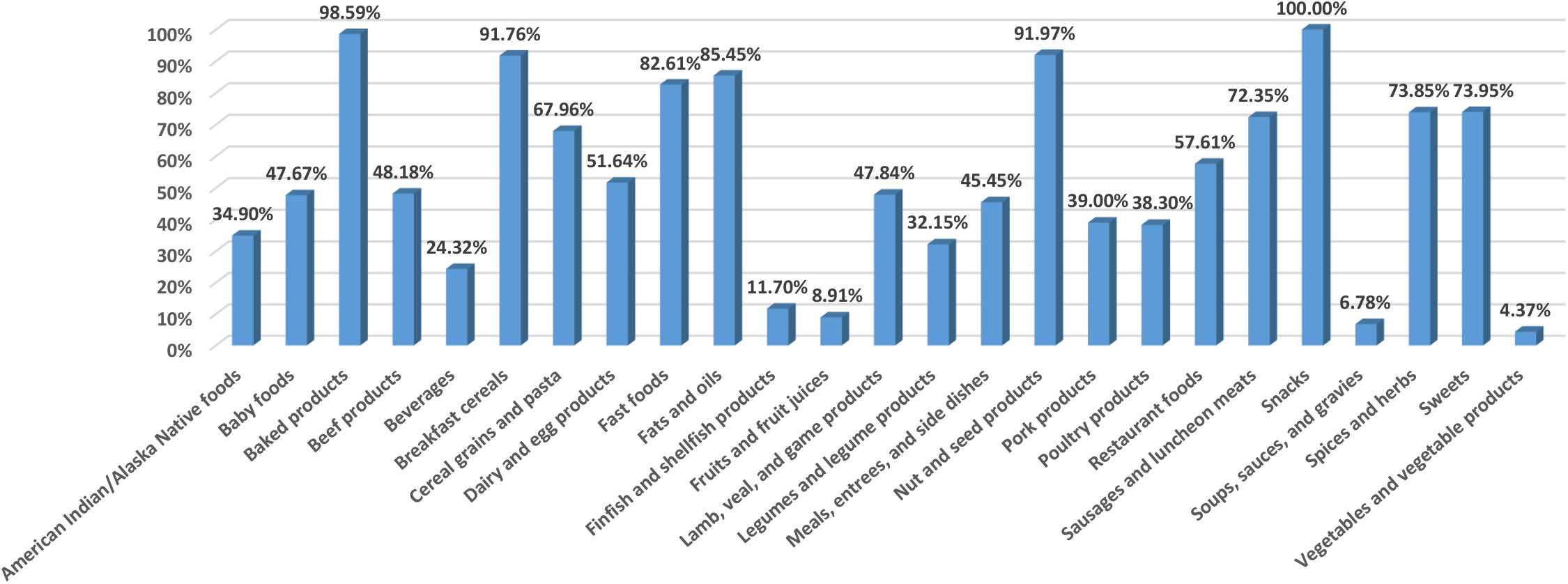
Exceeding the daily value for energy based on the protein source claim under CAC per 100 g or mL by consuming 1000 g of solid food or 2,000 mL of liquid food in food groups. All foods except the baby foods are based on the reference energy intake of 2,000 kcal for adults and children aged 4 years and older. The baby foods are based on the reference energy intake of 1,000 kcal for children 1 through 3 years of age.

### Excessive energy intake based on source claims for vitamins and minerals under CAC per 100 g or mL

Source claims for vitamins and minerals under CAC per 100 g or mL have caused excessive energy intake from some foods. If one solid food meets definitions of source claims for vitamins or minerals by 15% of the DV per 100 g or if one liquid food meets definitions of source claims for vitamins or minerals by 7.5% of the DV per 100 mL, 6.6667 eating occasions per day of that solid food or 13.333 eating occasions per day of that liquid food meet the DV for vitamins or minelals. If energy of one solid food per 100 g is more than 300 kcal or if energy of one liquid food per 100 mL is more than 150 kcal, 6.6667 eating occasions per day of that solid food or 13.333 eating occasions per day of that liquid food result in excess of the DV for energy in adults and children aged 4 years and older. Also, if energy of one solid baby food per 100 g is more than 150 kcal or if energy of one liquid baby food per 100 mL is more than 75 kcal, 6.6667 eating occasions per day of that solid baby food or 13.333 eating occasions per day of that liquid baby food result in excess of the DV for energy in children 1 through 3 years of age.

Exceeding the DV for energy based on source claims for vitamins and minerals under CAC per 100 g or mL by consuming 666.67 g of solid food or 1333.3 mL of liquid food in food groups was as follows: it was almost excellent to excellent in four food groups (fats and oils; nut and seed products; breakfast cereals; snacks); it was almost very good to very good in three food groups (cereal grains and pasta; sweets; baked products); it was almost good in one food group (spices and herbs); it was acceptable to satisfactory in seven food groups (fast foods; legumes and legume products; baby foods; restaurant foods; American Indian/Alaska Native foods; dairy and egg products; sausages and luncheon meats); and it was insufficient to almost insufficient in ten food groups (finfish and shellfish products; vegetables and vegetable products; fruits and fruit juices; soups, sauces, and gravies; poultry products; beef products; pork products; lamb, veal, and game products; meals, entrees, and side dishes; beverages) (Figure 4).

**Figure 4:**
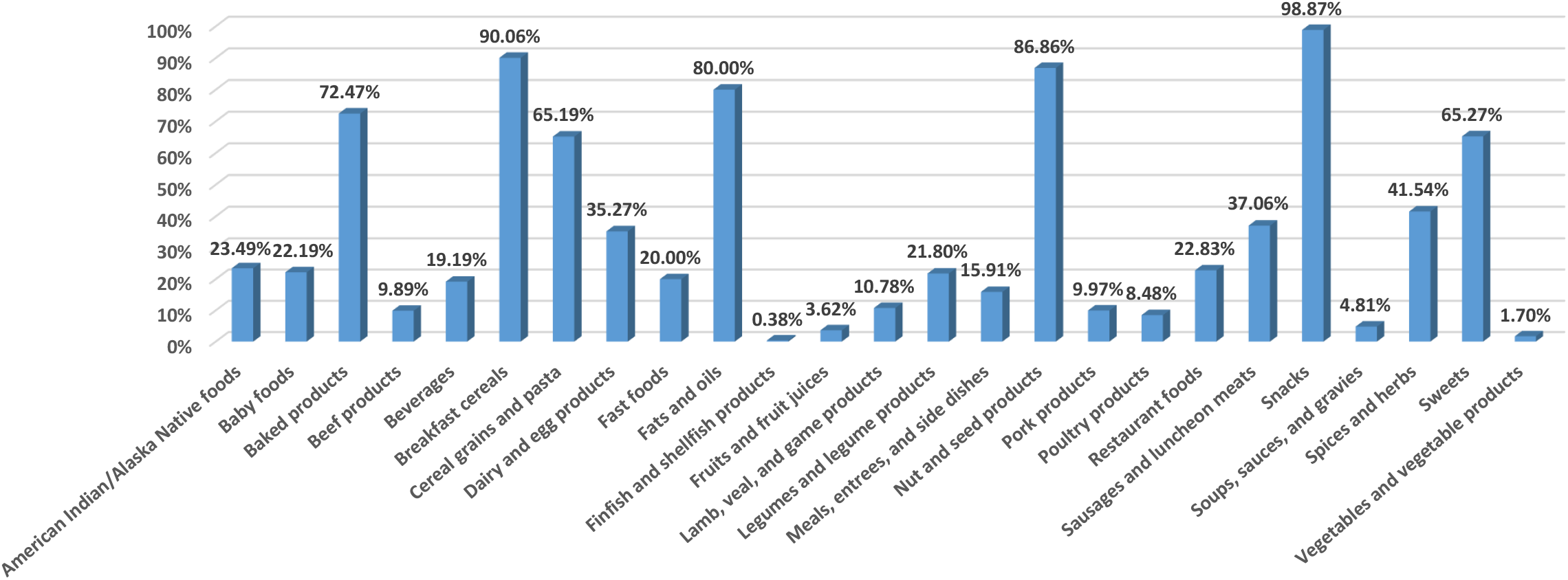
Exceeding the daily value for energy based on source claims for vitamins and minerals under CAC per 100 g or mL by consuming 666.67 g of solid food or 1333.3 mL of liquid food in food groups. All foods except the baby foods are based on the reference energy intake of 2,000 kcal for adults and children aged 4 years and older. The baby foods are based on the reference energy intake of 1,000 kcal for children 1 through 3 years of age.

### Exceeding the DV for low nutrients based on FDA per serving by consuming small amounts of foods per day

Exceeding the DV for saturated fat in low cholesterol foods under FDA per serving was started by consuming 4.45 RACCs in one food from the group of meals, entrees, and side dishes (NDB number 22952) (Table 3). Exceeding the DV for saturated fat in this low cholesterol food was due to the high saturated fat amount of the low cholesterol claim in meals and main dishes.

**Table 3:** Exceeding the daily value for saturated fat in one low cholesterol food under FDA per serving by consuming 4.45 RACCs.

| NDB No | Short description | Food group | RACC | Unit | Meal or main dish | Saturated fat (g/100 g) | Cholesterol (mg/100 g) | Low cholesterol | Saturated fat (g/4.45 RACCs) |
| --- | --- | --- | --- | --- | --- | --- | --- | --- | --- |
| 22952 | SWANSON,CHICK & DUMPLINGS | Meals, Entrees, and Side Dishes | 247 | g | Yes | 1.822 | 14 | Yes | 20.027 |

Exceeding the DV for cholesterol in low cholesterol foods under FDA per serving was started by consuming 6.9 RACCs in one food from the group of legumes and legume products (NDB number 16059) (Table 4). Exceeding the DV for cholesterol in this low cholesterol food was due to the high cholesterol amount of the low cholesterol claim in meals and main dishes.

**Table 4:** Exceeding the daily value for cholesterol in one low cholesterol food under FDA per serving by consuming 6.9 RACCs.

| NDB No | Short description | Food group | RACC | Unit | Meal or main dish | Saturated fat (g/100 g) | Cholesterol (mg/100 g) | Low cholesterol | Cholesterol (mg/6.9 RACCs) |
| --- | --- | --- | --- | --- | --- | --- | --- | --- | --- |
| 16059 | CHILI WITH BEANS,CANNED | Legumes and Legume Products | 256 | g | Yes | 1.133 | 17 | Yes | 300.288 |

Exceeding the DV for energy in low energy foods under FDA per serving was started by consuming 7.07 RACCs in one food from the group of meals, entrees, and side dishes (NDB number 22911) (Table 5). Exceeding the DV for energy in this low energy food was due to the high energy amount of the low energy claim in meals and main dishes.

**Table 5:** Exceeding the daily value for energy in one low energy food under FDA per serving by consuming 7.07 RACCs.

| NDB No | Short description | Food group | RACC | Unit | Meal or main dish | Energy (kcal/100 g) | Low energy | Energy (kcal/7.07 RACCs) |
| --- | --- | --- | --- | --- | --- | --- | --- | --- |
| 22911 | CHILI,NO BNS,CND ENTREE | Meals, Entrees, and Side Dishes | 240 | g | Yes | 118 | Yes | 2002.224 |

Exceeding the DV for saturated fat in low saturated fat foods under FDA per serving was started by consuming 8.01 RACCs in two foods from the group of meals, entrees, and side dishes (NDB numbers 22940 and 22942) (Table 6). Exceeding the DV for saturated fat in these two low saturated fat foods was due to the high saturated fat amount of the low saturated fat claim in meals and main dishes.

**Table 6:** Exceeding the daily value for saturated fat in two low saturated fat foods under FDA per serving by consuming 8.01 RACCs.

| NDB No | Short description | Food group | RACC | Unit | Meal or main dish | Energy (kcal/100 g) | Saturated fat (g/100 g) | Energy from saturated fat (%) | Low saturated fat | Saturated fat (g/8.01 RACCs) |
| --- | --- | --- | --- | --- | --- | --- | --- | --- | --- | --- |
| 22940 | SPAGHETTIOS,SPAGHETTIOS W/ MEATBALLS | Meals, Entrees, and Side Dishes | 252 | g | Yes | 95 | 0.992 | 9.398% | Yes | 20.024 |
| 22942 | CAMPBELL'S,SPAGHETTIOS,SPAGHETTIOS A TO Z'S W/ MEATBALLS | Meals, Entrees, and Side Dishes | 252 | g | Yes | 95 | 0.992 | 9.398% | Yes | 20.024 |

Exceeding the DV for total fat in low total fat foods under FDA per serving was started by consuming 11.11 RACCs in two foods from the group of meals, entrees, and side dishes (NDB numbers 22940 and 22942) (Table 7). Exceeding the DV for total fat in these two low total fat foods was due to the high total fat amount of the low total fat claim in meals and main dishes.

**Table 7:**
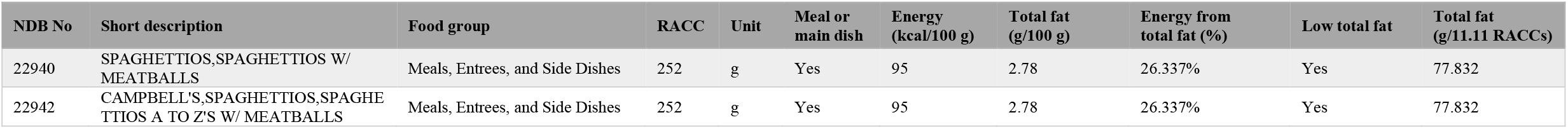
Exceeding the daily value for total fat in two low total fat foods under FDA per serving by consuming 11.11 RACCs.

Exceeding the DV for sodium in low sodium foods under FDA per serving was started by consuming 12.08 RACCs in one food from the group of baby foods (NDB number 3048) (Table 8). Exceeding the DV for sodium in this low sodium food was due to the high sodium amount of the low sodium claim in meals and main dishes.

**Table 8:** Exceeding the daily value for sodium in one low sodium food under FDA per serving by consuming 12.08 RACCs.

| NDB No | Short description | Food group | RACC | Unit | Meal or main dish | Sodium (mg/100 g) | Low sodium | Sodium (mg/12.08 RACCs) |
| --- | --- | --- | --- | --- | --- | --- | --- | --- |
| 3048 | BABYFOOD,MACARONI&CHS,TODD | Baby Foods | 170 | g | Yes | 112 | Yes | 2300.032 |

### Exceeding the DV for low nutrients based on CAC per 100 g or mL by consuming small amounts of foods per day

Exceeding the DV for saturated fat in low saturated fat and low cholesterol foods under CAC per 100 g or mL was started by consuming 1334 g in one food from the group of baked products (NDB number 18388) and two foods from the group of legumes and legume products (NDB numbers 16556 and 16618) (Table 9). Exceeding the DV for saturated fat in these three low saturated fat and low cholesterol foods was due to the high saturated fat amount of low saturated fat and low cholesterol claims.

**Table 9:** Exceeding the daily value for saturated fat in three low saturated fat and low cholesterol foods under CAC per 100 g or mL by consuming 1334 g.

| NDB No | Short description | Food group | Energy (kcal/100 g) | Cholesterol (mg/100 g) | Saturated fat (g/100 g) | Energy from saturated fat (%) | Low saturated fat | Low cholesterol | Saturated fat (g/1334 g) |
| --- | --- | --- | --- | --- | --- | --- | --- | --- | --- |
| 18388 | MUFFINS,WHEAT BRAN,TOASTER-TYPE W/RAISINS,TSTD | Baked Products | 313 | 9 | 1.5 | 4.313% | Yes | Yes | 20.01 |
| 16556 | MORNINGSTAR FARMS CHIK'N NUGGETS,FRZ,UNPREP | Legumes and Legume Products | 221 | 0 | 1.5 | 6.109% | Yes | Yes | 20.01 |
| 16618 | MORNINGSTAR FARMS SPICY INDIAN VEGGIE BURGER,FRZ,UNPREP | Legumes and Legume Products | 189 | 0 | 1.5 | 7.143% | Yes | Yes | 20.01 |

Exceeding the DV for cholesterol in low cholesterol foods under CAC per 100 g or mL was started by consuming 1501 g in one food from the group of finfish and shellfish products (NDB number 15138) (Table 10). Exceeding the DV for cholesterol in this low cholesterol food was due to the high cholesterol amount of the low cholesterol claim.

**Table 10:**
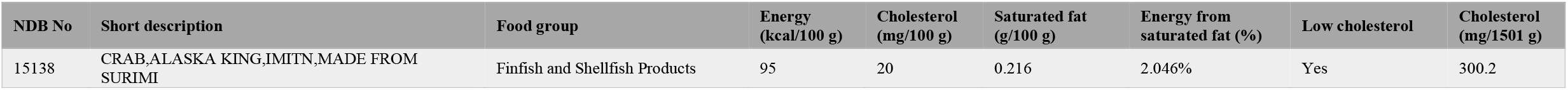
Exceeding the daily value for cholesterol in one low cholesterol food under CAC per 100 g or mL by consuming 1501 g.

| NDB No | Short description | Food group | Energy (kcal/100 g) | Cholesterol (mg/100 g) | Saturated fat (g/100 g) | Energy from saturated fat (%) | Low cholesterol | Cholesterol (mg/1501 g) |
| --- | --- | --- | --- | --- | --- | --- | --- | --- |
| 15138 | CRAB,ALASKA KING,IMITN,MADE FROM SURIMI | Finfish and Shellfish Products | 95 | 20 | 0.216 | 2.046% | Yes | 300.2 |

Exceeding the DV for sodium in low sodium foods under CAC per 100 g or mL was started by consuming 1722 mL in one food from the group of dairy and egg products (NDB number 1292) (Table 11). Exceeding the DV for sodium in this low sodium food was due to the high sodium amount of the low sodium claim in liquid foods.

**Table 11:** Exceeding the daily value for sodium in one low sodium food under CAC per 100 g or mL by consuming 1722 mL.

| NDB No | Short description | Food group | Density (g/mL) | Sodium (mg/100 g) | Sodium (mg/100 mL) | Low sodium | Sodium (mg/1722 mL) |
| --- | --- | --- | --- | --- | --- | --- | --- |
| 1292 | MILK,CHOC,FAT FREE,W/ ADDED VIT A & VITAMIN D | Dairy and Egg Products | 1.056689 | 110 | 116.2358 | Yes | 2001.581 |

Exceeding the DV for total fat in low total fat foods under CAC per 100 g or mL was started by consuming 2223 g in one food from the group of dairy and egg products (NDB number 1294) and one food from the group of sweets (NDB number 19232) (Table 12). Exceeding the DV for total fat in these two low total fat foods was due to the large RACC of these two foods.

**Table 12:** Exceeding the daily value for total fat in two low total fat foods under CAC per 100 g or mL by consuming 2223 g.

| NDB No | Short description | Food group | Total fat (g/100 g) | Low total fat | Total fat (g/2223 g) |
| --- | --- | --- | --- | --- | --- |
| 1294 | YOGURT,GREEK,FRUIT,WHL MILK | Dairy and Egg Products | 3 | Yes | 66.69 |
| 19232 | FLAN,CARAMEL CUSTARD,DRY MIX,PREP W/ WHL MILK | Sweets | 3 | Yes | 66.69 |

### Presence of saturated fat free, but not low saturated fat, foods based on CAC per 100 g or mL

On the basis of CAC per 100 g or mL, saturated fat free and low saturated fat claims are defined as absent and 10% or less of energy from saturated fat, respectively. Thus, some foods are saturated fat free, but not low saturated fat, based on CAC per 100 g or mL, even though the free claim amount of saturated fat is much lower than the low claim amount of saturated fat. According to CAC per 100 g or mL, three foods from the group of soups, sauces, and gravies (NDB numbers 6475, 6476, and 6480) are saturated fat free, but not low saturated fat (Table 13).

**Table 13:**
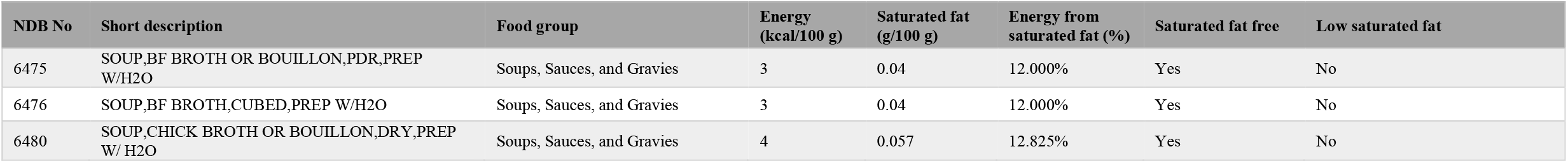
Three saturated fat free, but not low saturated fat, foods based on CAC per 100 g or mL.

### Presence of nutrient free, but not low nutrient, foods based on FDA per serving

On the basis of FDA per serving, if RACC is small, energy, sodium, and total fat are calculated per RACC for free claims and per 50 g of food for low claims. Thus, on the basis of FDA per serving some foods are free of, but not low in, energy, sodium, or total fat, even though the amounts of free claims are much lower than the amounts of low claims.

According to FDA per serving, one food from the group of fats and oils (NDB number 4679), one food from the group of fruits and fruit juices (NDB number 9216), one food from the group of vegetables and vegetable products (NDB number 11974), three foods from the group of baked products (NDB numbers 18371, 18373, and 18375), six foods from the group of sweets (NDB numbers 19310, 19337, 19868, 19909, 43158, and 44018), and 46 foods from the group of spices and herbs (NDB numbers 2001, 2002, 2003, 2004, 2005, 2006, 2007, 2008, 2009, 2010, 2011, 2012, 2013, 2014, 2015, 2016, 2017, 2018, 2019, 2020, 2021, 2022, 2023, 2024, 2025, 2026, 2027, 2028, 2030, 2031, 2032, 2033, 2034, 2035, 2036, 2037, 2038, 2039, 2041, 2042, 2043, 2049, 2050, 2051, 2066, and 44055) are energy free, but not low energy, foods (Table 14).

**Table 14:**
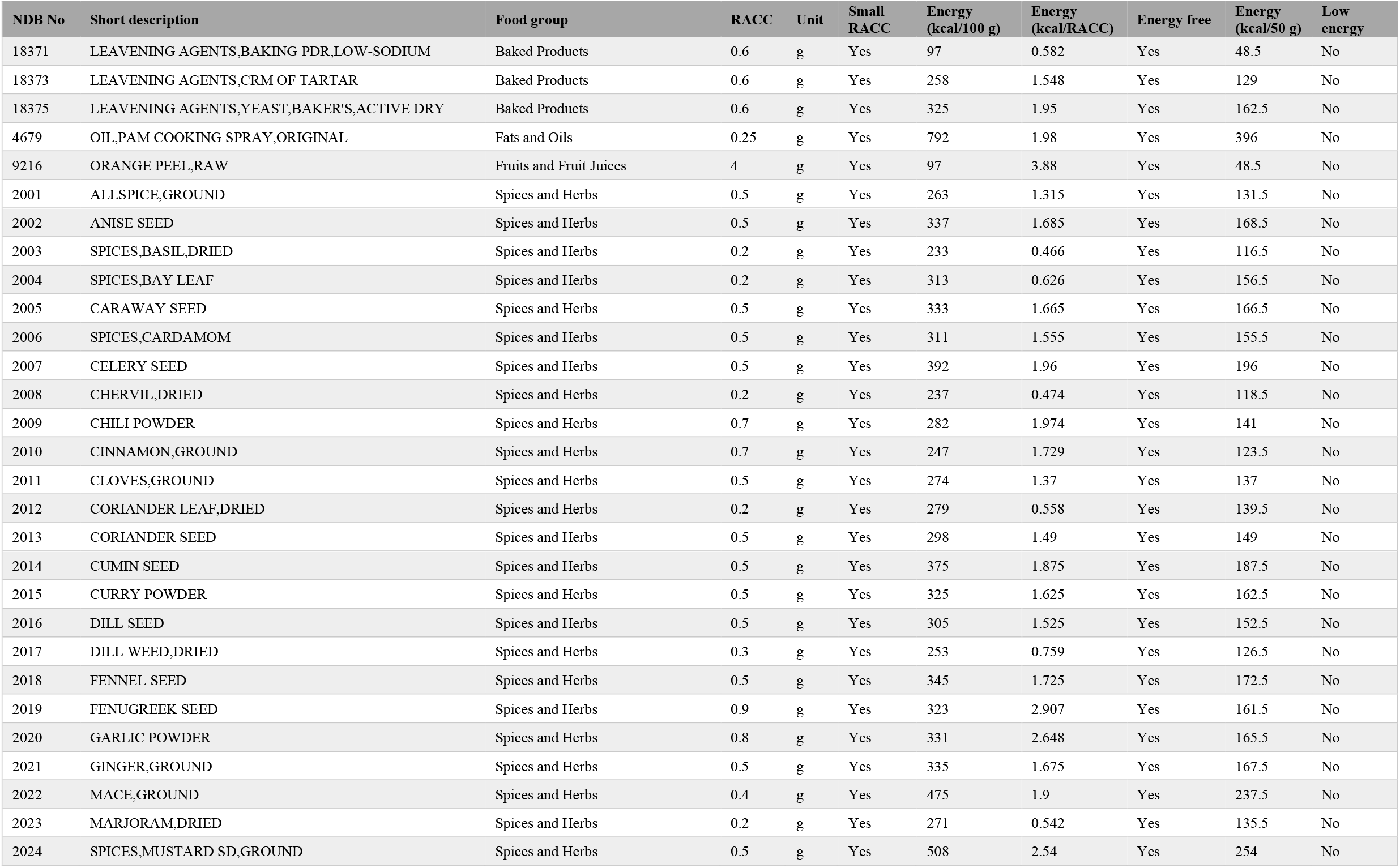
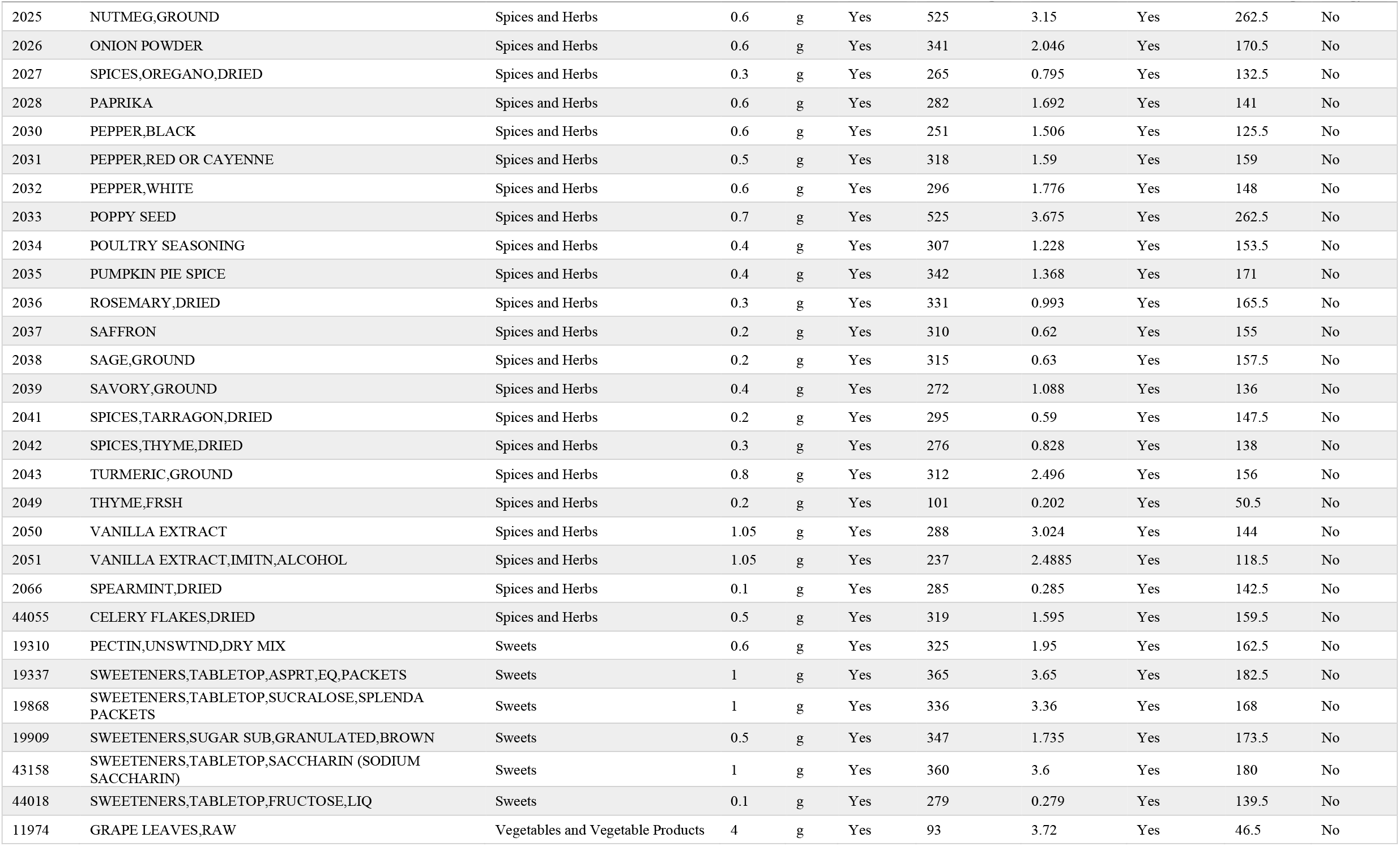
Fifty-eight energy free, but not low energy, foods based on FDA per serving.

According to FDA per serving, one food from the group of fats and oils (NDB number 4679), one food from the group of legumes and legume product (NDB number 16112), two foods from the group of baked products (NDB numbers 18242 and 18375), two foods from the group of dairy and egg products (NDB numbers 1206 and 42136), two foods from the group of vegetables and vegetable products (NDB numbers 11667 and 11978), and 28 foods from the group of spices and herbs (NDB numbers 2001, 2002, 2004, 2005, 2006, 2007, 2009, 2011, 2013, 2014, 2015, 2016, 2018, 2019, 2022, 2023, 2024, 2025, 2028, 2031, 2033, 2034, 2035, 2036, 2038, 2041, 2042, and 2066) are total fat free, but not low total fat, foods (Table 15).

**Table 15:** Thirty-six total fat free, but not low total fat, foods based on FDA per serving.

| NDB No | Short description | Food group | RACC | Unit | Small RACC | Total fat (g/100 g) | Total fat (g/RACC) | Total fat free | Total fat (g/50 g) | Low total fat |
| --- | --- | --- | --- | --- | --- | --- | --- | --- | --- | --- |
| 18242 | CROUTONS,PLAIN | Baked Products | 7 | g | Yes | 6.6 | 0.462 | Yes | 3.3 | No |
| 18375 | LEAVENING AGENTS,YEAST,BAKER'S,ACTIVE DRY | Baked Products | 0.6 | g | Yes | 7.61 | 0.04566 | Yes | 3.805 | No |
| 1206 | CREAM SUB,FLAV,PDR | Dairy and Egg Products | 2 | g | Yes | 21.47 | 0.4294 | Yes | 10.735 | No |
| 42136 | CREAM SUB,PDR,LT | Dairy and Egg Products | 2 | g | Yes | 15.7 | 0.314 | Yes | 7.85 | No |
| 4679 | OIL,PAM COOKING SPRAY,ORIGINAL | Fats and Oils | 0.25 | g | Yes | 78.69 | 0.196725 | Yes | 39.345 | No |
| 16112 | MISO | Legumes and Legume Products | 5.7 | g | Yes | 6.01 | 0.34257 | Yes | 3.005 | No |
| 2001 | ALLSPICE,GROUND | Spices and Herbs | 0.5 | g | Yes | 8.69 | 0.04345 | Yes | 4.345 | No |
| 2002 | ANISE SEED | Spices and Herbs | 0.5 | g | Yes | 15.9 | 0.0795 | Yes | 7.95 | No |
| 2004 | SPICES,BAY LEAF | Spices and Herbs | 0.2 | g | Yes | 8.36 | 0.01672 | Yes | 4.18 | No |
| 2005 | CARAWAY SEED | Spices and Herbs | 0.5 | g | Yes | 14.59 | 0.07295 | Yes | 7.295 | No |
| 2006 | SPICES,CARDAMOM | Spices and Herbs | 0.5 | g | Yes | 6.7 | 0.0335 | Yes | 3.35 | No |
| 2007 | CELERY SEED | Spices and Herbs | 0.5 | g | Yes | 25.27 | 0.12635 | Yes | 12.635 | No |
| 2009 | CHILI POWDER | Spices and Herbs | 0.7 | g | Yes | 14.28 | 0.09996 | Yes | 7.14 | No |
| 2011 | CLOVES,GROUND | Spices and Herbs | 0.5 | g | Yes | 13 | 0.065 | Yes | 6.5 | No |
| 2013 | CORIANDER SEED | Spices and Herbs | 0.5 | g | Yes | 17.77 | 0.08885 | Yes | 8.885 | No |
| 2014 | CUMIN SEED | Spices and Herbs | 0.5 | g | Yes | 22.27 | 0.11135 | Yes | 11.135 | No |
| 2015 | CURRY POWDER | Spices and Herbs | 0.5 | g | Yes | 14.01 | 0.07005 | Yes | 7.005 | No |
| 2016 | DILL SEED | Spices and Herbs | 0.5 | g | Yes | 14.54 | 0.0727 | Yes | 7.27 | No |
| 2018 | FENNEL SEED | Spices and Herbs | 0.5 | g | Yes | 14.87 | 0.07435 | Yes | 7.435 | No |
| 2019 | FENUGREEK SEED | Spices and Herbs | 0.9 | g | Yes | 6.41 | 0.05769 | Yes | 3.205 | No |
| 2022 | MACE,GROUND | Spices and Herbs | 0.4 | g | Yes | 32.38 | 0.12952 | Yes | 16.19 | No |
| 2023 | MARJORAM,DRIED | Spices and Herbs | 0.2 | g | Yes | 7.04 | 0.01408 | Yes | 3.52 | No |
| 2024 | SPICES,MUSTARD SD,GROUND | Spices and Herbs | 0.5 | g | Yes | 36.24 | 0.1812 | Yes | 18.12 | No |
| 2025 | NUTMEG,GROUND | Spices and Herbs | 0.6 | g | Yes | 36.31 | 0.21786 | Yes | 18.155 | No |
| 2028 | PAPRIKA | Spices and Herbs | 0.6 | g | Yes | 12.89 | 0.07734 | Yes | 6.445 | No |
| 2031 | PEPPER,RED OR CAYENNE | Spices and Herbs | 0.5 | g | Yes | 17.27 | 0.08635 | Yes | 8.635 | No |
| 2033 | POPPY SEED | Spices and Herbs | 0.7 | g | Yes | 41.56 | 0.29092 | Yes | 20.78 | No |
| 2034 | POULTRY SEASONING | Spices and Herbs | 0.4 | g | Yes | 7.53 | 0.03012 | Yes | 3.765 | No |
| 2035 | PUMPKIN PIE SPICE | Spices and Herbs | 0.4 | g | Yes | 12.6 | 0.0504 | Yes | 6.3 | No |
| 2036 | ROSEMARY,DRIED | Spices and Herbs | 0.3 | g | Yes | 15.22 | 0.04566 | Yes | 7.61 | No |
| 2038 | SAGE,GROUND | Spices and Herbs | 0.2 | g | Yes | 12.75 | 0.0255 | Yes | 6.375 | No |
| 2041 | SPICES,TARRAGON,DRIED | Spices and Herbs | 0.2 | g | Yes | 7.24 | 0.01448 | Yes | 3.62 | No |
| 2042 | SPICES,THYME,DRIED | Spices and Herbs | 0.3 | g | Yes | 7.43 | 0.02229 | Yes | 3.715 | No |
| 2066 | SPEARMINT,DRIED | Spices and Herbs | 0.1 | g | Yes | 6.03 | 0.00603 | Yes | 3.015 | No |
| 11667 | SEAWEED,SPIRULINA,DRIED | Vegetables and Vegetable Products | 5 | g | Yes | 7.72 | 0.386 | Yes | 3.86 | No |
| 11978 | PEPPERS,ANCHO,DRIED | Vegetables and Vegetable Products | 5 | g | Yes | 8.2 | 0.41 | Yes | 4.1 | No |

According to FDA per serving, one food from the group of spices and herbs (NDB number 2066) and two foods from the group of sweets (NDB numbers 19909 and 43158) are sodium free, but not low sodium, foods (Table 16).

**Table 16:** Three sodium free, but not low sodium, foods based on FDA per serving.

| NDB No | Short description | Food group | RACC | Unit | Small RACC | Sodium (mg/100 g) | Sodium (mg/RACC) | Sodium free | Sodium (mg/50 g) | Low sodium |
| --- | --- | --- | --- | --- | --- | --- | --- | --- | --- | --- |
| 2066 | SPEARMINT,DRIED | Spices and Herbs | 0.1 | g | Yes | 344 | 0.344 | Yes | 172 | No |
| 19909 | SWEETENERS,SUGAR SUB,GRANULATED,BROWN | Sweets | 0.5 | g | Yes | 572 | 2.86 | Yes | 286 | No |
| 43158 | SWEETENERS,TABLETOP,SACCHARIN (SODIUM SACCHARIN) | Sweets | 1 | g | Yes | 428 | 4.28 | Yes | 214 | No |

### Shortcomings of FDA definition for small RACC

There are shortcomings of FDA definition for small RACC (small RACC means RACC of 30 g or less or 2 tablespoons or less) as follows: (1) since foods can have different densities, RACCs for 2 tablespoons of some foods classified as small RACC under FDA definition, may be more than 30 g. For example, RACCs for 88 foods classified as small RACC under FDA definition, are 2 tablespoons, even though 2 tablespoons of these foods are more than 30 g (Table 17); (2) since foods can have different densities, RACCs for more than 2 tablespoons of some foods not classified as small RACC under FDA definition, may be less than 30 g. For example, RACCs for 11 foods not classified as small RACC under FDA definition, are more than 2 tablespoons, even though more than 2 tablespoons of these foods are less than 30 g (Table 18); and (3) RACCs for 2 tablespoons of some foods classified as small RACC under FDA definition, may be less than 30 g, so the nutrient content of these foods is calculated per 50 g of food. In contrast, RACCs for some foods not classified as small RACC under FDA definition, may be 31-49 g, so the nutrient content of these foods is calculated per less than 50 g of food.

**Table 17:**
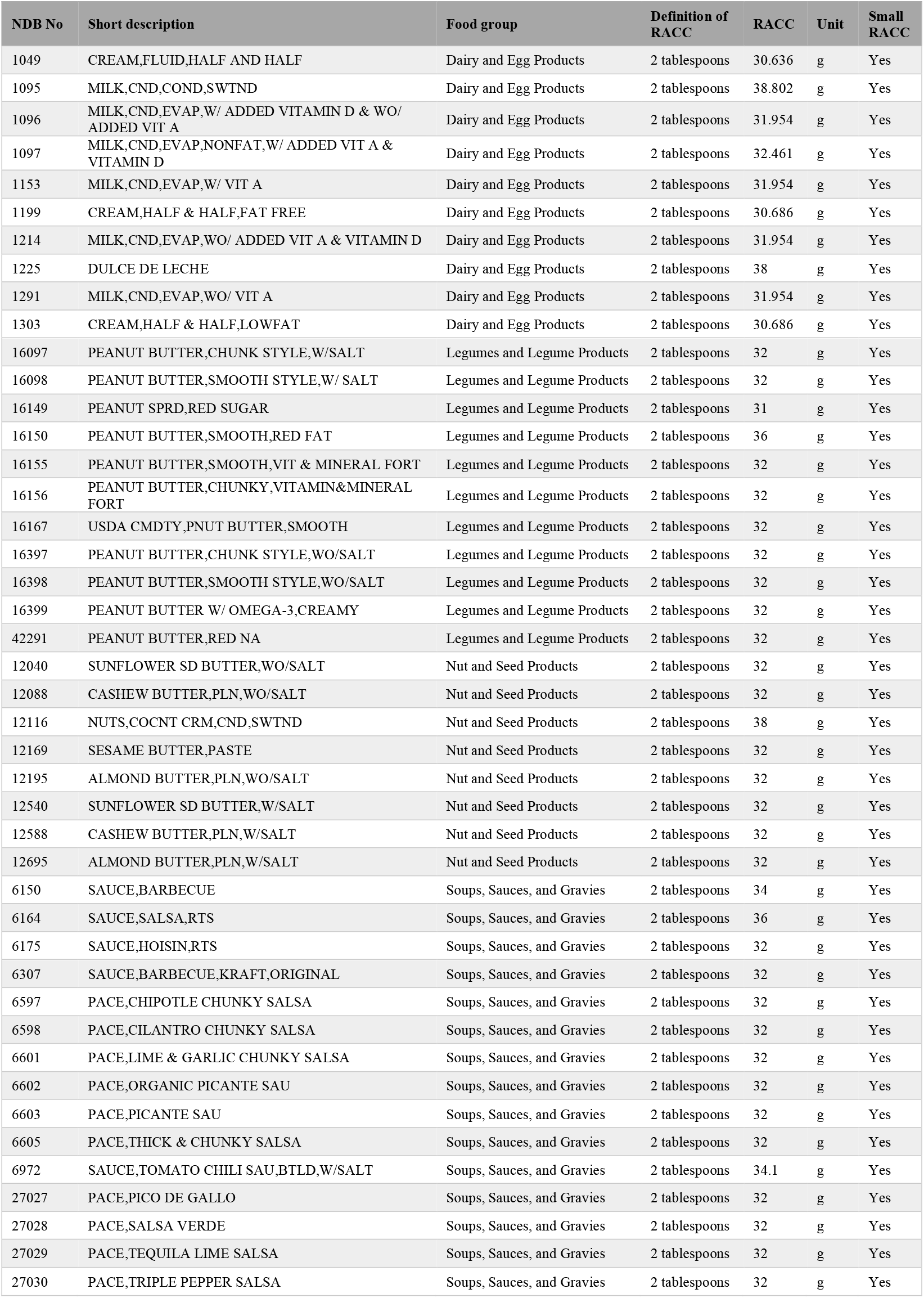
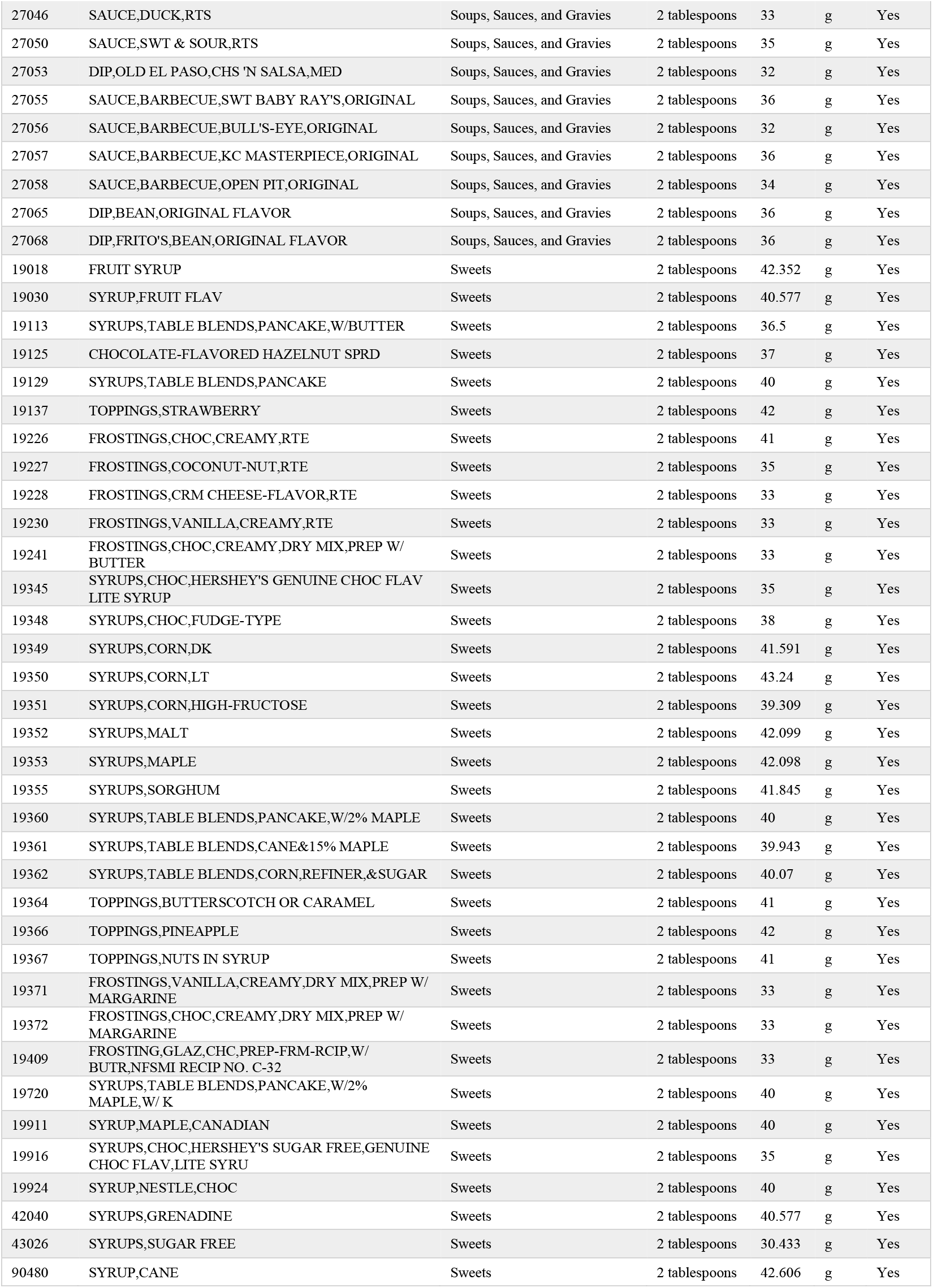
Eighty-eight foods with RACCs more than 30 g classified as small RACC under FDA definition.

**Table 18:** Eleven foods with RACCs less than 30 g not classified as small RACC under FDA definition.

| NDB No | Short description | Food group | Definition of RACC | RACC | Unit | Small RACC |
| --- | --- | --- | --- | --- | --- | --- |
| 20087 | WHEAT, SPROUTED | Cereal Grains and Pasta | 1/4 cup | 27 | g | No |
| 11001 | ALFALFA SEEDS, SPROUTED, RAW | Vegetables and Vegetable Products | 1/4 cup | 8.3 | g | No |
| 11043 | MUNG BNS, MATURE SEEDS, SPROUTED, RAW | Vegetables and Vegetable Products | 1/4 cup | 26 | g | No |
| 11046 | BEANS, NAVY, MATURE SEEDS, SPROUTED, RAW | Vegetables and Vegetable Products | 1/4 cup | 26 | g | No |
| 11248 | LENTILS, SPROUTED, RAW | Vegetables and Vegetable Products | 1/4 cup | 19.3 | g | No |
| 11452 | SOYBEANS, MATURE SEEDS, SPROUTED, RAW | Vegetables and Vegetable Products | 1/4 cup | 17.5 | g | No |
| 11453 | SOYBEANS, MATURE SEEDS, SPROUTED, CKD, STMD | Vegetables and Vegetable Products | 1/4 cup | 23.5 | g | No |
| 11454 | SOYBEANS, MATURE SEEDS, SPROUTED, CKD, STIR-FRIED | Vegetables and Vegetable Products | 1/4 cup | 17.5 | g | No |
| 11676 | RADISH SEEDS, SPROUTED, RAW | Vegetables and Vegetable Products | 1/4 cup | 9.5 | g | No |
| 11923 | SOYBEANS, MATURE SEEDS, SPROUTED, CKD, STMD, W/SALT | Vegetables and Vegetable Products | 1/4 cup | 23.5 | g | No |
| 11924 | SOYBEANS, MATURE SEEDS, SPROUTED, CKD, STIR-FRIED, W/SALT | Vegetables and Vegetable Products | 1/4 cup | 17.5 | g | No |

### Calculation of nutrient content in inappropriate amounts of foods based on CAC per 100 kcal

Nutrient content values for source and high claims under CAC per 100 kcal were calculated in all 25 food groups with a high average amount of foods. On the basis of CAC per 100 kcal, nutrient content values for source and high claims are calculated per 100 kcal of foods. Source and high claims are based on 20 eating occasions per day in CAC per 100 kcal and 10 eating occasions per day in the proposed method. According to source and high claims, 200 kcal of food in CAC per 100 kcal is equivalent (not the same quantity) to one serving in the proposed method. Since 200 kcal of foods in CAC per 100 kcal may be more or less than servings in the proposed method, the nutrient content per 200 kcal of foods may be more or less than the nutrient content of servings in the proposed method. 200 kcal of food is calculated by formula 10.

**Furmula 10:** 200 kcal of food _(g)_ = (200 × 100) ÷ energy _(kcal/100 g)_

For example, if raw spinach contains 23 kcal of energy per 100 g, the weight of 200 kcal of raw spinach is calculated by formula 10 as follows.

200 kcal of food: (200 × 100) ÷ 23 = 869.57 _(g)_

According to source and high claims, the average of 200 kcal of foods based on CAC per 100 kcal was higher than the average of servings based on the proposed method in all 25 food groups (spices and herbs; beverages; vegetables and vegetable products; fats and oils; baby foods; soups, sauces, and gravies; sweets; fruits and fruit juices; American Indian/Alaska Native foods; finfish and shellfish products; baked products; dairy and egg products; nut and seed products; legumes and legume products; sausages and luncheon meats; snacks; cereal grains and pasta; poultry products; pork products; lamb, veal, and game products; breakfast cereals; beef products; fast foods; restaurant foods; meals, entrees, and side dishes) (Figure 5). The 100 kcal basis may be in excess of the portion typically consumed, notably for vegetables and fruits (Drewnowski *et al*., 2009).

**Figure 5:**
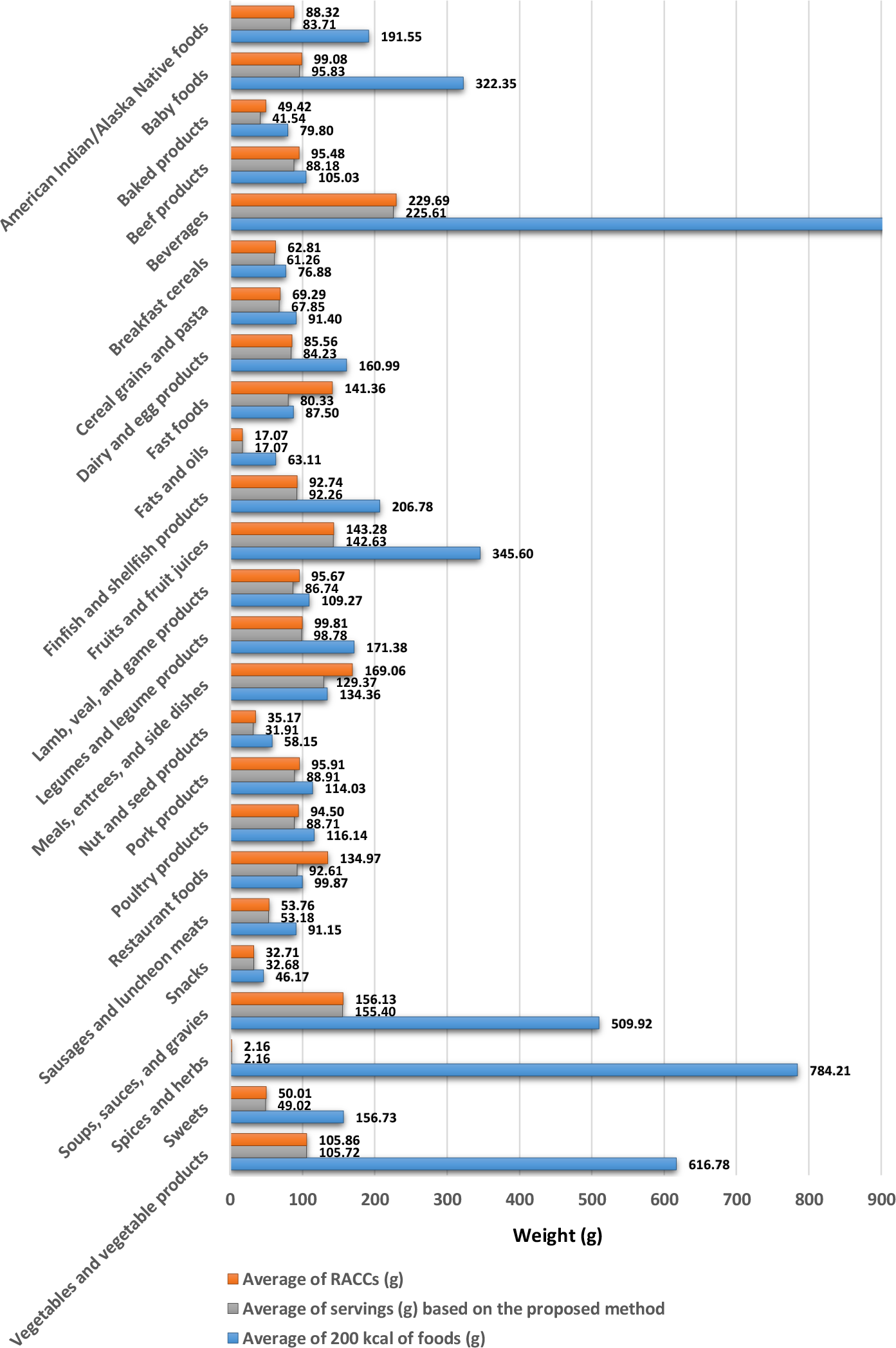
Average of 200 kcal of foods based on CAC per 100 kcal, average of servings based on the proposed method, and average of RACCs based on FDA and CAC per serving for source and high claims in food groups

### Calculation of nutrient content in inappropriate amounts of foods based on FDA and CAC per serving

Nutrient content values for source and high claims under FDA and CAC per serving were calculated in 23 food groups with a high average amount of foods. According to source and high claims, the average of RACCs was higher than the average of servings based on the proposed method in 23 food groups (fast foods; restaurant foods; meals, entrees, and side dishes; baked products; lamb, veal, and game products; nut and seed products; beef products; pork products; poultry products; American Indian/Alaska Native foods; baby foods; breakfast cereals; cereal grains and pasta; sweets; beverages; dairy and egg products; sausages and luncheon meats; legumes and legume products; finfish and shellfish products; soups, sauces, and gravies; fruits and fruit juices; vegetables and vegetable products; snacks) and the average of RACCs and the average of servings based on the proposed method were the same in two food groups (fats and oils; spices and herbs) (Figure 5).

### Calculation of nutrient content in inappropriate amounts of foods based on CAC per 100 g or mL

Nutrient content values for source and high claims under CAC per 100 g or mL were calculated in 20 food groups with a high average amount of foods and five food groups with a low average amount of foods. Since 100 g or mL of foods may be more or less than servings in the proposed method, the nutrient content of foods per 100 g or mL may be more or less than servings in the proposed method. According to source and high claims, the average of 100 g or mL of foods was higher than the average of servings based on the proposed method in 20 food groups (spices and herbs; fats and oils; nut and seed products; snacks; baked products; sweets; sausages and luncheon meats; breakfast cereals; cereal grains and pasta; fast foods; dairy and egg products; American Indian/Alaska Native foods; lamb, veal, and game products; beef products; poultry products; pork products; finfish and shellfish products; restaurant foods; baby foods; legumes and legume products) and the average of 100 g or mL of foods was lower than the average of servings based on the proposed method in five food groups (beverages; soups, sauces, and gravies; fruits and fruit juices; meals, entrees, and side dishes; vegetables and vegetable products).

### High sodium amount of the low sodium claim for liquid foods per 100 g based on CAC per 100 g or mL

The high sodium amount of the low sodium claim for liquid foods and determination of the low sodium claim for liquid foods per 100 g based on CAC per 100 g or mL have increased low sodium liquid foods. On the basis of CAC per 100 g or mL, low claims for cholesterol, energy, saturated fat, and total fat are determined per 100 g in solid foods and per 100 mL in liquid foods, but the low sodium claim is determined per 100 g in solid and liquid foods. In addition, amounts of low claims for cholesterol, energy, saturated fat, and total fat in liquid foods are half of solid foods, but the sodium amount of the low sodium claim in solid and liquid foods is the same. For example, if sodium amounts of the low sodium claim were 120 mg or less per 100 g in solid foods and 60 mg or less per 100 mL in liquid foods, 83.27% of liquid foods would be low sodium. Since the sodium amount of the low sodium claim based on CAC per 100 g or mL in solid and liquid foods is 120 mg or less per 100 g, 96.83% of liquid foods are low sodium.

### Determination of the sodium free claim for liquid foods per 100 g based on CAC per 100 g or mL

Determination of the sodium free claim for liquid foods per 100 g based on CAC per 100 g or mL has increased sodium free liquid foods. On the basis of CAC per 100 g or mL, free claims for cholesterol, energy, saturated fat, and total fat are determined per 100 g in solid foods and per 100 mL in liquid foods, but the sodium free claim is determined per 100 g in solid and liquid foods. Also, amounts of free claims for cholesterol, energy, saturated fat, sodium, and total fat in solid and liquid foods are the same. For example, if the sodium amount of the sodium free claim was 5 mg or less per 100 mL in liquid foods, 27.46% of liquid foods would be sodium free. Since the sodium amount of the sodium free claim based on CAC per 100 g or mL is 5 mg or less per 100 g in liquid foods, 30.99% of liquid foods are sodium free.

### High dietary fiber amount of the dietary fiber source claim for liquid foods per 100 g based on CAC per 100 g or mL

On the basis of CAC per 100 g or mL, the high dietary fiber amount of the dietary fiber source claim for liquid foods and determination of the dietary fiber source claim for liquid foods per 100 g have decreased dietary fiber source liquid foods. On the basis of CAC per 100 g or mL, source and high claims for vitamins, minerals, and protein are determined per 100 g in solid foods and per 100 mL in liquid foods, but source and high claims for dietary fiber are determined per 100 g in solid and liquid foods. In addition, amounts of source and high claims for vitamins, minerals, and protein in liquid foods are half of solid foods, but dietary fiber amounts of source and high claims in solid and liquid foods are the same. For example, if the dietary fiber source claim was 5-9% of the DV per 100 mL in liquid foods, 1.49% of liquid foods would be dietary fiber source. Since the dietary fiber source claim based on CAC per 100 g or mL is 10-19% of the DV per 100 g in liquid foods, 0% of liquid foods are dietary fiber source.

### Presence of energy from saturated fat for cholesterol free and low cholesterol claims based on CAC per 100 g or mL

The presence of energy from saturated fat for cholesterol free and low cholesterol claims based on CAC per 100 g or mL has decreased cholesterol free and low cholesterol foods. On the basis of CAC per 100 g or mL, cholesterol free and low cholesterol claims are defined as 10% or less of energy from saturated fat, although energy from saturated fat does not raise blood cholesterol levels. Difference between the absence and presence of energy from saturated fat (not more than 10% of energy from saturated fat) at CAC per 100 g or mL for cholesterol free and low cholesterol foods was seen in seven food groups (soups, sauces, and gravies; baby foods; meals, entrees, and side dishes; dairy and egg products; fruits and fruit juices; vegetables and vegetable products; sweets) and eight food groups (soups, sauces, and gravies; meals, entrees, and side dishes; baby foods; dairy and egg products; sweets; fruits and fruit juices; vegetables and vegetable products; poultry products), respectively (Figure 6).

**Figure 6:**
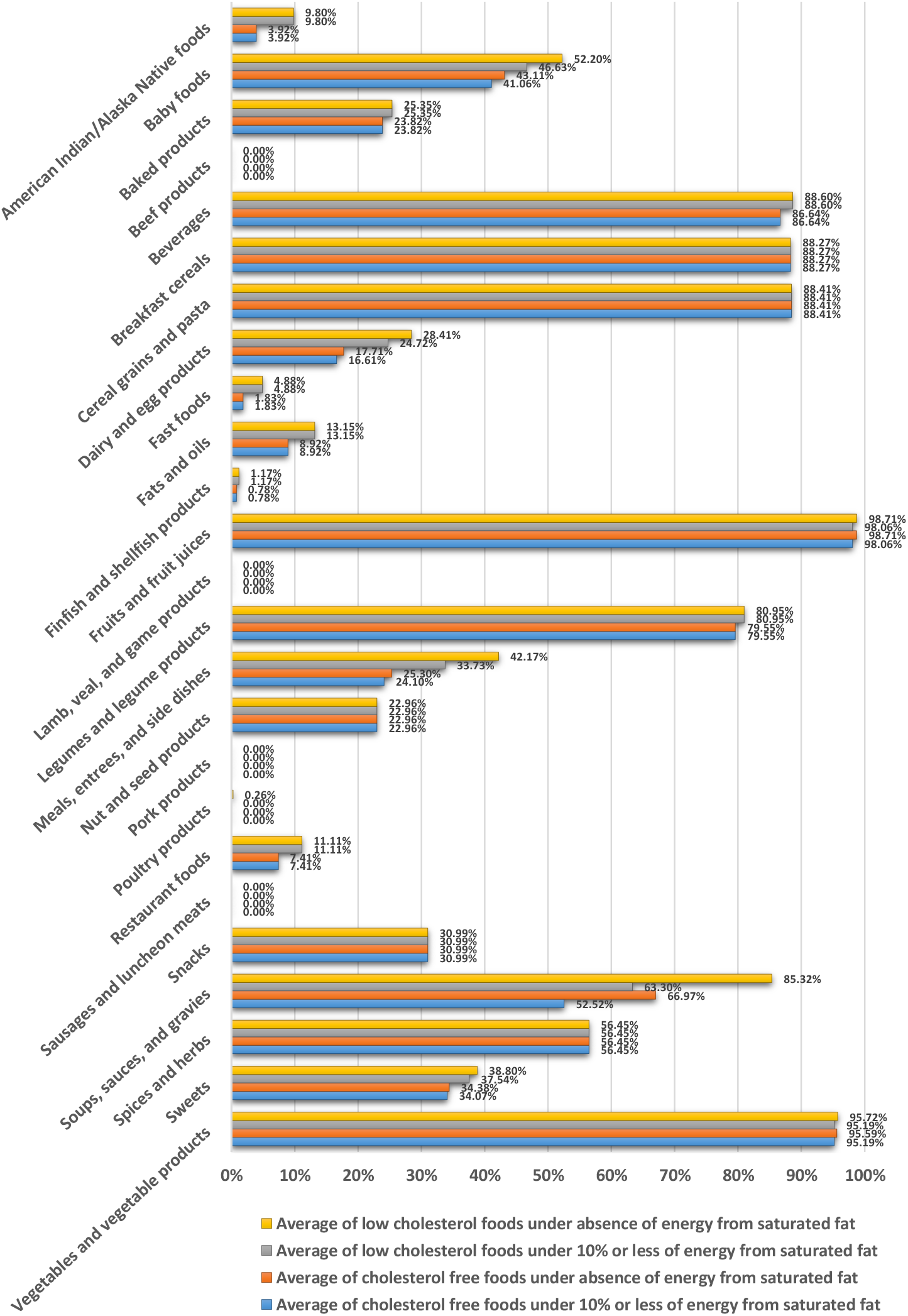
Difference between the absence and presence of energy from saturated fat (not more than 10% of energy from saturated fat) at CAC per 100 g or mL for cholesterol free and low cholesterol foods in food groups

### Absence of energy from saturated fat for the saturated fat free claim based on CAC per 100 g or mL

Absence of energy from saturated fat for the saturated fat free claim based on CAC per 100 g or mL has increased saturated fat free foods. Since energy from saturated fat based on CAC per 100 g or mL for the saturated fat free claim is absent, the portion of saturated fat in energy is not considered for this claim. Difference between the absence and presence of energy from saturated fat (not more than 9% of energy from saturated fat) at CAC per 100 g or mL for saturated fat free foods was seen in two food groups (soups, sauces, and gravies; beverages) (Figure 7).

**Figure 7:**
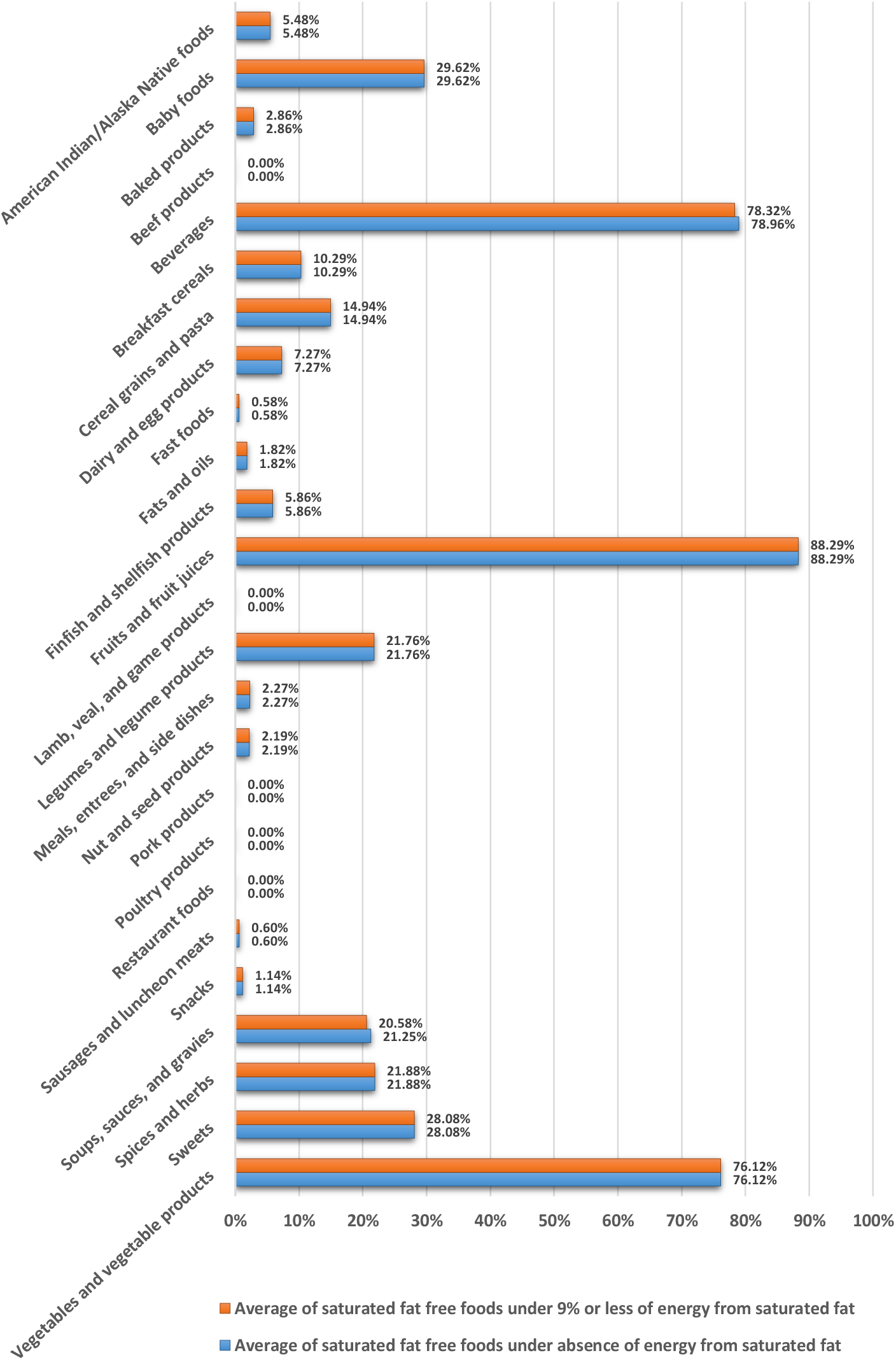
Difference between the absence and presence of energy from saturated fat (not more than 9% of energy from saturated fat) at CAC per 100 g or mL for saturated fat free foods in food groups

### High portion of energy from saturated fat for the low saturated fat claim based on CAC per 100 g or mL

High portion of energy from saturated for the low saturated fat claim based on CAC per 100 g or mL has increased low saturated fat foods. On the basis of CAC per 100 g or mL, the low saturated fat claim is defined as 10% or less of energy from saturated fat, although energy from saturated fat based on the portion of saturated fat in energy is 9%. Difference between 9% or less and 10% or less of energy from saturated fat at CAC per 100 g or mL for low saturated fat foods was seen in 16 food groups (American Indian/Alaska Native foods; soups, sauces, and gravies; pork products; meals, entrees, and side dishes; lamb, veal, and game products; beef products; finfish and shellfish products; sweets; poultry products; spices and herbs; legumes and legume products; beverages; sausages and luncheon meats; baby foods; fast foods; vegetables and vegetable products) (Figure 8).

**Figure 8:**
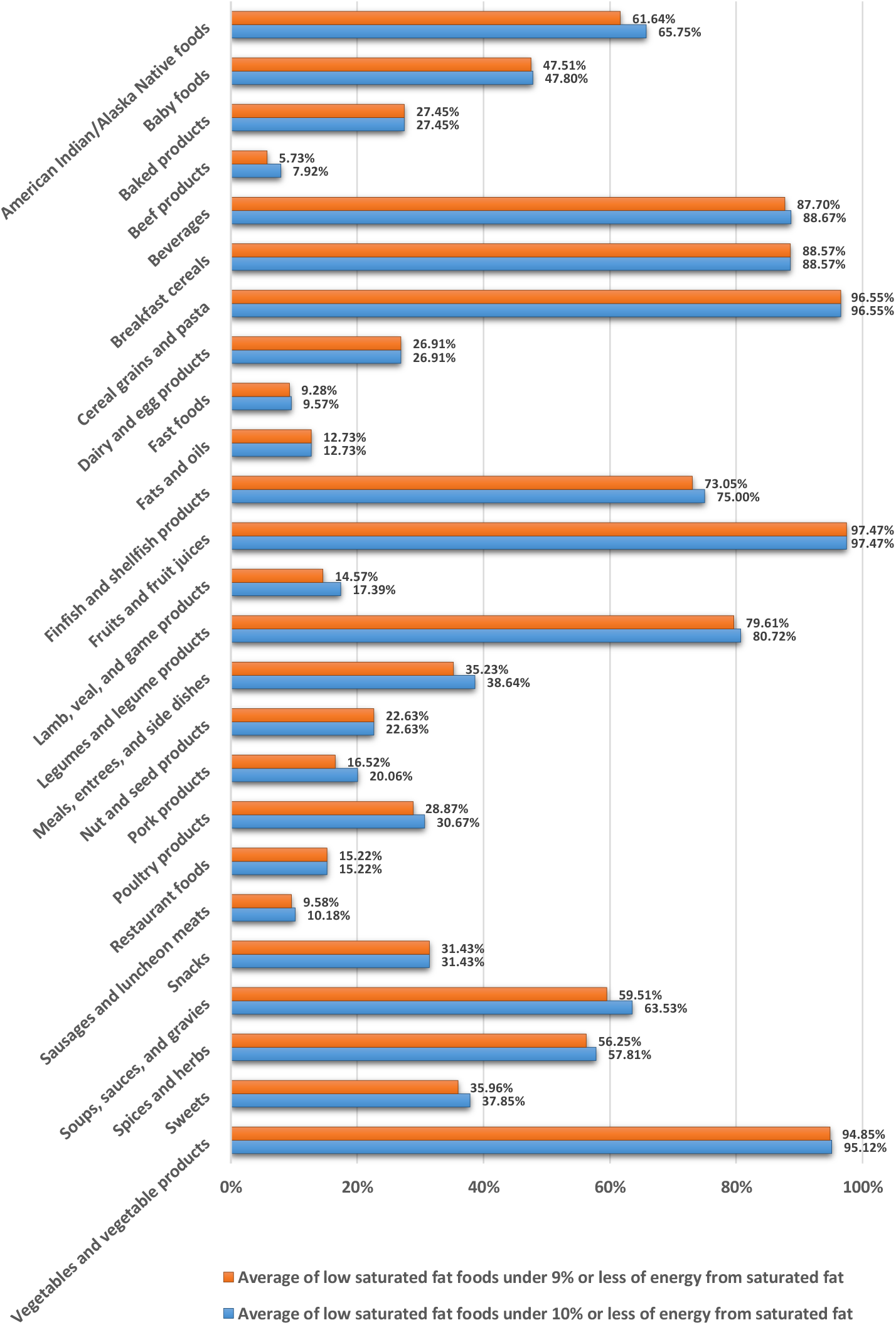
Difference between 9% or less and 10% or less of energy from saturated fat at CAC per 100 g or mL for low saturated fat foods in food groups

### Energy from saturated fat for saturated fat free and low saturated fat claims based on FDA per serving

Absence of energy from saturated fat for the saturated fat free claim based on FDA per serving has increased saturated fat free foods. Since energy from saturated fat based on FDA per serving for the saturated fat free claim is absent, the portion of saturated fat in energy is not considered for this claim. High portion of energy from saturated fat for the low saturated fat claim based on FDA per serving has increased low saturated fat foods. On the basis of FDA per serving, the low saturated fat claim is defined as 15% or less of energy from saturated fat in all foods excluding meals and main dishes and less than 10% of energy from saturated fat in meals and main dishes. However, energy from saturated fat based on the portion of saturated fat in energy is 9%. Since portion of energy from saturated fat under FDA per serving for the low saturated fat claim is high, the portion of saturated fat in energy is considered high for this claim. Difference between energy from saturated fat as FDA per serving and 9% or less of energy from saturated fat at FDA per serving for low saturated fat foods was seen in 18 food groups (soups, sauces, and gravies; spices and herbs; fats and oils; sausages and luncheon meats; baby foods; meals, entrees, and side dishes; dairy and egg products; legumes and legume products; beverages; fruits and fruit juices; baked products; finfish and shellfish products; pork products; fast foods; cereal grains and pasta; vegetables and vegetable products; poultry products; lamb, veal, and game products) (Figure 9).

**Figure 9:**
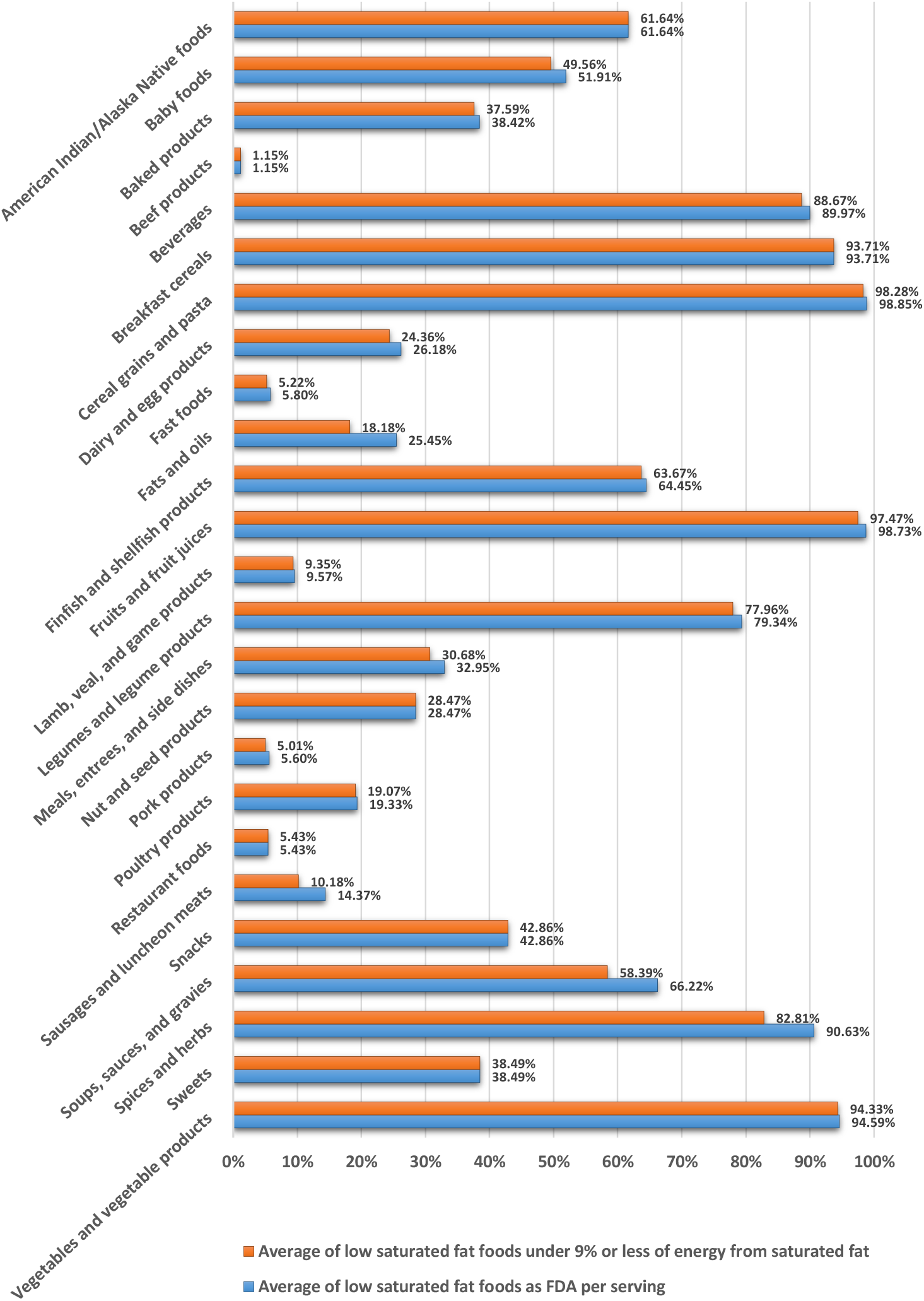
Difference between energy from saturated fat as FDA per serving (15% or less of energy from saturated fat in all foods excluding meals and main dishes and less than 10% of energy from saturated fat in meals and main dishes) and 9% or less of energy from saturated fat at FDA per serving for low saturated fat foods in food groups

### Absence of energy from total fat for total fat free and low total fat claims based on CAC per 100 g or mL

Absence of energy from total fat for total fat free and low total fat claims based on CAC per 100 g or mL has increased total fat free and low total fat foods. Since energy from total fat based on CAC per 100 g or mL for total fat free and low total fat claims is absent, the portion of total fat in energy is not considered for these claims. Difference between the absence and presence of energy from total fat (not more than 30% of energy from total fat) at CAC per 100 g or mL for total fat free and low total fat foods was seen in three food groups (soups, sauces, and gravies; beverages; vegetables and vegetable products) and 12 food groups (soups, sauces, and gravies; baby foods; legumes and legume products; restaurant foods; spices and herbs; finfish and shellfish products; fats and oils; beverages; vegetables and vegetable products; sweets; poultry products; lamb, veal, and game products), respectively (Figure 10).

**Figure 10:**
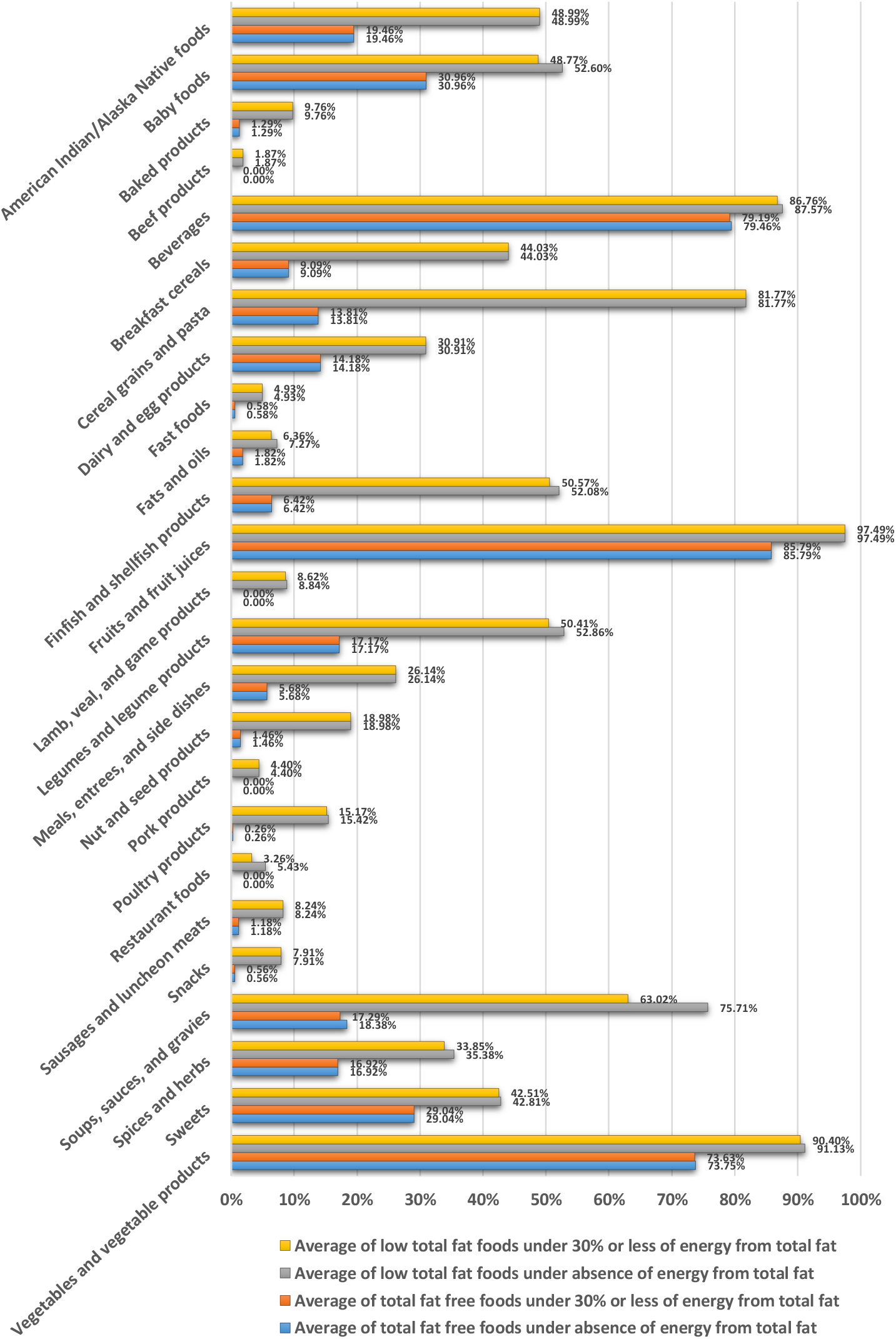
Difference between the absence and presence of energy from total fat (not more than 30% of energy from total fat) at CAC per 100 g or mL for total fat free and low total fat foods in food groups

### Energy from total fat for total fat free and low total fat claims based on FDA per serving

On the basis of FDA per serving, the absence of energy from total fat for the low total fat claim (in all foods excluding meals and main dishes) has increased low total fat foods. Also, the absence of energy from total fat for the total fat free claim has increased total fat free foods. Energy from total fat based on FDA per serving is absent in all foods for the total fat free claim and in all foods excluding meals and main dishes for the low total fat claim. Since energy from total fat based on FDA per serving for the total fat free claim is absent, the portion of total fat in energy is not considered for this claim. Also, since energy from total fat based on FDA per serving for the low total fat claim is absent in all foods excluding meals and main dishes, the portion of total fat in energy for the low total fat claim is not considered in all foods excluding meals and main dishes. On the basis of FDA per serving, low portion of energy from total fat for the low total fat claim in meals and main dishes has decreased low total fat foods in meals and main dishes. On the basis of FDA per serving, the low total fat claim is defined as 30% or less of energy from total fat in meals and main dishes, although energy from total fat based on the portion of total fat in energy is 35%. Difference between energy from total fat as FDA per serving and 35% or less of energy from total fat at FDA per serving for total fat free and low total fat foods was seen in six food groups (spices and herbs; fats and oils; soups, sauces, and gravies; dairy and egg products; beverages; vegetables and vegetable products) and 11 food groups (soups, sauces, and gravies; spices and herbs; dairy and egg products; fats and oils; legumes and legume products; beverages; vegetables and vegetable products; baby foods; sweets; pork products; fast foods), respectively (Figure 11).

**Figure 11:**
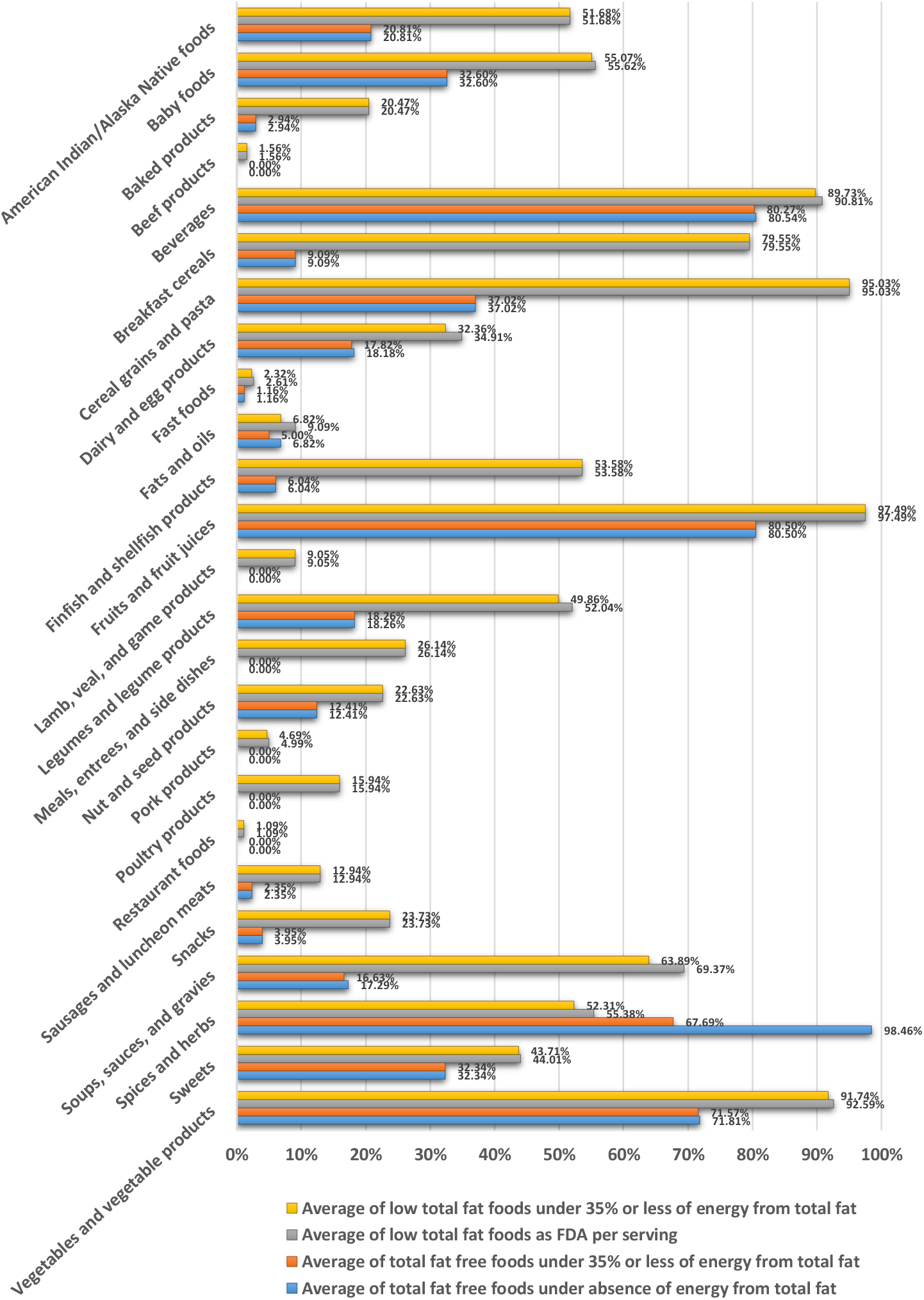
Difference between energy from total fat as FDA per serving (the absence of energy from total fat for the total fat free claim in all foods, the absence of energy from total fat for the low total fat claim in all foods excluding meals and main dishes, and 30% or less of energy from total fat for the low total fat claim in meals and main dishes) and 35% or less of energy from total fat at FDA per serving for total fat free and low total fat foods in food groups

### Calcium source and high calcium foods

Calcium as a nutrient is most commonly associated with the formation and metabolism of bone (IOM, 2011). Calcium in the circulatory system, extracellular fluid, muscle, and other tissues is critical for mediating vascular contraction and vasodilatation, muscle function, nerve transmission, intracellular signaling, and hormonal secretion (IOM, 2011). Calcium is classically associated with dairy products; milk, yogurt, and cheese are rich sources of calcium, providing the major share of calcium from foods in the general diet in the United States and Canada (IOM, 2011). According to the proposed method, the average of calcium source and high calcium foods in food groups was as follows: it was good in one food group (dairy and egg products); it was almost good in one food group (baby foods); it was acceptable in two food groups (breakfast cereals; fast foods); it was almost insufficient in seven food groups (meals, entrees, and side dishes; legumes and legume products; restaurant foods; beverages; sweets; snacks; American Indian/Alaska Native foods); it was insufficient in 10 food groups (vegetables and vegetable products; baked products; finfish and shellfish products; soups, sauces, and gravies; nut and seed products; fruits and fruit juices; sausages and luncheon meats; poultry products; pork products; beef products); and it was absent in four food groups (cereal grains and pasta; fats and oils; lamb, veal, and game products; spices and herbs) (Figure 12). High calcium and calcium source foods based on the proposed method are given in Tables S1 and S2, respectively.

**Figure 12:**
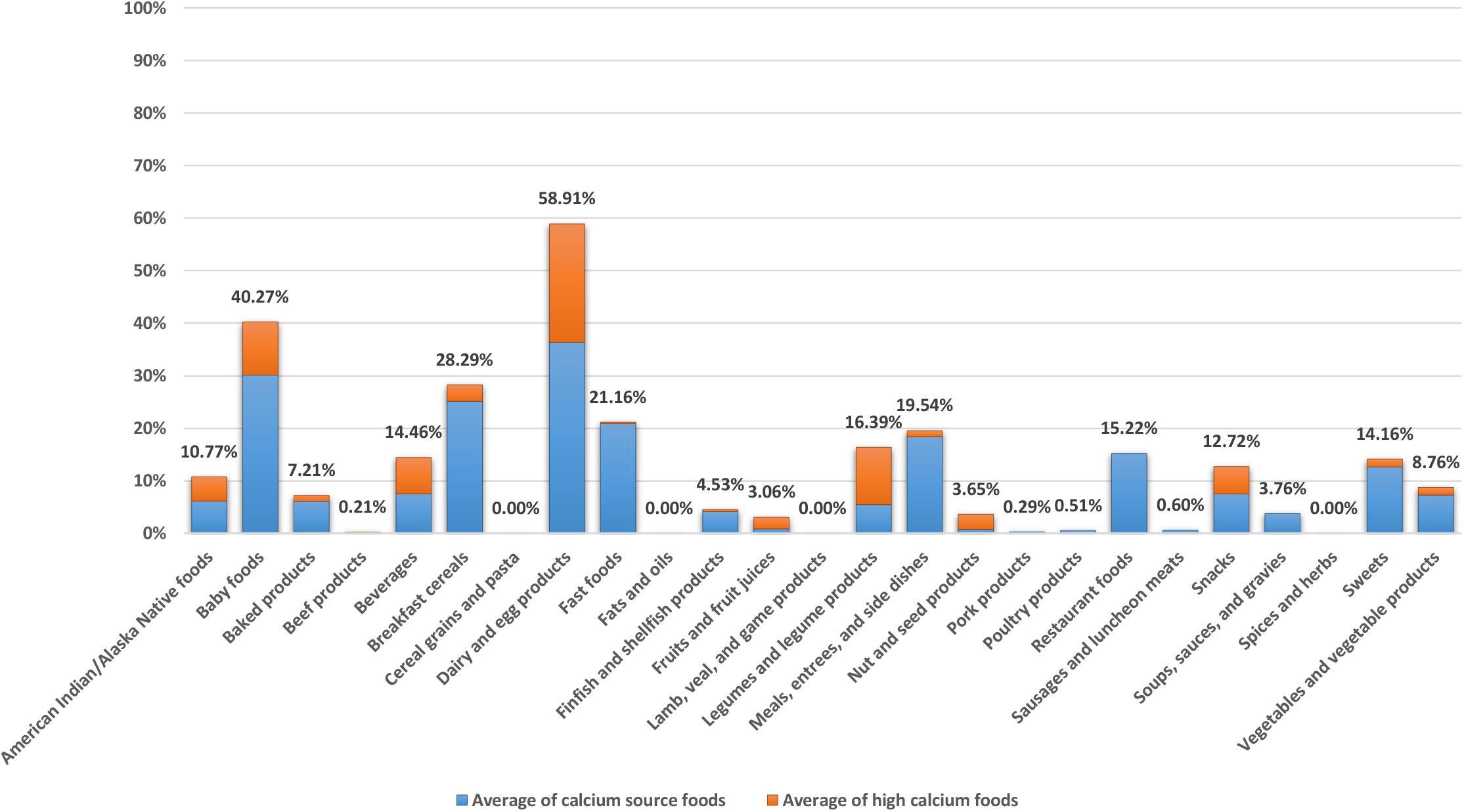
Average of calcium source and high calcium foods based on the proposed method in food groups. All calcium source and high calcium foods except the calcium source and high calcium baby foods are based on the reference energy intake of 2,000 kcal for adults and children aged 4 years and older. The calcium source and high calcium baby foods are based on the reference energy intake of 1,000 kcal for children 1 through 3 years of age.

### Cholesterol free and low cholesterol foods

Cholesterol plays an important role in steroid hormone and bile acid biosynthesis and serves as an integral component of cell membranes (IOM, 2005b). Cholesterol is found in all animal tissues, so that some are present in all foods of animal origin (Sardesai, 2003). The majority of cholesterol is consumed from eggs and meat (FASEB, 1995). There is much evidence to indicate a positive linear trend between cholesterol intake and low density lipoprotein (LDL) cholesterol concentration, and therefore increased risk of coronary heart disease (IOM, 2005b). According to the proposed method, the average of cholesterol free foods in food groups was as follows: it was excellent in four food groups (fruits and fruit juices; vegetables and vegetable products; breakfast cereals; cereal grains and pasta); it was almost excellent in two food groups (spices and herbs; beverages); it was very good in one food group (legumes and legume products); it was almost good in one food group (snacks); it was satisfactory in three food groups (baby foods; sweets; soups, sauces, and gravies); it was acceptable in two food groups (baked products; nut and seed products); it was almost insufficient in one food group (fats and oils); it was insufficient in six food groups (meals, entrees, and side dishes; dairy and egg products; American Indian/Alaska Native foods; restaurant foods; fast foods; finfish and shellfish products); and it was absent in five food groups (beef products; lamb, veal, and game products; pork products; poultry products; sausages and luncheon meats). Also, the average of low cholesterol foods in food groups was as follows: it was excellent in four food groups (fruits and fruit juices; vegetables and vegetable products; breakfast cereals; cereal grains and pasta); it was almost excellent in two food groups (beverages; spices and herbs); it was very good in one food group (legumes and legume products); it was almost very good in one food group (soups, sauces, and gravies); it was almost good in two food groups (snacks; baby foods); it was satisfactory in two food groups (sweets; baked products); it was acceptable in four food groups (nut and seed products; dairy and egg products; fats and oils; meals, entrees, and side dishes); it was insufficient in five food groups (American Indian/Alaska Native foods; fast foods; restaurant foods; finfish and shellfish products; sausages and luncheon meats); and it was absent in four food groups (beef products; lamb, veal, and game products; pork products; poultry products) (Figure 13). Cholesterol free and low cholesterol foods based on the proposed method are given in Tables 3 and S4, respectively.

**Figure 13:**
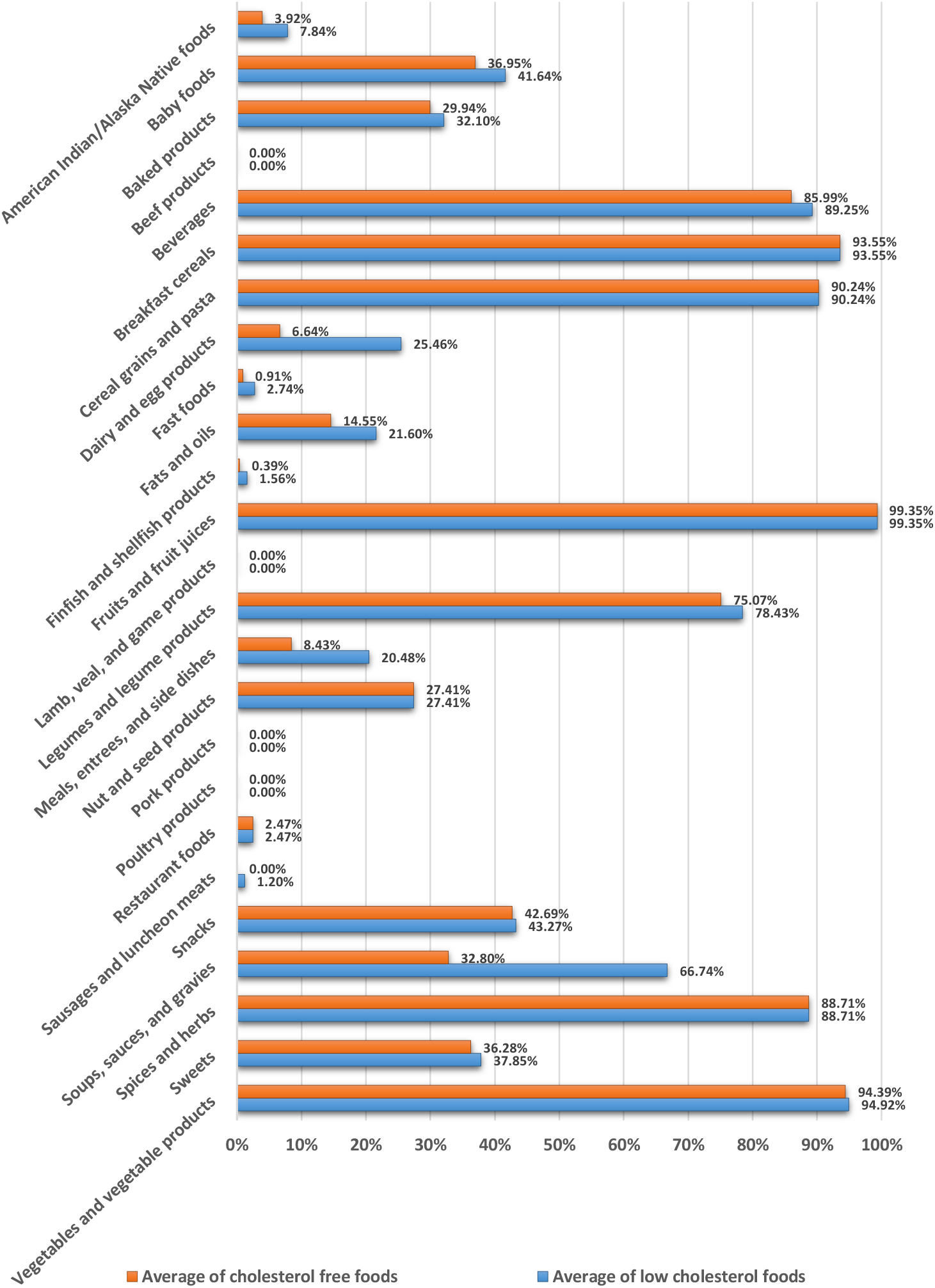
Averages of cholesterol free and low cholesterol foods based on the proposed method in food groups. All cholesterol free and low cholesterol foods except the cholesterol free and low cholesterol baby foods are based on the reference energy intake of 2,000 kcal for adults and children aged 4 years and older. The cholesterol free and low cholesterol baby foods are based on the reference energy intake of 1,000 kcal for children 1 through 3 years of age.

### Choline source and high choline foods

Choline functions as a precursor for acetylcholine, phospholipids, and the methyl donor betaine (IOM, 1998). The main dietary sources of choline in the United States consist primarily of animal-based products that are particularly rich in choline—meat, poultry, fish, dairy products, and eggs (Chester *et al*., 2011; Hollenbeck, 2012; Leermakers *et al*., 2015; Sanders and Zeisel, 2007; Zeisel, 2014). According to the proposed method, the average of choline source and high choline foods in food groups was as follows: it was excellent in one food group (lamb, veal, and game products); it was almost excellent in three food groups (pork products; finfish and shellfish products; beef products); it was almost very good in one food group (poultry products); it was satisfactory in one food group (American Indian/Alaska Native foods); it was almost insufficient in four food groups (fast foods; baby foods; legumes and legume products; restaurant foods); it was insufficient in eight food groups (dairy and egg products; meals, entrees, and side dishes; sausages and luncheon meats; vegetables and vegetable products; beverages; soups, sauces, and gravies; breakfast cereals; baked products); and it was absent in seven food groups (cereal grains and pasta; fats and oils; fruits and fruit juices; nut and seed products; snacks; spices and herbs; sweets) (Figure 14). High choline and choline source foods based on the proposed method are given in Tables S5 and S6, respectively.

**Figure 14:**
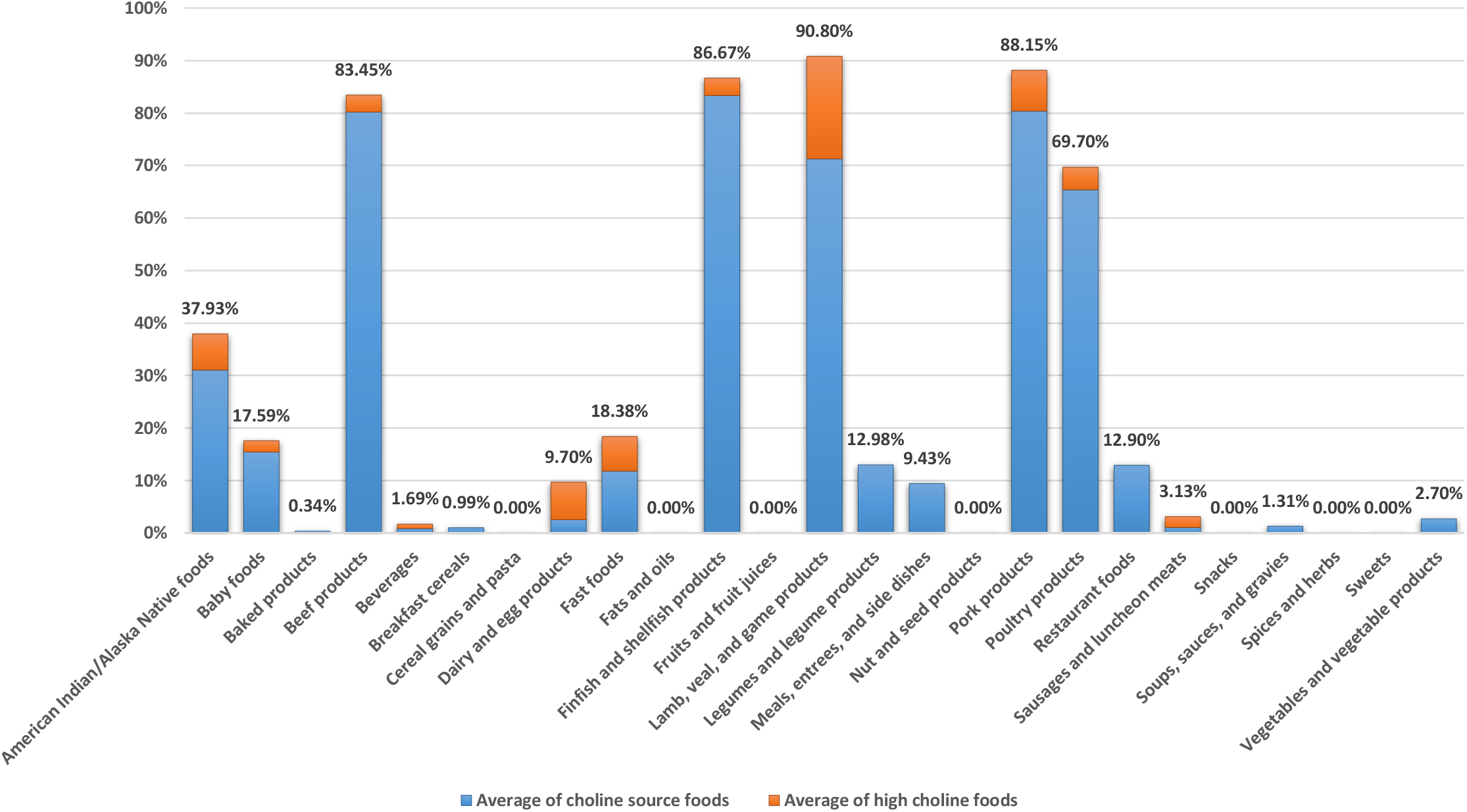
Average of choline source and high choline foods based on the proposed method in food groups. All choline source and high choline foods except the choline source and high choline baby foods are based on the reference energy intake of 2,000 kcal for adults and children aged 4 years and older. The choline source and high choline baby foods are based on the reference energy intake of 1,000 kcal for children 1 through 3 years of age.

### Copper source and high copper foods

Copper in living organisms, including humans, forms an essential component of many enzymes (cuproenzymes) and proteins (Bawa, 2008). The biochemical role for copper is primarily catalytic, with many copper metalloenzymes acting as oxidases to achieve the reduction of molecular oxygen (Bawa, 2008). Foods that contribute substantial amounts of copper to the U.S. diet include those high in copper, such as organ meats, grains, and cocoa products, and those relatively low in copper that are consumed in substantial amounts, such as tea, potatoes, milk, and chicken (IOM, 2001). According to the proposed method, the average of copper source and high copper foods in food groups was as follows: it was excellent in two food groups (legumes and legume products; nut and seed products); it was almost excellent in one food group (baby foods); it was almost very good in one food group (lamb, veal, and game products); it was good in three food groups (breakfast cereals; cereal grains and pasta; meals, entrees, and side dishes); it was almost good in five food groups (fruits and fruit juices; vegetables and vegetable products; soups, sauces, and gravies; snacks; American Indian/Alaska Native foods); it was satisfactory in four food groups (pork products; finfish and shellfish products; poultry products; beef products); it was acceptable in four food groups (restaurant foods; fast foods; sweets; baked products); it was almost insufficient in three food groups (beverages; sausages and luncheon meats; dairy and egg products); and it was absent in two food groups (fats and oils; spices and herbs) (Figure 15). High copper and copper source foods based on the proposed method are given in Tables S7 and S8, respectively.

**Figure 15:**
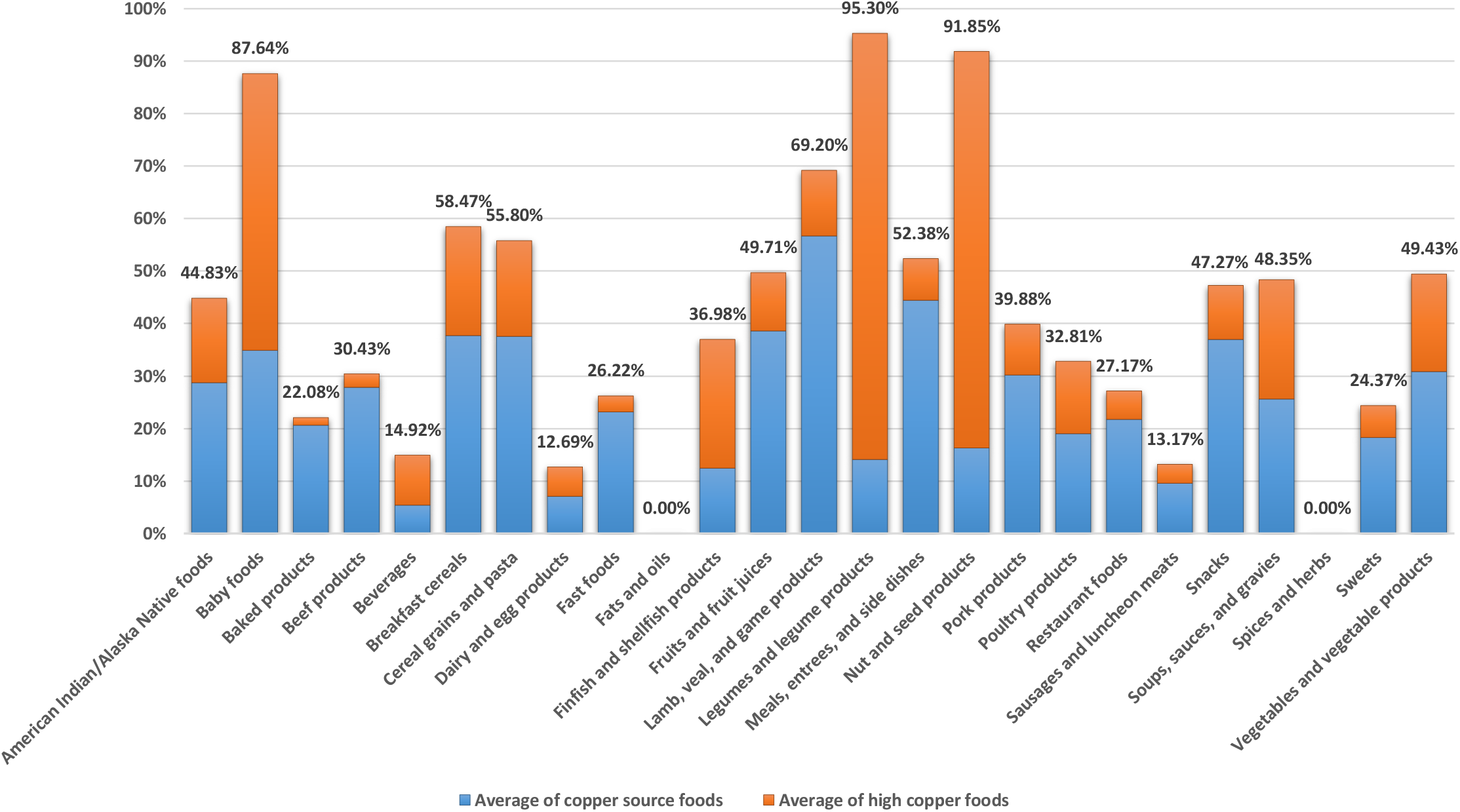
Average of copper source and high copper foods based on the proposed method in food groups. All copper source and high copper foods except the copper source and high copper baby foods are based on the reference energy intake of 2,000 kcal for adults and children aged 4 years and older. The copper source and high copper baby foods are based on the reference energy intake of 1,000 kcal for children 1 through 3 years of age.

### Dietary fiber source and high dietary fiber foods

Dietary fiber consists of nondigestible carbohydrates and lignin that are intrinsic and intact in plants (Slavin, 2003). Fibers have different properties that result in different physiological effects, including laxation, attenuation of blood glucose levels, and normalization of serum cholesterol levels (IOM, 2006). Dietary fiber is found in most fruits, vegetables, legumes, nuts, and grains (IOM, 2006). According to the proposed method, the average of dietary fiber source and high dietary fiber foods in food groups was as follows: it was good in two food groups (breakfast cereals; legumes and legume products); it was satisfactory in six food groups (meals, entrees, and side dishes; American Indian/Alaska Native foods; nut and seed products; vegetables and vegetable products; fruits and fruit juices; cereal grains and pasta); it was acceptable in two food groups (soups, sauces, and gravies; baby foods); it was almost insufficient in two food groups (snacks; baked products); it was insufficient in five food groups (restaurant foods; fast foods; sweets; beverages; dairy and egg products); and it was absent in eight food groups (beef products; fats and oils; finfish and shellfish products; lamb, veal, and game products; pork products; poultry products; sausages and luncheon meats; spices and herbs) (Figure 16). High dietary fiber and dietary fiber source foods based on the proposed method are given in Tables S9 and S10, respectively.

**Figure 16:**
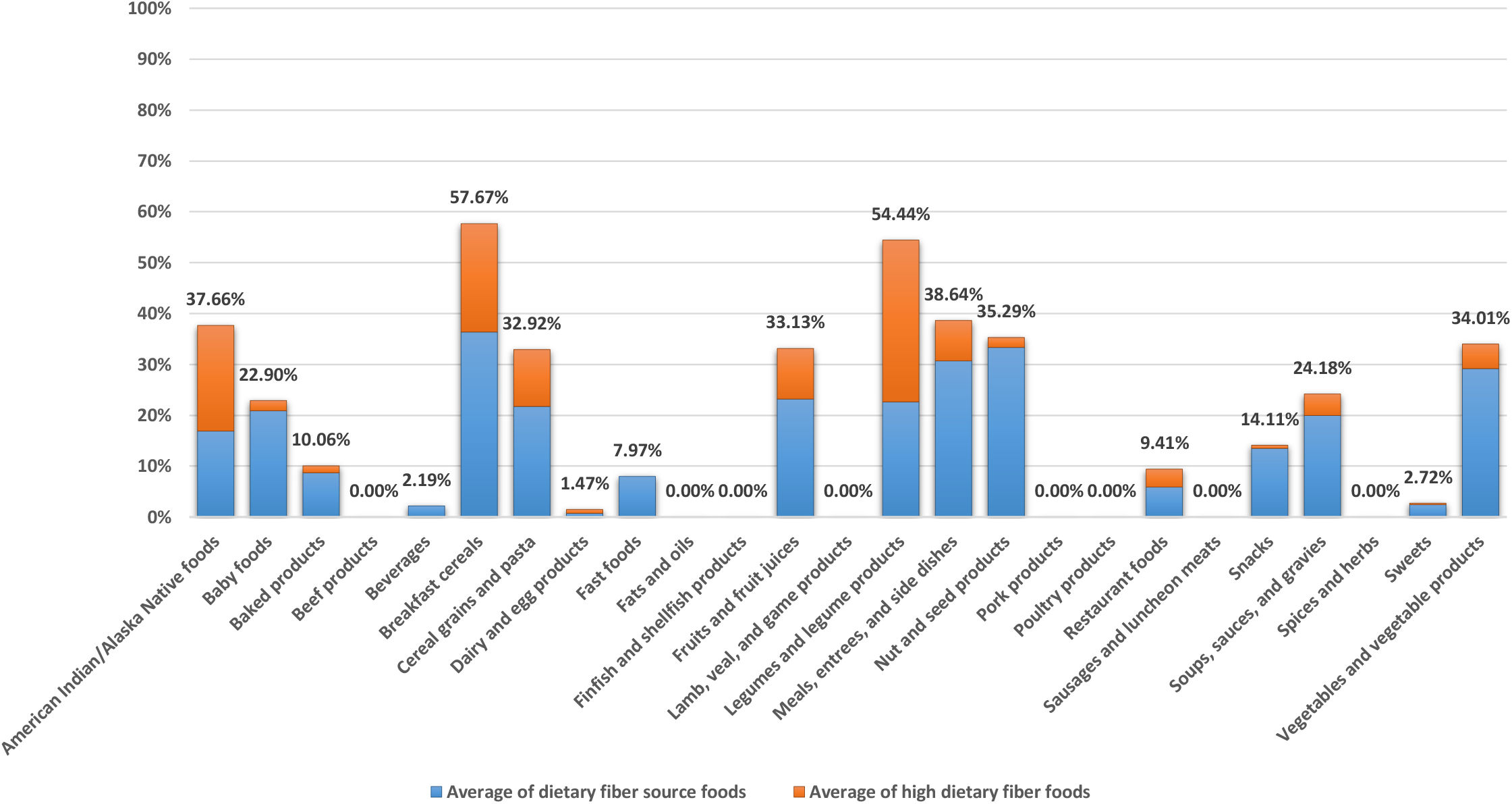
Average of dietary fiber source and high dietary fiber foods based on the proposed method in food groups. All dietary fiber source and high dietary fiber foods except the dietary fiber source and high dietary fiber baby foods are based on the reference energy intake of 2,000 kcal for adults and children aged 4 years and older. The dietary fiber source and high dietary fiber baby foods are based on the reference energy intake of 1,000 kcal for children 1 through 3 years of age.

### Energy free and low energy foods

Energy is required to sustain the body’s various functions, including respiration, circulation, physical work, and maintenance of core body temperature (IOM, 2005b; Willihnganz *et al*., 2019). Carbohydrate, fat, protein, and alcohol provide all of the energy supplied by foods and are generally referred to as macronutrients (in contrast to vitamins and minerals, usually referred to as micronutrients) (IOM, 2005b). On average, carbohydrate, fat, protein, and alcohol provide 4, 9, 4, and 7 kcal per g, respectively. Imbalances between intake and expenditure result in gains or losses of body components, mainly in the form of fat, and these determine changes in body weight (IOM, 2005b). According to the proposed method, the average of energy free foods in food groups was as follows: it was almost insufficient in two food groups (beverages; spices and herbs); it was insufficient in eight food groups (soups, sauces, and gravies; vegetables and vegetable products; fruits and fruit juices; sweets; fats and oils; finfish and shellfish products; baby foods; baked products); and it was absent in 15 food groups (American Indian/Alaska Native foods; beef products; breakfast cereals; cereal grains and pasta; dairy and egg products; fast foods; lamb, veal, and game products; legumes and legume products; meals, entrees, and side dishes; nut and seed products; pork products; poultry products; restaurant foods; sausages and luncheon meats; snacks). Also, the average of low energy foods in food groups was as follows: it was almost excellent in two food groups (vegetables and vegetable products; spices and herbs); it was almost very good in one food group (soups, sauces, and gravies); it was good in one food group (fruits and fruit juices); it was almost good in two food groups (beverages; dairy and egg products); it was satisfactory in two food groups (finfish and shellfish products; American Indian/Alaska Native foods); it was acceptable in two food groups (sausages and luncheon meats; fats and oils); it was almost insufficient in three food groups (sweets; legumes and legume products; nut and seed products); it was insufficient in 11 food groups (baby foods; cereal grains and pasta; pork products; fast foods; snacks; lamb, veal, and game products; baked products; restaurant foods; beef products; poultry products; breakfast cereals); and it was absent in one food group (meals, entrees, and side dishes) (Figure 17). Energy free and low energy foods based on the proposed method are given in Tables S11 and S12, respectively.

**Figure 17:**
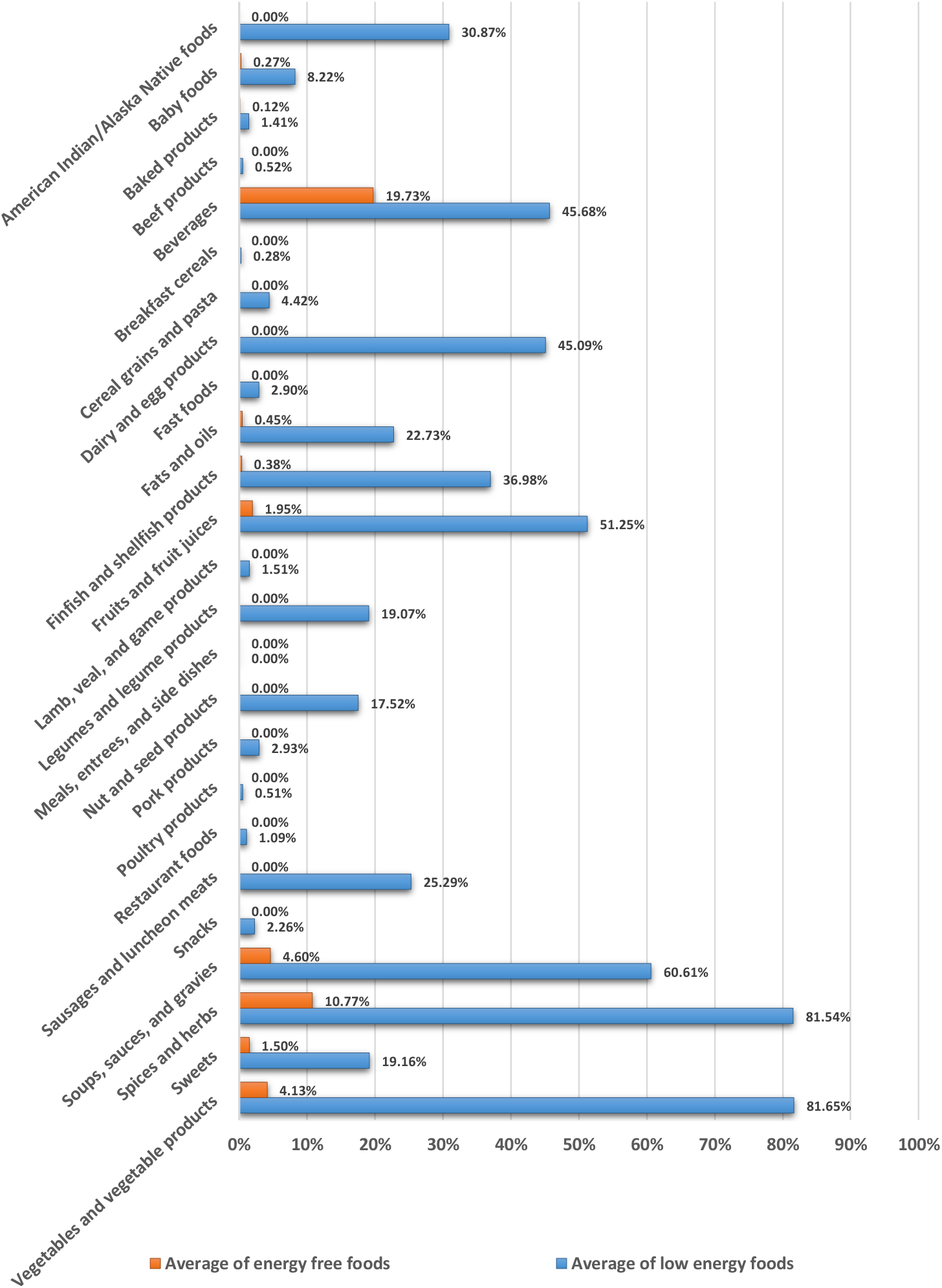
Averages of energy free and low energy foods based on the proposed method in food groups. All energy free and low energy foods except the energy free and low energy baby foods are based on the reference energy intake of 2,000 kcal for adults and children aged 4 years and older. The energy free and low energy baby foods are based on the reference energy intake of 1,000 kcal for children 1 through 3 years of age.

### Folate source and high folate foods

Folate (also known as folacin, folic acid, and vitamin B_9_) functions as a coenzyme in single-carbon transfers in the metabolism of nucleic and amino acids (IOM, 1998). Folate is a generic term that includes both the naturally occurring form of the vitamin (food folate or pteroylpolyglutamates) and the monoglutamate form (folic acid or pteroylmonoglutamic acid), which is used in fortified foods and dietary supplements (IOM, 2006). Orange juice, white breads, dried beans, green salad, and ready-to-eat breakfast cereals are the major food sources of folate on a given day, contributing 37% of total folate intake (Subar *et al*., 1989). According to the proposed method, the average of folate source and high folate foods in food groups was as follows: it was very good in three food groups (breakfast cereals; meals, entrees, and side dishes; fast foods); it was almost very good in one food group (legumes and legume products); it was almost good in two food groups (baby foods; baked products); it was satisfactory in three food groups (cereal grains and pasta; restaurant foods; vegetables and vegetable products); it was acceptable in one food group (snacks); it was almost insufficient in three food groups (American Indian/Alaska Native foods; fruits and fruit juices; nut and seed products); it was insufficient in nine food groups (beverages; soups, sauces, and gravies; finfish and shellfish products; poultry products; dairy and egg products; lamb, veal, and game products; sausages and luncheon meats; pork products; beef products); and it was absent in three food groups (fats and oils; spices and herbs; sweets) (Figure 18). High folate and folate source foods based on the proposed method are given in Tables S13 and S14, respectively.

**Figure 18:**
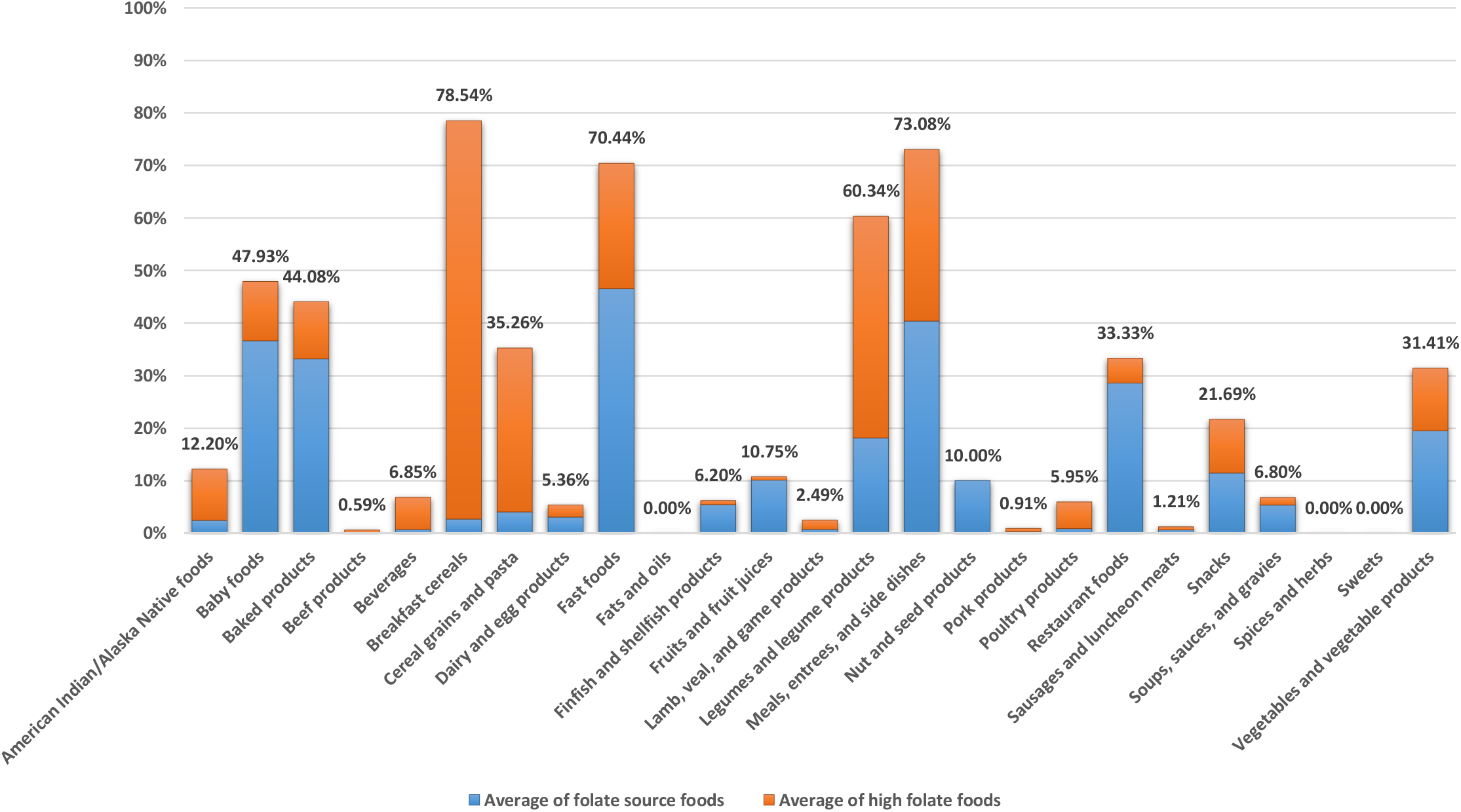
Average of folate source and high folate foods based on the proposed method in food groups. All folate source and high folate foods except the folate source and high folate baby foods are based on the reference energy intake of 2,000 kcal for adults and children aged 4 years and older. The folate source and high folate baby foods are based on the reference energy intake of 1,000 kcal for children 1 through 3 years of age.

### Iron source and high iron foods

Iron functions as a component of a number of proteins, including enzymes and hemoglobin, the latter being important for the transport of oxygen to tissues throughout the body for metabolism (IOM, 2001). About half of the iron from meat, poultry, and fish is heme iron, which is highly bioavailable; the remainder is nonheme, which is less readily absorbed by the body (IOM, 2006). Iron in dairy foods, eggs, and all plant-based foods is entirely nonheme (IOM, 2006). According to the proposed method, the average of iron source and high iron foods in food groups was as follows: it was very good in one food group (breakfast cereals); it was almost very good in one food group (beef products); it was good in one food group (baby foods); it was almost good in two food groups (legumes and legume products; American Indian/Alaska Native foods); it was satisfactory in two food groups (lamb, veal, and game products; meals, entrees, and side dishes); it was acceptable in four food groups (fast foods; poultry products; cereal grains and pasta; nut and seed products); it was almost insufficient in five food groups (baked products; vegetables and vegetable products; restaurant foods; snacks; finfish and shellfish products); it was insufficient in seven food groups (sausages and luncheon meats; soups, sauces, and gravies; fruits and fruit juices; beverages; pork products; dairy and egg products; sweets); and it was absent in two food groups (fats and oils; spices and herbs) (Figure 19). High iron and iron source foods based on the proposed method are given in Tables S15 and S16, respectively.

**Figure 19:**
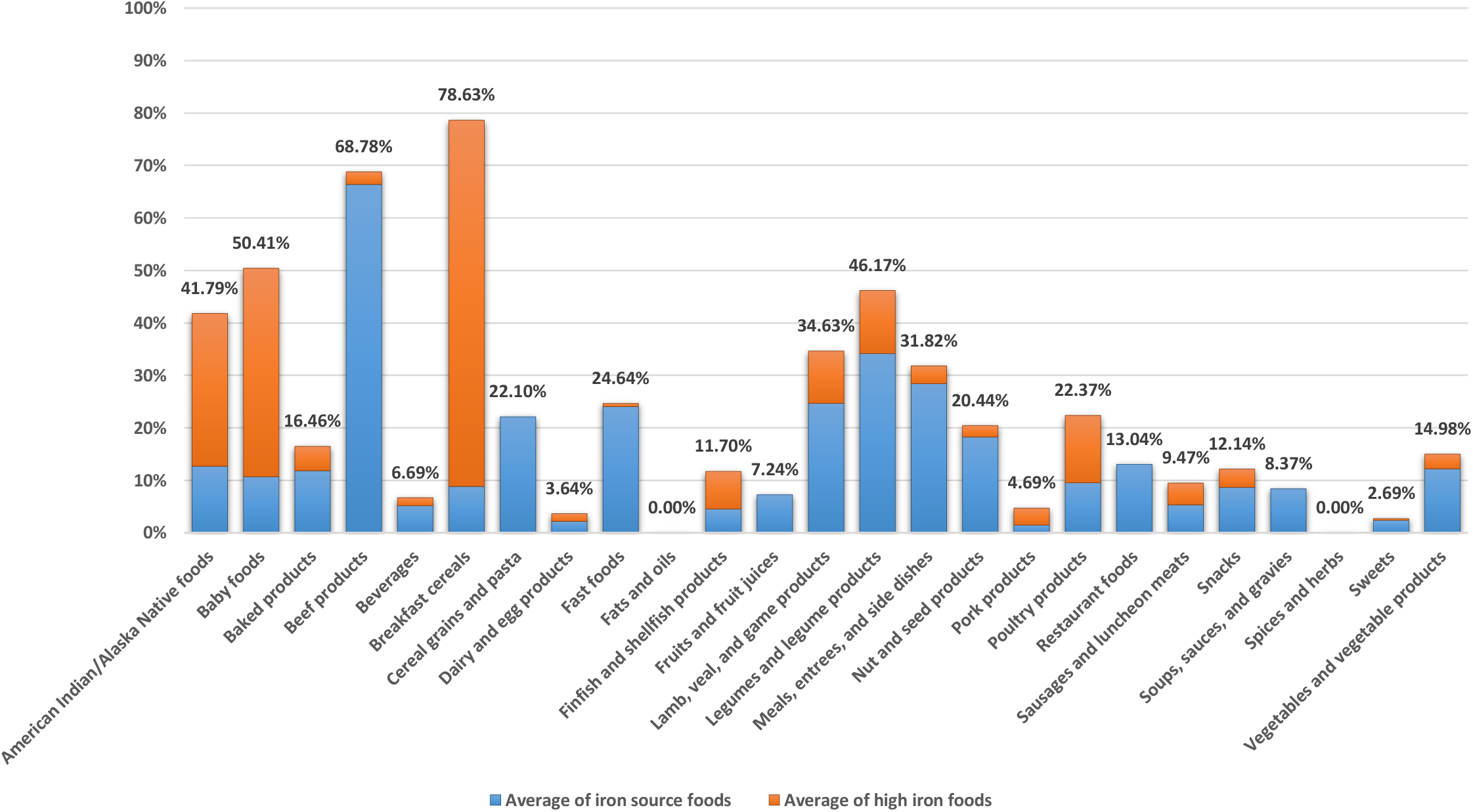
Average of iron source and high iron foods based on the proposed method in food groups. All iron source and high iron foods except the iron source and high iron baby foods are based on the reference energy intake of 2,000 kcal for adults and children aged 4 years and older. The iron source and high iron baby foods are based on the reference energy intake of 1,000 kcal for children 1 through 3 years of age.

### Magnesium source and high magnesium foods

Magnesium is a required cofactor for over 300 enzyme systems (Wacker and Parisi, 1968a; Wacker and Parisi, 1968b; Wacker and Parisi, 1968c), as well as magnesium is involved in bone health and in the maintenance of intracellular levels of potassium and calcium (IOM, 2006). Analyses from the 1989 Total Diet Study of FDA indicated that approximately 45 percent of dietary magnesium was obtained from vegetables, fruits, grains, and nuts, whereas about 29 percent was obtained from milk, meat, and eggs (Pennington and Young, 1991). According to the proposed method, the average of magnesium source and high magnesium foods in food groups was as follows: it was almost very good in one food group (legumes and legume products); it was good in two food groups (nut and seed products; baby foods); it was satisfactory in two food groups (breakfast cereals; cereal grains and pasta); it was almost insufficient in four food groups (American Indian/Alaska Native foods; snacks; finfish and shellfish products; vegetables and vegetable products); it was insufficient in 11 food groups (beverages; meals, entrees, and side dishes; dairy and egg products; restaurant foods; sweets; soups, sauces, and gravies; fruits and fruit juices; baked products; poultry products; lamb, veal, and game products; fast foods); and it was absent in five food groups (beef products; fats and oils; pork products; sausages and luncheon meats; spices and herbs) (Figure 20). High magnesium and magnesium source foods based on the proposed method are given in Tables S17 and S18, respectively.

**Figure 20:**
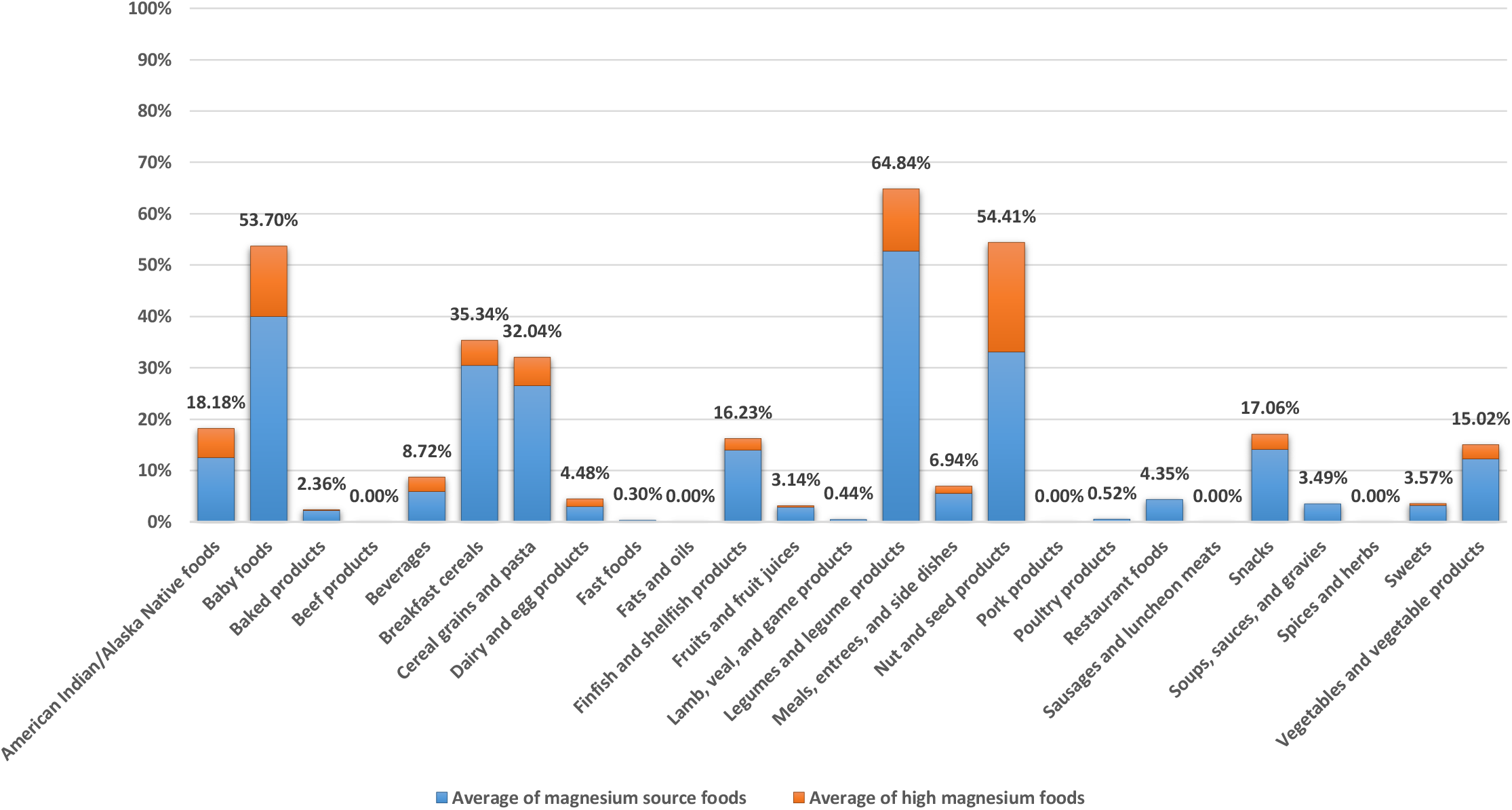
Average of magnesium source and high magnesium foods based on the proposed method in food groups. All magnesium source and high magnesium foods except the magnesium source and high magnesium baby foods are based on the reference energy intake of 2,000 kcal for adults and children aged 4 years and older. The magnesium source and high magnesium baby foods are based on the reference energy intake of 1,000 kcal for children 1 through 3 years of age.

### Manganese source and high manganese foods

Manganese is involved in the formation of bone and in amino acid, lipid, and carbohydrate metabolism (IOM, 2001). Based on the Total Diet Study, grain products contributed 37 percent of dietary manganese, while beverages (tea) and vegetables contributed 20 and 18 percent, respectively, to the adult male diet (Pennington and Young, 1991). According to the proposed method, the average of manganese source and high manganese foods in food groups was as follows: it was excellent in one food group (legumes and legume products); it was almost excellent in one food group (nut and seed products); it was very good in three food groups (meals, entrees, and side dishes; breakfast cereals; cereal grains and pasta); it was almost very good in one food group (snacks); it was almost good in two food groups (vegetables and vegetable products; soups, sauces, and gravies); it was satisfactory in two food groups (baked products; American Indian/Alaska Native foods); it was acceptable in five food groups (fast foods; restaurant foods; baby foods; fruits and fruit juices; beverages); it was almost insufficient in two food groups (finfish and shellfish products; sweets); it was insufficient in seven food groups (sausages and luncheon meats; spices and herbs; poultry products; lamb, veal, and game products; beef products; dairy and egg products; pork products); and it was absent in one food group (fats and oils) (Figure 21). High manganese and manganese source foods based on the proposed method are given in Tables S19 and S20, respectively.

**Figure 21:**
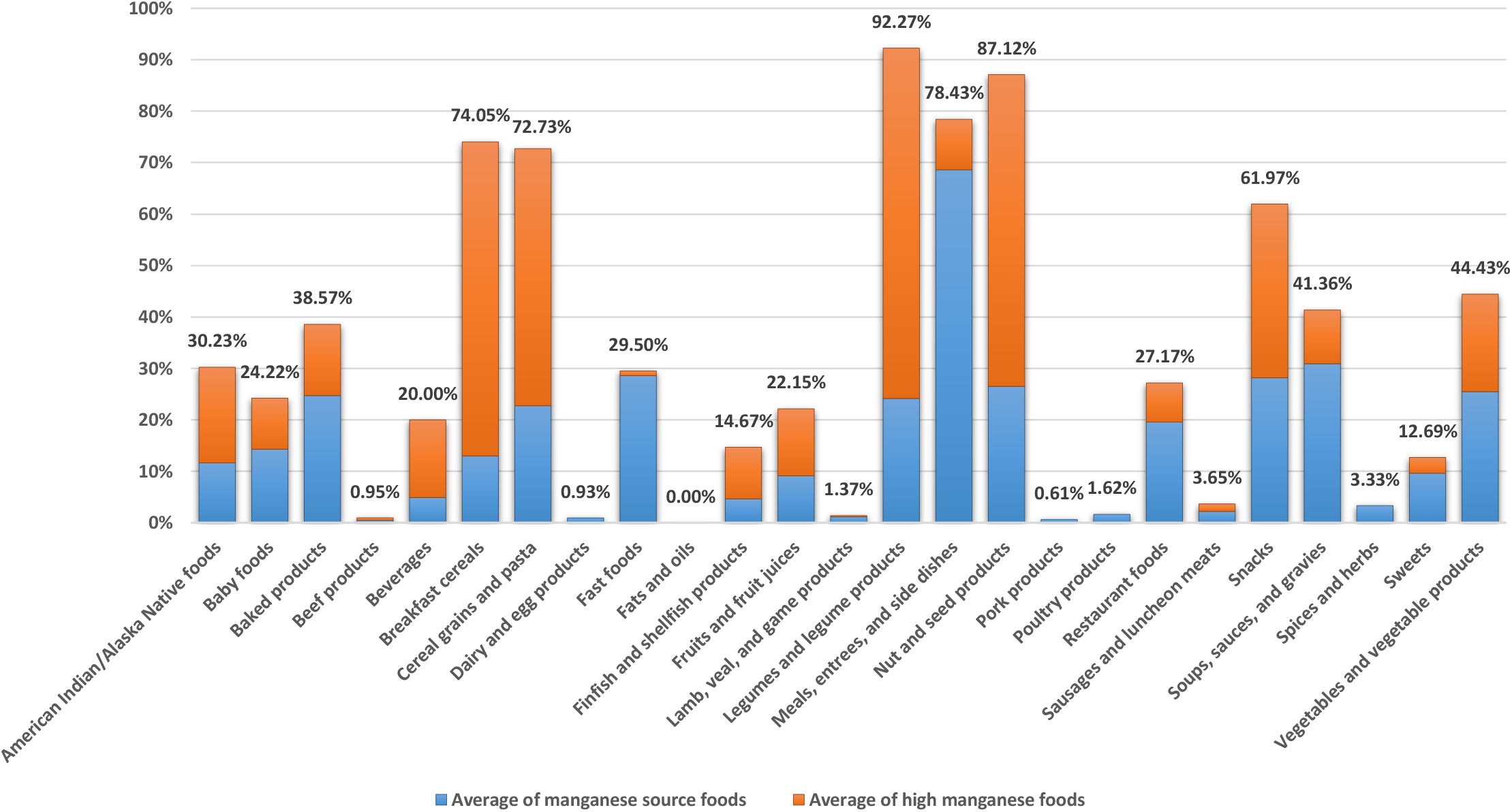
Average of manganese source and high manganese foods based on the proposed method in food groups. All manganese source and high manganese foods except the manganese source and high manganese baby foods are based on the reference energy intake of 2,000 kcal for adults and children aged 4 years and older. The manganese source and high manganese baby foods are based on the reference energy intake of 1,000 kcal for children 1 through 3 years of age.

### Niacin source and high niacin foods

Niacin (also known as vitamin B_3_) functions as a cosubstrate or coenzyme for the transfer of the hydride ion with numerous dehydrogenases (IOM, 1998). Data obtained from the 1995 Continuing Survey of Food Intakes by Individuals (CSFII) indicate that the greatest contribution to the niacin intake of the U.S. adult population comes from mixed dishes high in meat, fish, or poultry; poultry as an entree; enriched and whole-grain breads and bread products; and fortified ready-to-eat cereals (IOM, 1998). According to the proposed method, the average of niacin source and high niacin foods in food groups was as follows: it was excellent in four food groups (poultry products; beef products; lamb, veal, and game products; pork products); it was almost excellent in two food groups (breakfast cereals; meals, entrees, and side dishes); it was very good in two food groups (finfish and shellfish products; fast foods); it was almost very good in two food groups (sausages and luncheon meats; baby foods); it was good in two food groups (cereal grains and pasta; American Indian/Alaska Native foods); it was almost good in one food group (restaurant foods); it was satisfactory in one food group (snacks); it was acceptable in three food groups (baked products; legumes and legume products; nut and seed products); it was almost insufficient in three food groups (beverages; soups, sauces, and gravies; vegetables and vegetable products); it was insufficient in three food groups (fruits and fruit juices; dairy and egg products; sweets); and it was absent in two food groups (fats and oils; spices and herbs) (Figure 22). High niacin and niacin source foods based on the proposed method are given in Tables S21 and S22, respectively.

**Figure 22:**
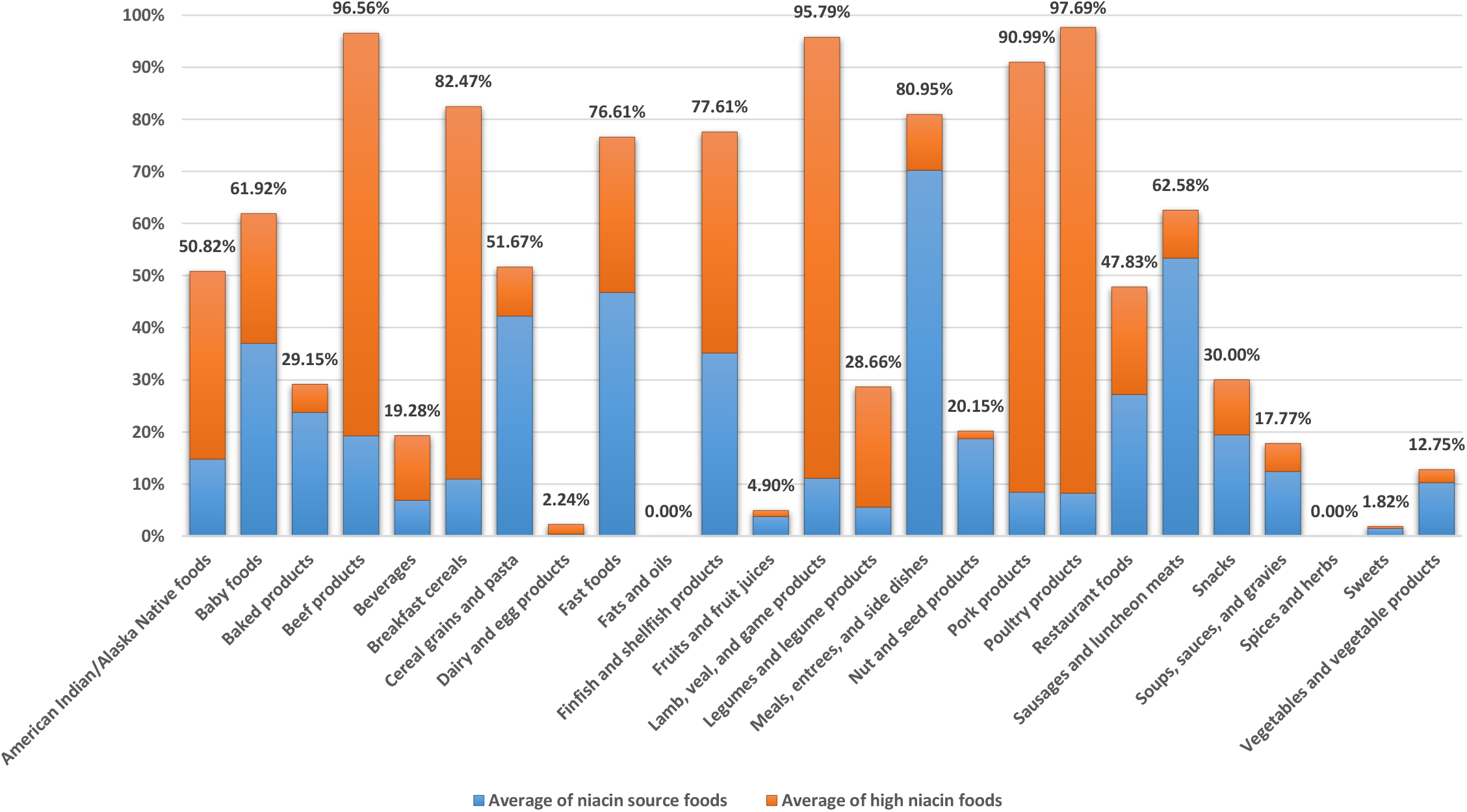
Average of niacin source and high niacin foods based on the proposed method in food groups. All niacin source and high niacin foods except the niacin source and high niacin baby foods are based on the reference energy intake of 2,000 kcal for adults and children aged 4 years and older. The niacin source and high niacin baby foods are based on the reference energy intake of 1,000 kcal for children 1 through 3 years of age.

### Pantothenic acid source and high pantothenic acid foods

Pantothenic acid (also known as vitamin B_5_) functions as a component of coenzyme A and phosphopantetheine, which are involved in fatty acid metabolism (IOM, 1998). Chicken, beef, potatoes, oat cereals, tomato products, liver, kidney, yeast, egg yolk, broccoli, and whole grains are reported to be major sources of pantothenic acid (Plesofsky-Vig, 1996; Walsh *et al*., 1981). According to the proposed method, the average of pantothenic acid source and high pantothenic acid foods in food groups was as follows: it was almost excellent in one food group (poultry products); it was very good in one food group (pork products); it was almost very good in one food group (lamb, veal, and game products); it was good in two food groups (baby foods; finfish and shellfish products); it was almost good in three food groups (beef products; dairy and egg products; American Indian/Alaska Native foods); it was satisfactory in three food groups (fast foods; meals, entrees, and side dishes; restaurant foods); it was acceptable in two food groups (legumes and legume products; beverages); it was almost insufficient in five food groups (breakfast cereals; sausages and luncheon meats; vegetables and vegetable products; cereal grains and pasta; soups, sauces, and gravies); it was insufficient in five food groups (snacks; nut and seed products; fruits and fruit juices; sweets; baked products); and it was absent in two food groups (fats and oils; spices and herbs) (Figure 23). High pantothenic acid and pantothenic acid source foods based on the proposed method are given in Tables S23 and S24, respectively.

**Figure 23:**
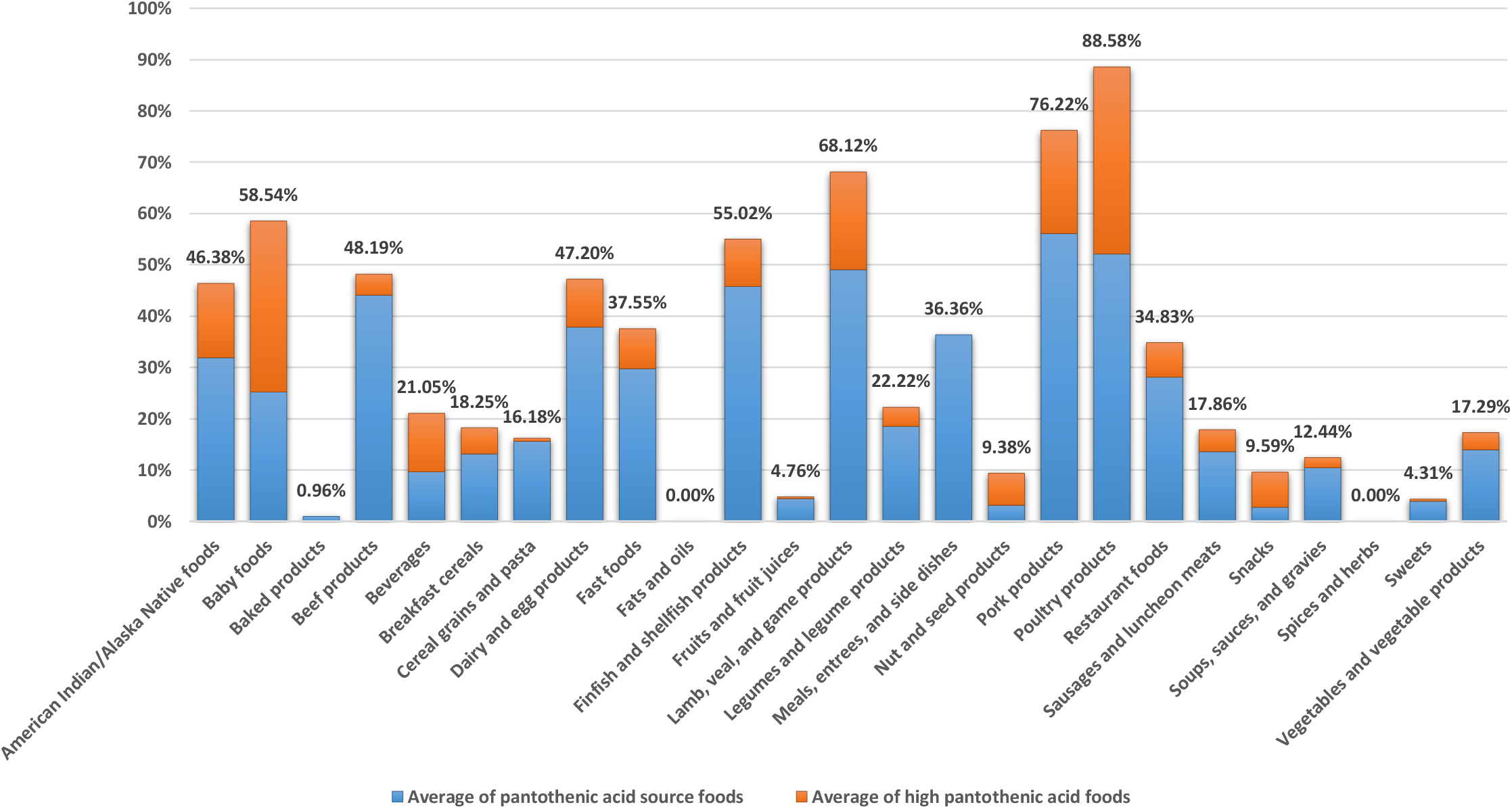
Average of pantothenic acid source and high pantothenic acid foods based on the proposed method in food groups. All pantothenic acid source and high pantothenic acid foods except the pantothenic acid source and high pantothenic acid baby foods are based on the reference energy intake of 2,000 kcal for adults and children aged 4 years and older. The pantothenic acid source and high pantothenic acid baby foods are based on the reference energy intake of 1,000 kcal for children 1 through 3 years of age.

### Phosphorus source and high phosphorus foods

Phosphorus is a major component of bones and teeth (IOM, 2006). Phosphorus helps maintain a normal pH in the body and is involved in metabolic processes (IOM, 2006). Phosphorus is found naturally in many foods in the form of phosphate and as a food additive in the form of various phosphate salts, which are used for nonnutrient functions during food processing, such as moisture retention, smoothness, and binding (IOM, 2006). In the United States, dairy products contribute about 20% of total phosphorus intakes, and bakery products (e.g., breads, tortillas, and sweet bakery products) contribute 10% (Moshfegh *et al*., 2016). According to the proposed method, the average of phosphorus source and high phosphorus foods in food groups was as follows: it was excellent in one food group (finfish and shellfish products); it was almost excellent in four food groups (pork products; beef products; lamb, veal, and game products; poultry products); it was almost very good in two food groups (fast foods; dairy and egg products); it was good in three food groups (restaurant foods; legumes and legume products; American Indian/Alaska Native foods); it was almost good in three food groups (nut and seed products; baby foods; meals, entrees, and side dishes); it was satisfactory in one food group (breakfast cereals); it was acceptable in one food group (cereal grains and pasta); it was almost insufficient in four food groups (beverages; soups, sauces, and gravies; baked products; sausages and luncheon meats); it was insufficient in four food groups (sweets; snacks; vegetables and vegetable products; fruits and fruit juices); and it was absent in two food groups (fats and oils; spices and herbs) (Figure 24). High phosphorus and phosphorus source foods based on the proposed method are given in Tables S25 and S26, respectively.

**Figure 24:**
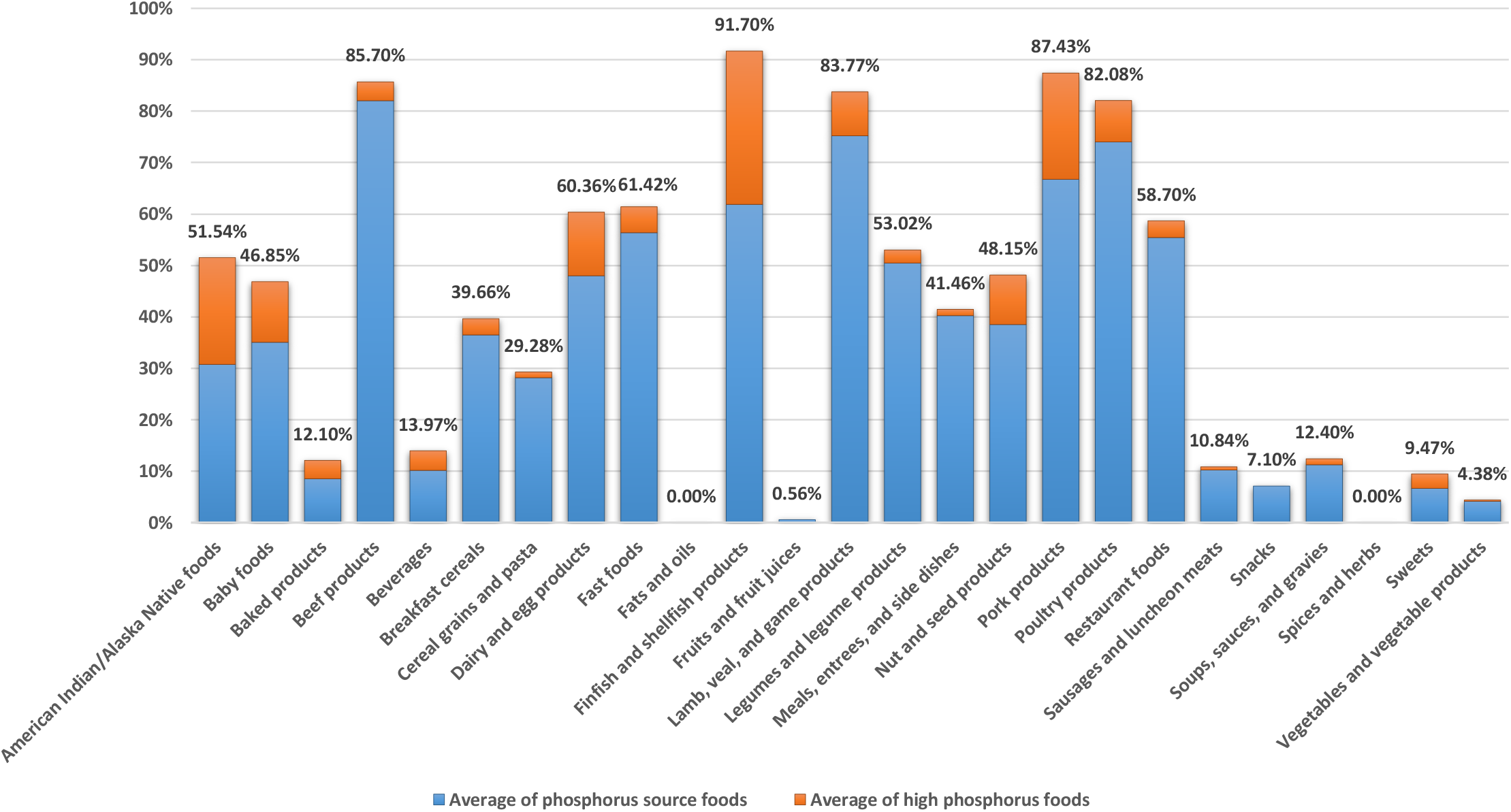
Average of phosphorus source and high phosphorus foods based on the proposed method in food groups. All phosphorus source and high phosphorus foods except the phosphorus source and high phosphorus baby foods are based on the reference energy intake of 2,000 kcal for adults and children aged 4 years and older. The phosphorus source and high phosphorus baby foods are based on the reference energy intake of 1,000 kcal for children 1 through 3 years of age.

### Potassium source and high potassium foods

Potassium, the major intracellular cation in the body, is required for normal cellular function (IOM, 2005a), although the mineral is found in both the intracellular and the extracellular fluids (IOM, 2006). Relatively small changes in the concentration of extracellular potassium greatly affect the extracellular/intracellular potassium ratio and thereby affect nerve transmission, muscle contraction, and vascular tone (Appel, 2012). Milk, coffee, tea, other nonalcoholic beverages, and potatoes are the top sources of potassium in the diets of U.S. adults (O’Neil *et al*., 2012). Among children in the United States, milk, fruit juice, potatoes, and fruit are the top sources (Keast *et al*., 2013). According to the proposed method, the average of potassium source and high potassium foods in food groups was as follows: it was almost insufficient in three food groups (legumes and legume products; soups, sauces, and gravies; vegetables and vegetable products); it was insufficient in 17 food groups (finfish and shellfish products; American Indian/Alaska Native foods; pork products; meals, entrees, and side dishes; fruits and fruit juices; beverages; dairy and egg products; restaurant foods; nut and seed products; breakfast cereals; baby foods; snacks; sausages and luncheon meats; fast foods; poultry products; lamb, veal, and game products; beef products); and it was absent in five food groups (baked products; cereal grains and pasta; fats and oils; spices and herbs; sweets) (Figure 25). High potassium and potassium source foods based on the proposed method are given in Tables S27 and S28, respectively.

**Figure 25:**
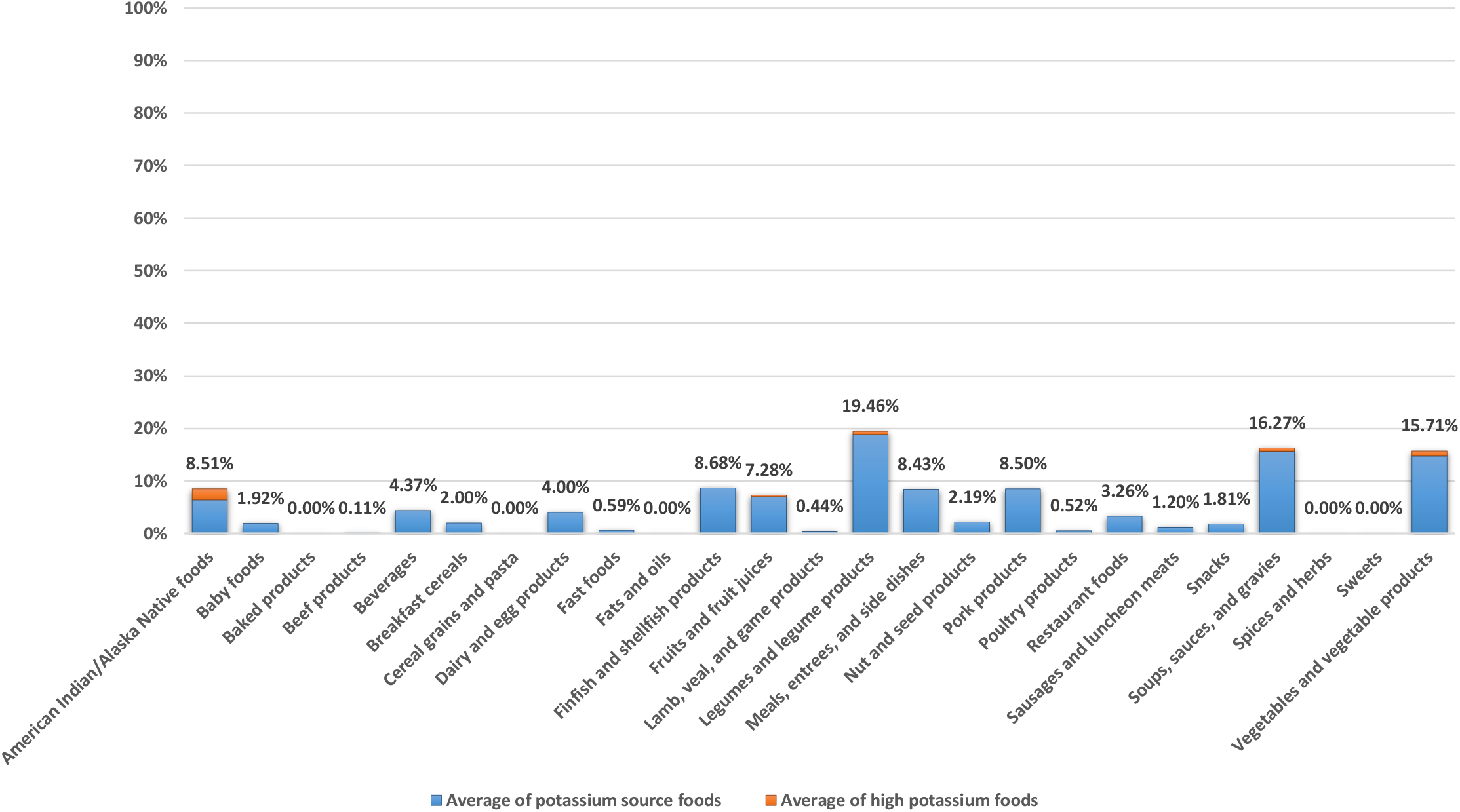
Average of potassium source and high potassium foods based on the proposed method in food groups. All potassium source and high potassium foods except the potassium source and high potassium baby foods are based on the reference energy intake of 2,000 kcal for adults and children aged 4 years and older. The potassium source and high potassium baby foods are based on the reference energy intake of 1,000 kcal for children 1 through 3 years of age.

### Protein source and high protein foods

Proteins are the major structural components of all cells of the body, and amino acids are the basic building blocks of proteins (Antonio *et al*., 2008). Proteins also function as enzymes, in membranes, as transport carriers, and as hormones; and their component amino acids serve as precursors for nucleic acids, hormones, vitamins, and other important molecules (IOM, 2005b). Protein from animal sources such as meat, poultry, fish, eggs, milk, cheese, and yogurt provide all nine indispensable amino acids, and for this reason are referred to as “complete proteins” (IOM, 2005b). Proteins from plants, legumes, grains, nuts, seeds, and vegetables tend to be deficient in one or more of the indispensable amino acids and are called “incomplete proteins” (IOM, 2005b). According to the proposed method, the average of protein source and high protein foods in food groups was as follows: it was excellent in seven food groups (poultry products; beef products; finfish and shellfish products; lamb, veal, and game products; sausages and luncheon meats; pork products; meals, entrees, and side dishes); it was almost excellent in two food groups (legumes and legume products; fast foods); it was very good in one food group (restaurant foods); it was almost very good in three food groups (dairy and egg products; baby foods; American Indian/Alaska Native foods); it was almost good in one food group (nut and seed products); it was satisfactory in one food group (cereal grains and pasta); it was acceptable in two food groups (soups, sauces, and gravies; breakfast cereals); it was almost insufficient in one food group (snacks); it was insufficient in five food groups (beverages; baked products; vegetables and vegetable products; sweets; fruits and fruit juices); and it was absent in two food groups (fats and oils; spices and herbs) (Figure 26). High protein and protein source foods based on the proposed method are given in Tables S29 and S30, respectively.

**Figure 26:**
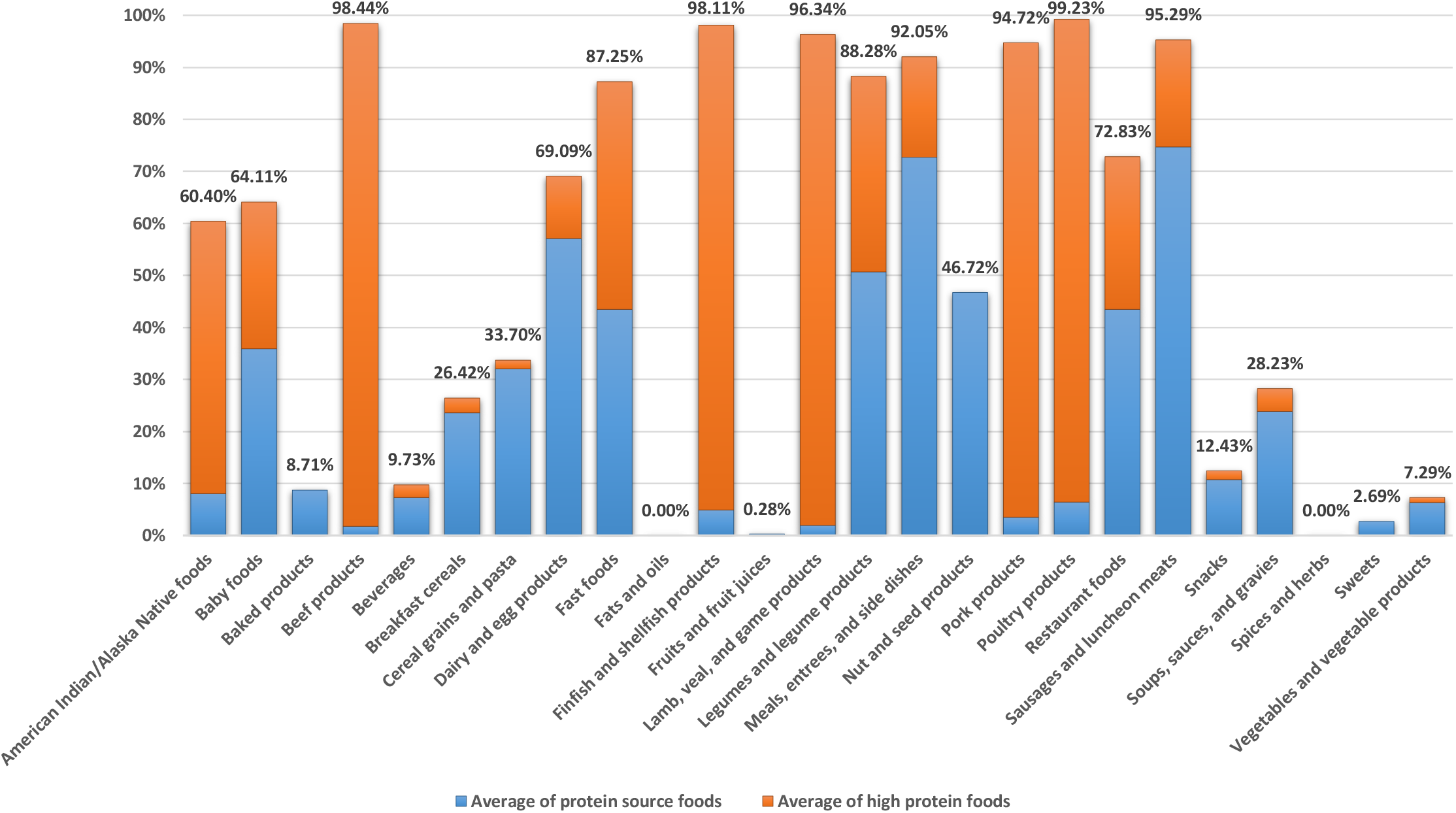
Average of protein source and high protein foods based on the proposed method in food groups. All protein source and high protein foods except the protein source and high protein baby foods are based on the reference energy intake of 2,000 kcal for adults and children aged 4 years and older. The protein source and high protein baby foods are based on the reference energy intake of 1,000 kcal for children 1 through 3 years of age.

### Riboflavin source and high riboflavin foods

Riboflavin (also known as vitamin B_2_) is required for the flavoenzymes of the respiratory chain (Depeint *et al*., 2006). Data obtained from the 1995 Continuing Survey of Food Intakes by Individuals (CSFII) indicate that the greatest contribution to the riboflavin intake of the U.S. adult population comes from milk and milk drinks followed by bread products and fortified cereals (IOM, 1998). Other sources of riboflavin are organ meats (IOM, 1998). According to the proposed method, the average of riboflavin source and high riboflavin foods in food groups was as follows: it was almost excellent in two food groups (pork products; lamb, veal, and game products); it was very good in four food groups (breakfast cereals; poultry products; beef products; fast foods); it was almost very good in two food groups (baby foods; meals, entrees, and side dishes); it was good in one food group (dairy and egg products); it was almost good in one food group (American Indian/Alaska Native foods); it was satisfactory in four food groups (restaurant foods; finfish and shellfish products; legumes and legume products; baked products); it was acceptable in three food groups (beverages; sausages and luncheon meats; cereal grains and pasta); it was almost insufficient in five food groups (snacks; sweets; vegetables and vegetable products; soups, sauces, and gravies; nut and seed products); it was insufficient in one food group (fruits and fruit juices); and it was absent in two food groups (fats and oils; spices and herbs) (Figure 27). High riboflavin and riboflavin source foods based on the proposed method are given in Tables S31 and S32, respectively.

**Figure 27:**
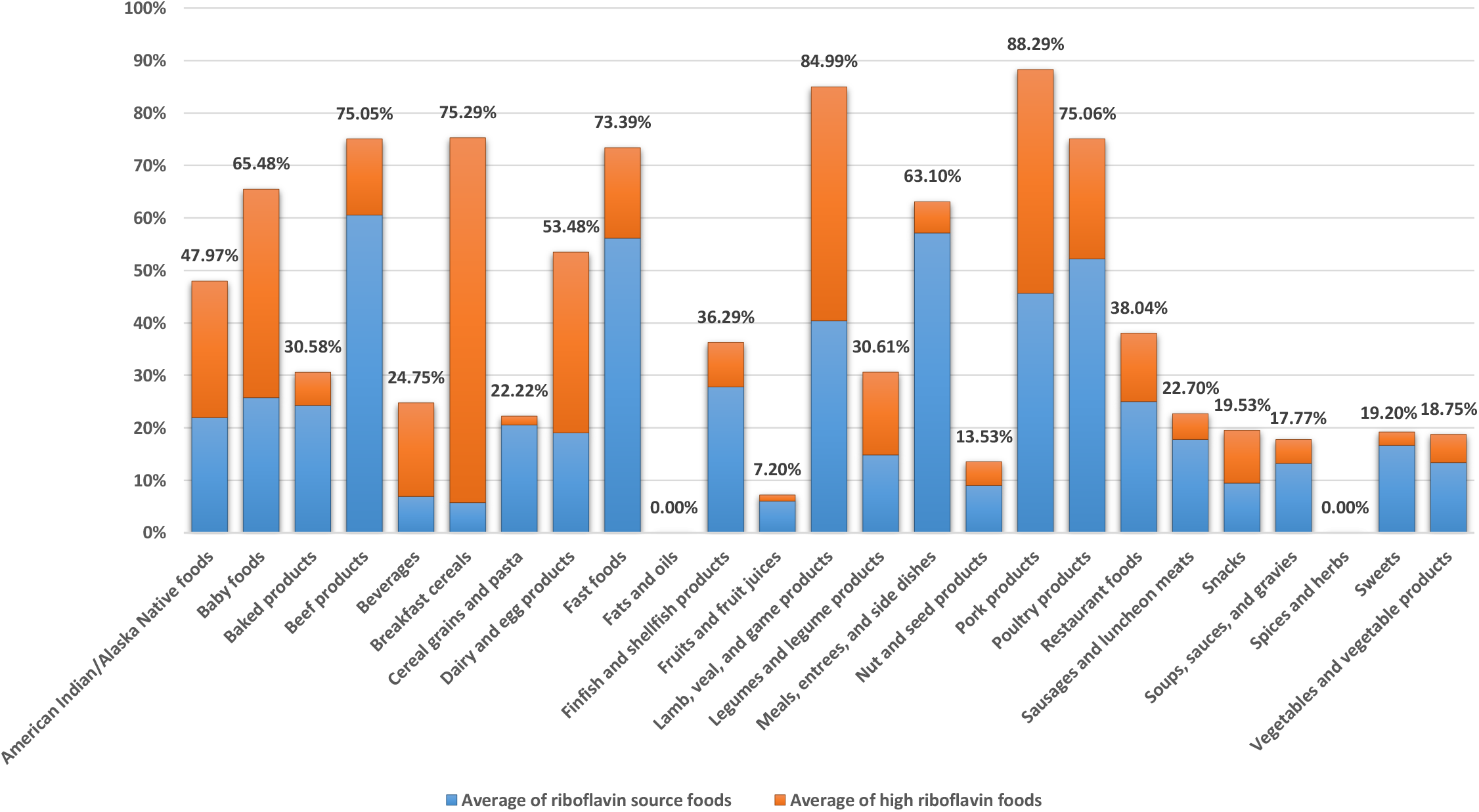
Average of riboflavin source and high riboflavin foods based on the proposed method in food groups. All riboflavin source and high riboflavin foods except the riboflavin source and high riboflavin baby foods are based on the reference energy intake of 2,000 kcal for adults and children aged 4 years and older. The riboflavin source and high riboflavin baby foods are based on the reference energy intake of 1,000 kcal for children 1 through 3 years of age.

### Saturated fat free and low saturated fat foods

Saturated fatty acids are synthesized by the body to provide an adequate level needed for their physiological and structural functions (IOM, 2005b). Sources of saturated fatty acids tend to be foods of animal sources, including whole milk, cream, butter, cheese, and fatty meats such as pork and beef (USDA and HHS, 2000). Some coconut oil, palm kernel oil, and palm oil are very high in saturated fat; they often are used in bakery goods, processed foods, popcorn oils, and nondairy creamers (Grundy, 1993). There is a positive linear trend between total saturated fatty acid intake and total and low density lipoprotein (LDL) cholesterol concentration and increased risk of coronary heart disease (Caggiula and Mustad, 1997; IOM, 2005b; Siri-Tarino *et al*., 2010). According to the proposed method, the average of saturated fat free foods in food groups was as follows: it was almost excellent in two food groups (fruits and fruit juices; beverages); it was very good in one food group (vegetables and vegetable products); it was satisfactory in two food groups (cereal grains and pasta; spices and herbs); it was acceptable in four food groups (sweets; baby foods; legumes and legume products; soups, sauces, and gravies); it was almost insufficient in two food groups (nut and seed products; breakfast cereals); it was insufficient in nine food groups (dairy and egg products; finfish and shellfish products; baked products; American Indian/Alaska Native foods; snacks; fats and oils; sausages and luncheon meats; meals, entrees, and side dishes; fast foods); and it was absent in five food groups (beef products; lamb, veal, and game products; pork products; poultry products; restaurant foods). Also, the average of low saturated fat foods in food groups was as follows: it was excellent in four food groups (cereal grains and pasta; fruits and fruit juices; vegetables and vegetable products; breakfast cereals); it was almost excellent in two food groups (beverages; spices and herbs); it was very good in one food group (legumes and legume products); it was almost very good in two food groups (finfish and shellfish products; American Indian/Alaska Native foods); it was good in one food group (soups, sauces, and gravies); it was almost good in two food groups (snacks; baby foods); it was satisfactory in two food groups (sweets; baked products); it was acceptable in three food groups (nut and seed products; dairy and egg products; meals, entrees, and side dishes); it was almost insufficient in three food groups (poultry products; fats and oils; sausages and luncheon meats); and it was insufficient in five food groups (lamb, veal, and game products; restaurant foods; pork products; fast foods; beef products) (Figure 28). Saturated fat free and low saturated fat foods based on the proposed method are given in Tables S33 and S34, respectively.

**Figure 28:**
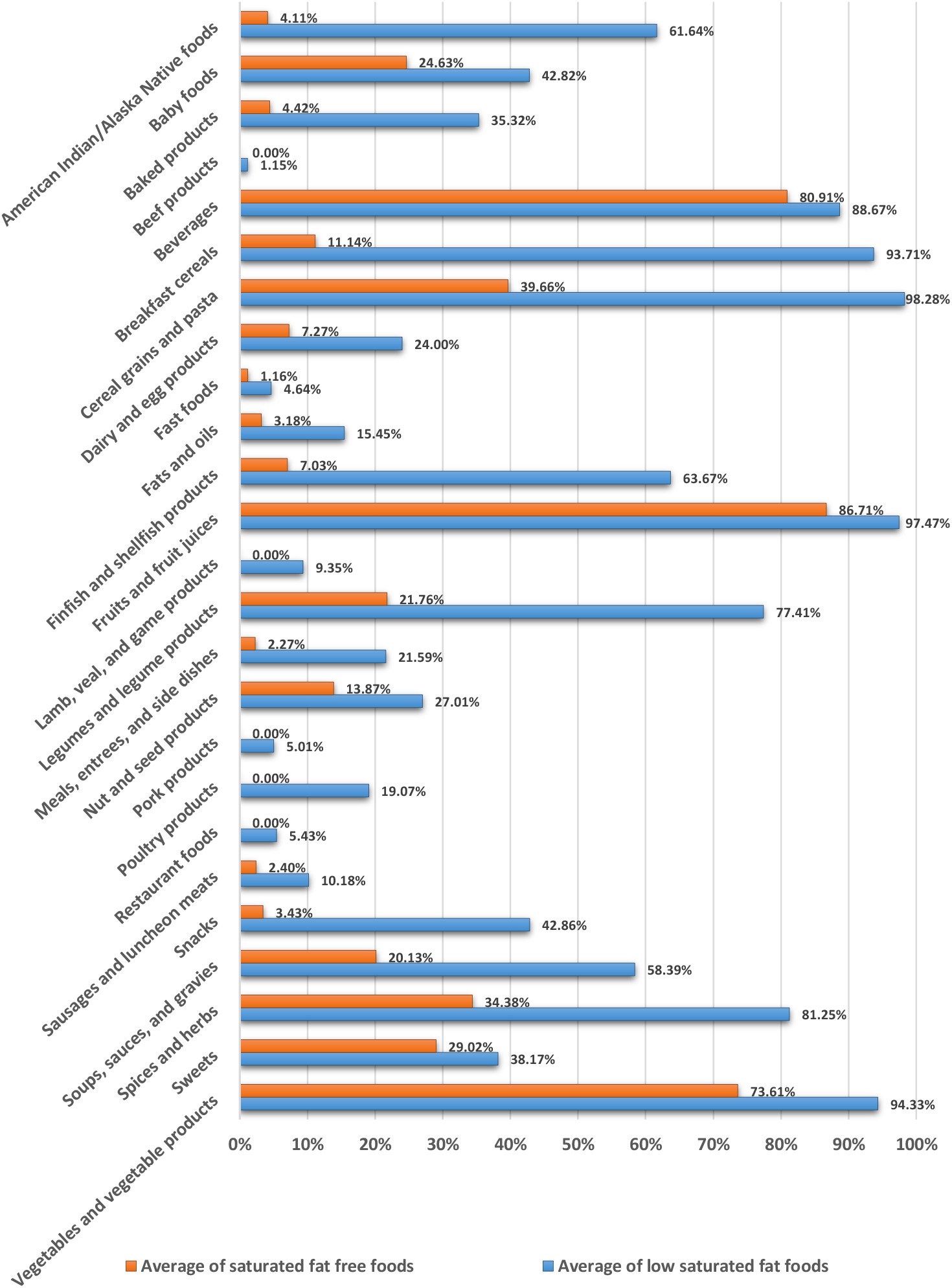
Averages of saturated fat free and low saturated fat foods based on the proposed method in food groups. All saturated fat free and low saturated fat foods except the saturated fat free and low saturated fat baby foods are based on the reference energy intake of 2,000 kcal for adults and children aged 4 years and older. The saturated fat free and low saturated fat baby foods are based on the reference energy intake of 1,000 kcal for children 1 through 3 years of age.

### Selenium source and high selenium foods

Selenium functions through selenoproteins, several of which are oxidant defense enzymes (IOM, 2000). The selenium content of food varies depending on the selenium content of the soil where the animal was raised or the plant was grown (IOM, 2000). The major food sources of selenium in the American diet are breads, grains, meat, poultry, fish, and eggs (Chun *et al*., 2010). According to the proposed method, the average of selenium source and high selenium foods in food groups was as follows: it was excellent in six food groups (finfish and shellfish products; poultry products; meals, entrees, and side dishes; pork products; beef products; fast foods); it was almost excellent in one food group (sausages and luncheon meats); it was very good in three food groups (lamb, veal, and game products; restaurant foods; American Indian/Alaska Native foods); it was almost very good in one food group (cereal grains and pasta); it was good in three food groups (breakfast cereals; baby foods; baked products); it was almost good in one food group (dairy and egg products); it was satisfactory in one food group (nut and seed products); it was acceptable in three food groups (soups, sauces, and gravies; legumes and legume products; snacks); it was insufficient in three food groups (beverages; vegetables and vegetable products; sweets); and it was absent in three food groups (fats and oils; fruits and fruit juices; spices and herbs) (Figure 29). High selenium and selenium source foods based on the proposed method are given in Tables S35 and S36, respectively.

**Figure 29:**
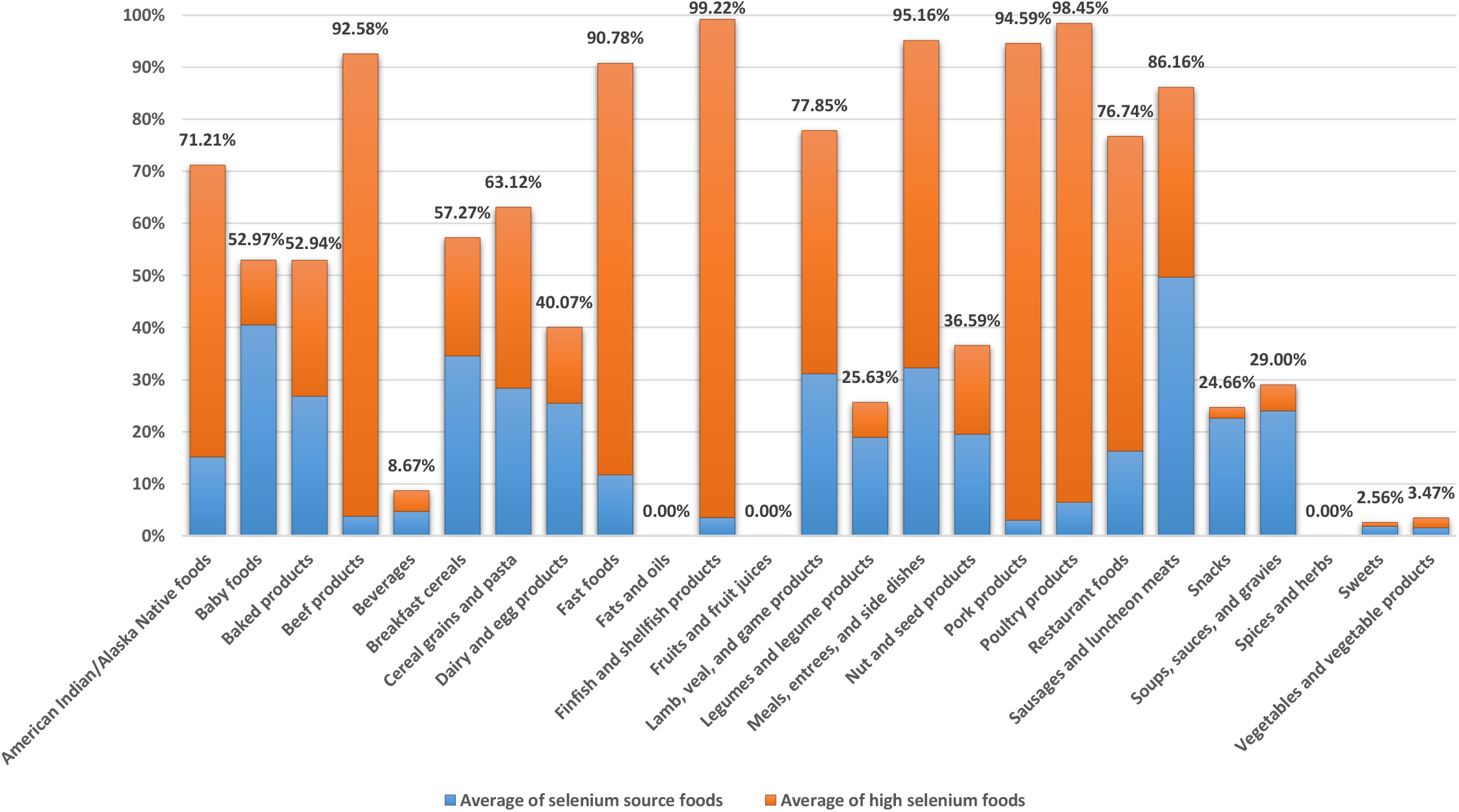
Average of selenium source and high selenium foods based on the proposed method in food groups. All selenium source and high selenium foods except the selenium source and high selenium baby foods are based on the reference energy intake of 2,000 kcal for adults and children aged 4 years and older. The selenium source and high selenium baby foods are based on the reference energy intake of 1,000 kcal for children 1 through 3 years of age.

### Sodium free, very low sodium, and low sodium foods

Sodium is the principal cation of the extracellular fluid and functions as the osmotic determinant in regulating extracellular fluid volume and thus plasma volume (IOM, 2005a). Sodium chloride (salt) is the primary form of sodium in the diet (IOM, 2005a). Only about 12 percent of the total sodium chloride consumed is naturally occurring (Mattes and Donnelly, 1991). The major adverse effect of increased sodium chloride intake is elevated blood pressure, which has been shown to be an etiologically related risk factor for cardiovascular and renal diseases (IOM, 2005a). According to the proposed method, the average of sodium free foods in food groups was as follows: it was almost excellent in one food group (fruits and fruit juices); it was very good in one food group (cereal grains and pasta); it was almost very good in one food group (nut and seed products); it was good in one food group (fats and oils); it was almost good in one food group (spices and herbs); it was satisfactory in one food group (beverages); it was acceptable in five food groups (vegetables and vegetable products; legumes and legume products; baby foods; American Indian/Alaska Native foods; sweets); it was almost insufficient in two food groups (breakfast cereals; snacks); it was insufficient in eight food groups (dairy and egg products; meals, entrees, and side dishes; baked products; soups, sauces, and gravies; sausages and luncheon meats; pork products; fast foods; beef products); and it was absent in four food groups (finfish and shellfish products; lamb, veal, and game products; poultry products; restaurant foods). Also, the average of very low sodium foods in food groups was as follows: it was excellent in one food group (fruits and fruit juices); it was almost excellent in one food group (cereal grains and pasta); it was very good in two food groups (nut and seed products; spices and herbs); it was almost very good in two food groups (beverages; baby foods); it was good in five food groups (sweets; American Indian/Alaska Native foods; fats and oils; vegetables and vegetable products; beef products); it was satisfactory in one food group (legumes and legume products); it was acceptable in five food groups (lamb, veal, and game products; pork products; finfish and shellfish products; dairy and egg products; breakfast cereals); it was almost insufficient in two food groups (snacks; poultry products); and it was insufficient in six food groups (baked products; restaurant foods; soups, sauces, and gravies; sausages and luncheon meats; meals, entrees, and side dishes; fast foods). In addition, the average of low sodium foods in food groups was as follows: it was excellent in four food groups (beef products; fruits and fruit juices; lamb, veal, and game products; baby foods); it was almost excellent in four food groups (nut and seed products; cereal grains and pasta; sweets; spices and herbs); it was very good in two food groups (beverages; American Indian/Alaska Native foods); it was almost very good in three food groups (poultry products; finfish and shellfish products; vegetables and vegetable products); it was good in two food groups (pork products; fats and oils); it was almost good in three food groups (snacks; dairy and egg products; legumes and legume products); it was acceptable in two food groups (breakfast cereals; baked products); and it was insufficient in five food groups (soups, sauces, and gravies; restaurant foods; fast foods; sausages and luncheon meats; meals, entrees, and side dishes) (Figure 30). Sodium free, very low sodium, and low sodium foods based on the proposed method are given in Tables S37, S38, and S39, respectively.

**Figure 30:**
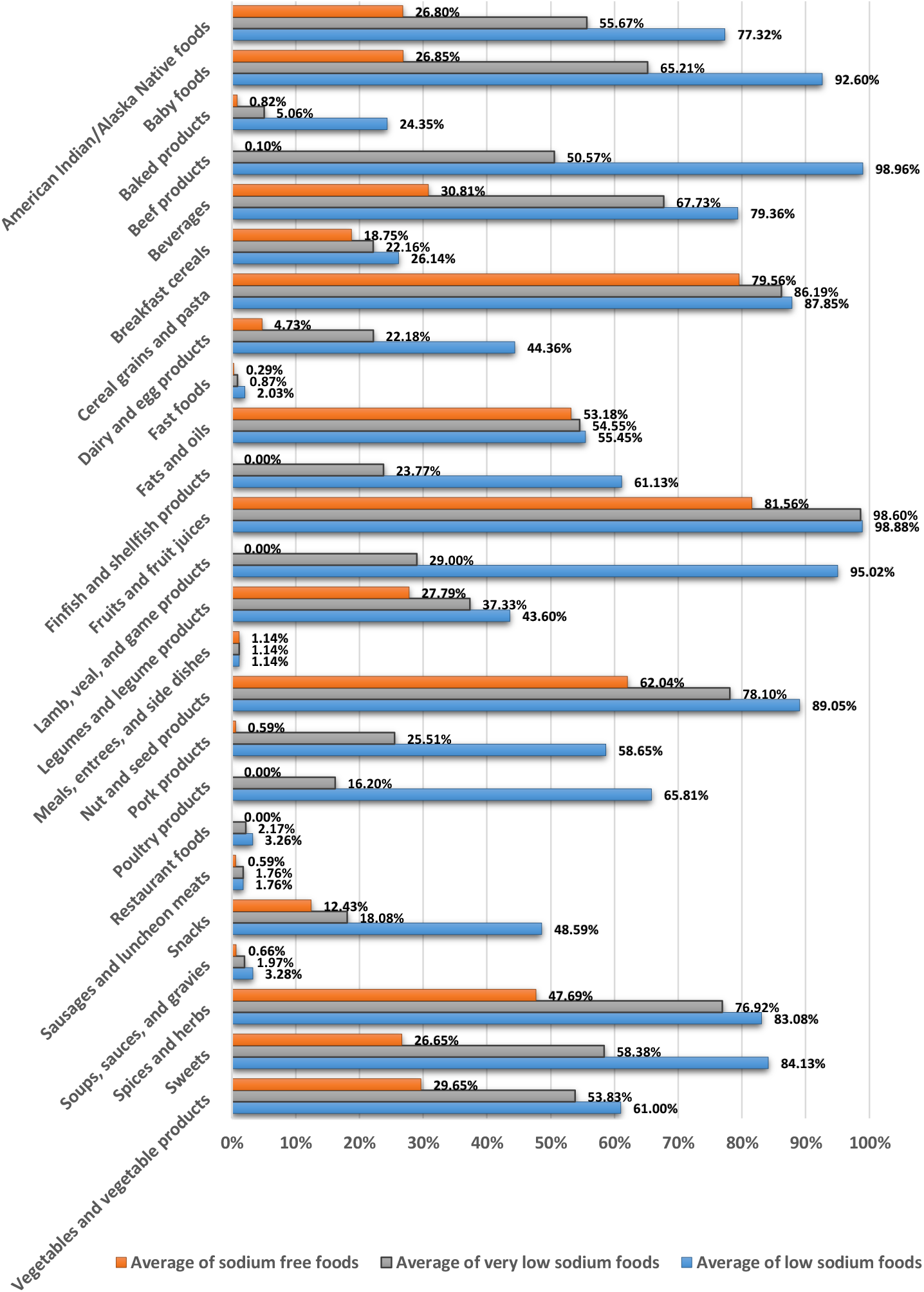
Averages of sodium free, very low sodium, and low sodium foods based on the proposed method in food groups. All sodium free, very low sodium, and low sodium foods except the sodium free, very low sodium, and low sodium baby foods are based on the reference energy intake of 2,000 kcal for adults and children aged 4 years and older. The sodium free, very low sodium, and low sodium baby foods are based on the reference energy intake of 1,000 kcal for children 1 through 3 years of age.

### Thiamin source and high thiamin foods

Thiamin (also known as vitamin B_1_ and aneurin) functions as a coenzyme in the metabolism of carbohydrates and branched-chain amino acids (IOM, 1998). Data obtained from the 1995 Continuing Survey of Food Intakes by Individuals (CSFII) indicate that the greatest contribution to thiamin intake of the U.S. adult population comes from the following enriched, fortified, or whole-grain products: bread and bread products, mixed foods whose main ingredient is grain, and ready-to-eat cereals (IOM, 1998). Other sources include pork and ham products and cereals and meat substitutes fortified with vitamins (IOM, 1998). According to the proposed method, the average of thiamin source and high thiamin foods in food groups was as follows: it was excellent in one food group (breakfast cereals); it was almost excellent in one food group (pork products); it was very good in one food group (meals, entrees, and side dishes); it was almost very good in one food group (fast foods); it was good in three food groups (legumes and legume products; cereal grains and pasta; baby foods); it was almost good in two food groups (baked products; sausages and luncheon meats); it was satisfactory in one food group (nut and seed products); it was acceptable in four food groups (American Indian/Alaska Native foods; snacks; lamb, veal, and game products; finfish and shellfish products); it was almost insufficient in five food groups (restaurant foods; vegetables and vegetable products; poultry products; beverages; fruits and fruit juices); it was insufficient in five food groups (soups, sauces, and gravies; dairy and egg products; beef products; sweets; fats and oils); and it was absent in one food group (spices and herbs) (Figure 31). High thiamin and thiamin source foods based on the proposed method are given in Tables S40 and S41, respectively.

**Figure 31:**
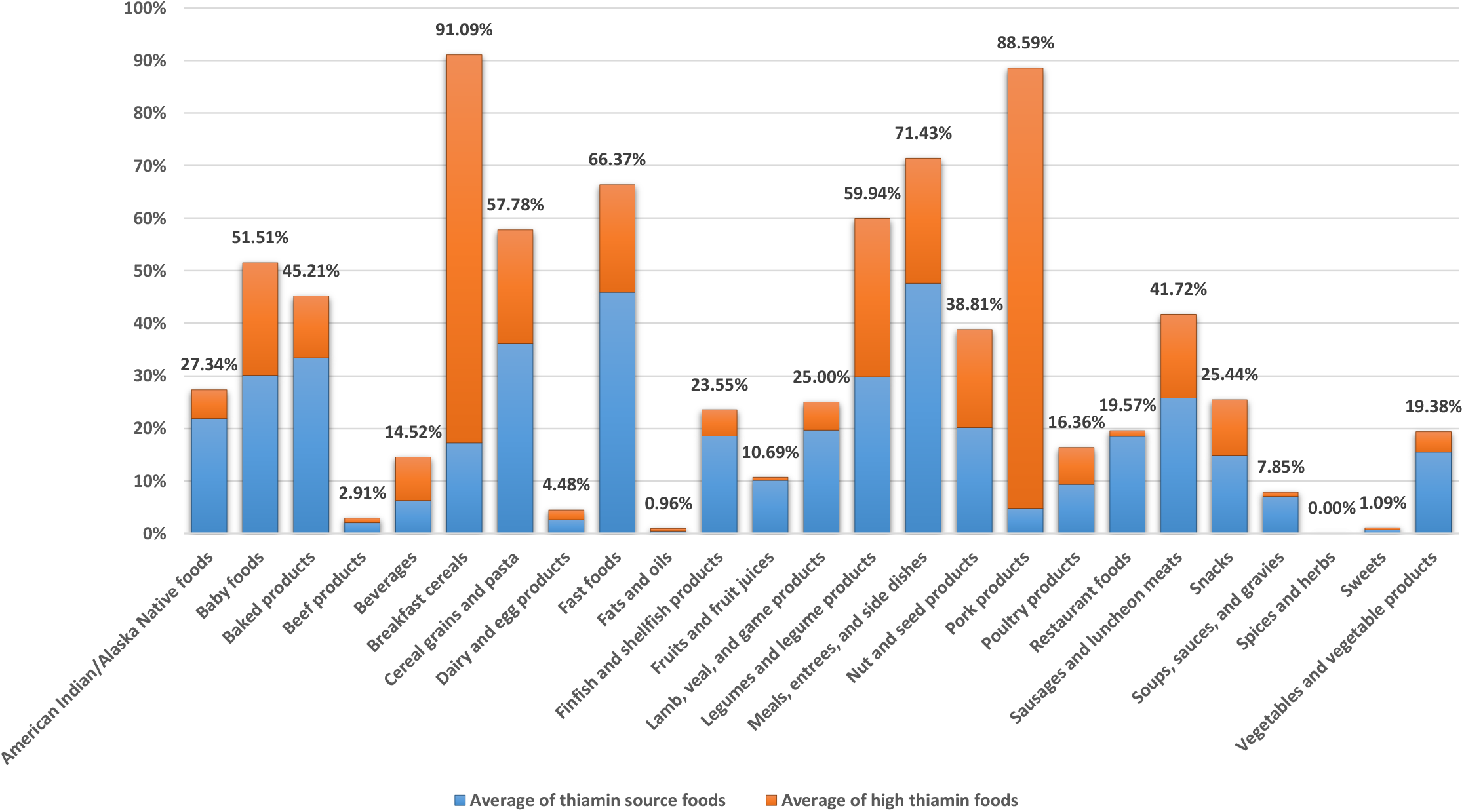
Average of thiamin source and high thiamin foods based on the proposed method in food groups. All thiamin source and high thiamin foods except the thiamin source and high thiamin baby foods are based on the reference energy intake of 2,000 kcal for adults and children aged 4 years and older. The thiamin source and high thiamin baby foods are based on the reference energy intake of 1,000 kcal for children 1 through 3 years of age.

### Total fat free and low total fat foods

Fat is a major source of fuel energy for the body and aids in the absorption of fat-soluble vitamins (vitamins A, D, E, and K) and carotenoids (IOM, 2005b). Fatty acids function in cell signaling and alter the expression of specific genes involved in lipid and carbohydrate metabolism (IOM, 2006). The principal foods that contribute to fat intake are butter, margarine, vegetable oils, visible fat on meat and poultry products, whole milk, egg yolks, nuts, and baked goods (e.g., cookies, doughnuts, and cakes) (IOM, 2005b). High-fat diets in excess of energy needs can cause obesity (IOM, 2006). Several studies have shown associations between high-fat intakes and an increased risk of coronary heart disease, cancer, and insulin resistance, however, the type of fatty acid consumed is very important in defining these associations (IOM, 2006). According to the proposed method, the average of total fat free foods in food groups was as follows: it was very good in two food groups (beverages; fruits and fruit juices); it was almost very good in one food group (vegetables and vegetable products); it was satisfactory in two food groups (sweets; spices and herbs); it was acceptable in one food group (cereal grains and pasta); it was almost insufficient in five food groups (baby foods; dairy and egg products; soups, sauces, and gravies; American Indian/Alaska Native foods; legumes and legume products); it was insufficient in eight food groups (nut and seed products; breakfast cereals; snacks; fats and oils; finfish and shellfish products; sausages and luncheon meats; baked products; fast foods); and it was absent in six food groups (beef products; lamb, veal, and game products; meals, entrees, and side dishes; pork products; poultry products; restaurant foods). Also, the average of low total fat foods in food groups was as follows: it was excellent in four food groups (cereal grains and pasta; fruits and fruit juices; vegetables and vegetable products; beverages); it was almost excellent in one food group (breakfast cereals); it was almost very good in two food groups (spices and herbs; soups, sauces, and gravies); it was good in four food groups (American Indian/Alaska Native foods; legumes and legume products; finfish and shellfish products; sweets); it was almost good in one food group (baby foods); it was satisfactory in two food groups (snacks; dairy and egg products); it was acceptable in three food groups (baked products; nut and seed products; poultry products); it was almost insufficient in four food groups (meals, entrees, and side dishes; lamb, veal, and game products; pork products; sausages and luncheon meats); and it was insufficient in four food groups (beef products; fats and oils; fast foods; restaurant foods) (Figure 32). Total fat free and low total fat foods based on the proposed method are given in Tables S42 and S43, respectively.

**Figure 32:**
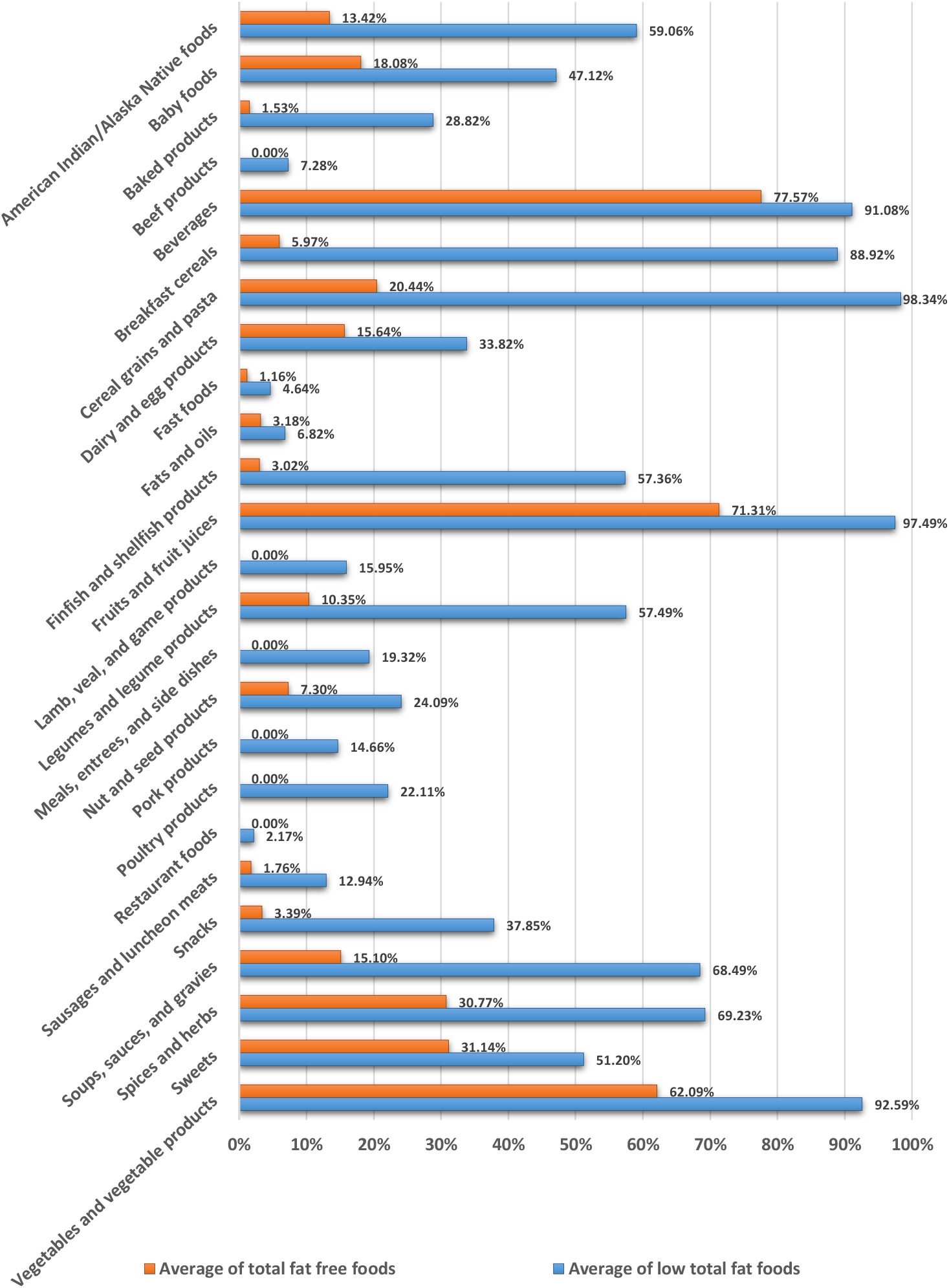
Averages of total fat free and low total fat foods based on the proposed method in food groups. All total fat free and low total fat foods except the total fat free and low total fat baby foods are based on the reference energy intake of 2,000 kcal for adults and children aged 4 years and older. The total fat free and low total fat baby foods are based on the reference energy intake of 1,000 kcal for children 1 through 3 years of a.

### Vitamin A source and high vitamin A foods

Vitamin A is a fat-soluble vitamin that is important for normal vision, gene expression, reproduction, embryonic development, growth, and immune function (IOM, 2001). Preformed vitamin A (retinol) is naturally found in animal-based foods, whereas dietary carotenoids (provitamin A carotenoids), which are converted to vitamin A in the body, are present in oils, fruits, and vegetables (IOM, 2006). Common dietary sources of preformed vitamin A in the United States and Canada include liver, dairy products, and fish (IOM, 2001). According to data collected from the 1994– 1996 Continuing Survey of Food Intakes by Individuals (CFSII), the major contributors of vitamin A from foods were grains and vegetables (approximately 55 percent), followed by dairy and meat products (approximately 30 percent) (IOM, 2001). According to the proposed method, the average of vitamin A source and high vitamin A foods in food groups was as follows: it was good in two food groups (breakfast cereals; baby foods); it was acceptable in three food groups (vegetables and vegetable products; dairy and egg products; American Indian/Alaska Native foods); it was almost insufficient in five food groups (baked products; fats and oils; soups, sauces, and gravies; beverages; finfish and shellfish products); it was insufficient in 12 food groups (snacks; poultry products; meals, entrees, and side dishes; fruits and fruit juices; sausages and luncheon meats; restaurant foods; fast foods; sweets; legumes and legume products; lamb, veal, and game products; beef products; pork products); and it was absent in three food groups (cereal grains and pasta; nut and seed products; spices and herbs) (Figure 33). High vitamin A and vitamin A source foods based on the proposed method are given in Tables S44 and S45, respectively.

**Figure 33:**
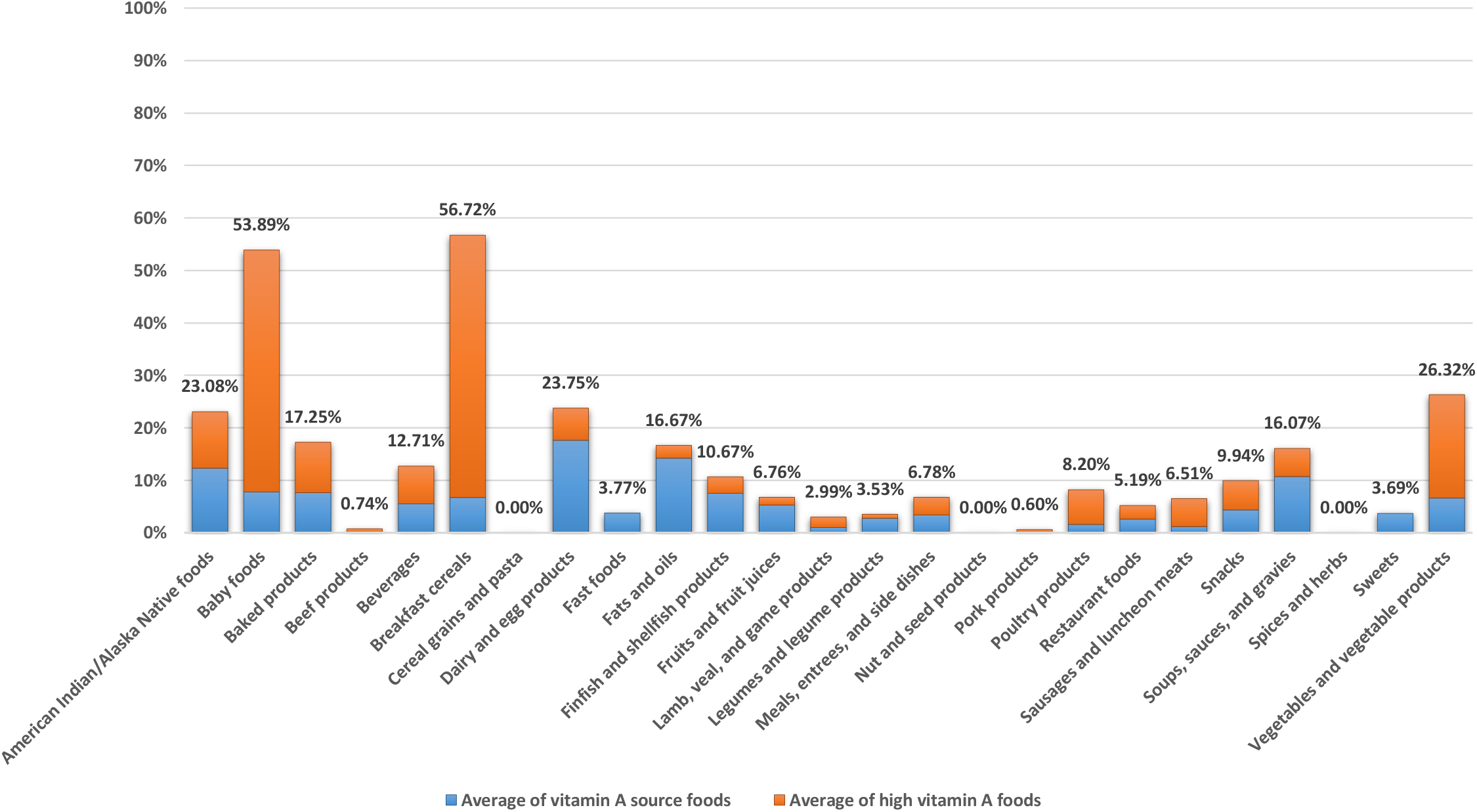
Average of vitamin A source and high vitamin A foods based on the proposed method in food groups. All vitamin A source and high vitamin A foods except the vitamin A source and high vitamin A baby foods are based on the reference energy intake of 2,000 kcal for adults and children aged 4 years and older. The vitamin A source and high vitamin A baby foods are based on the reference energy intake of 1,000 kcal for children 1 through 3 years of age.

### Vitamin B_6_ source and high vitamin B_6_ foods

Vitamin B_6_ (pyridoxine and related compounds) functions as a coenzyme in the metabolism of amino acids, glycogen, and sphingoid bases (IOM, 1998). Data obtained from the 1995 Continuing Survey of Food Intakes by Individuals (CSFII) indicates that the greatest contribution to vitamin B_6_ intake of the U.S. adult population comes from fortified, ready-to-eat cereals; mixed foods (including sandwiches) with meat, fish, or poultry as the main ingredient; white potatoes and other starchy vegetables; and noncitrus fruits (IOM, 1998). Especially rich sources are highly fortified cereals; beef liver and other organ meats; and highly fortified, soy-based meat substitutes (IOM, 1998). According to the proposed method, the average of vitamin B_6_ source and high vitamin B_6_ foods in food groups was as follows: it was excellent in one food group (beef products); it was almost excellent in two food groups (pork products; poultry products); it was very good in two food groups (breakfast cereals; baby foods); it was good in two food groups (finfish and shellfish products; lamb, veal, and game products); it was almost good in one food group (American Indian/Alaska Native foods); it was satisfactory in one food group (restaurant foods); it was acceptable in seven food groups (vegetables and vegetable products; legumes and legume products; sausages and luncheon meats; meals, entrees, and side dishes; nut and seed products; fast foods; snacks); it was almost insufficient in four food groups (beverages; cereal grains and pasta; baked products; fruits and fruit juices); it was insufficient in four food groups (soups, sauces, and gravies; dairy and egg products; sweets; fats and oils); and it was absent in one food group (spices and herbs) (Figure 34). High vitamin B_6_ and vitamin B_6_ source foods based on the proposed method are given in Tables S46 and S47, respectively.

**Figure 34:**
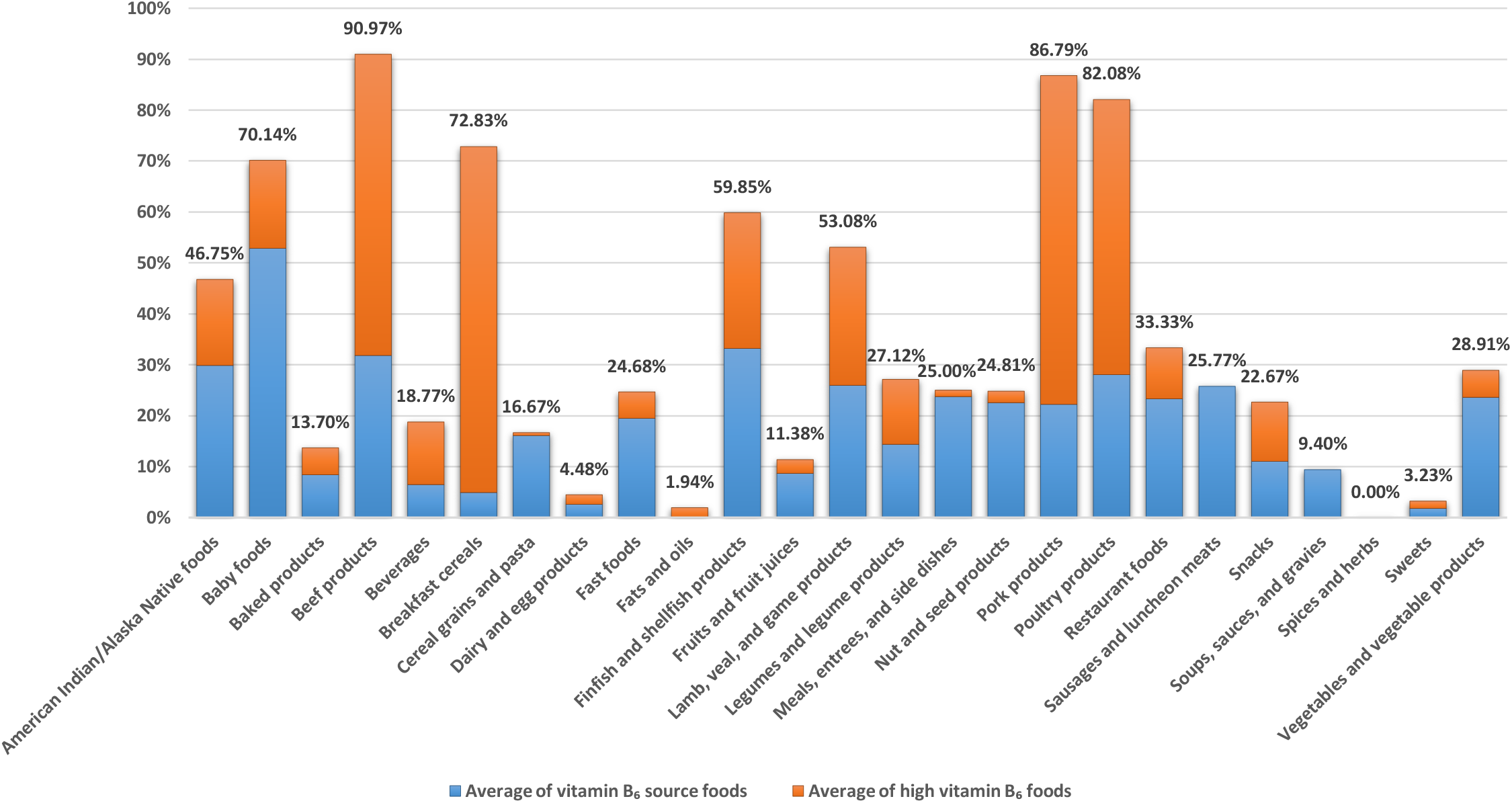
Average of vitamin B_6_ source and high vitamin B_6_ foods based on the proposed method in food groups. All vitamin B_6_ source and high vitamin B_6_ foods except the vitamin B_6_ source and high vitamin B_6_ baby foods are based on the reference energy intake of 2,000 kcal for adults and children aged 4 years and older. The vitamin B_6_ source and high vitamin B_6_ baby foods are based on the reference energy intake of 1,000 kcal for children 1 through 3 years of age.

### Vitamin B_12_ source and high vitamin B_12_ foods

Vitamin B_12_ (also known as cobalamin) functions as a coenzyme for a critical methyl transfer reaction that converts homocysteine to methionine and for a separate reaction that converts L-methylmalonylcoenzyme A (CoA) to succinyl-CoA (IOM, 1998). Humans eat this vitamin preformed in animal foods: meat, milk, eggs and fish (Truswell, 2007). B_12_ is not supplied by commonly eaten plant foods unless they have been exposed to bacterial action that has produced the vitamin; contaminated with soil, insects, or other substances that contain B_12_; or fortified with B_12_ (e.g., fortified ready-to-eat breakfast cereals and meal replacement formulas) (IOM, 1998). According to the proposed method, the average of vitamin B_12_ source and high vitamin B_12_ foods in food groups was as follows: it was excellent in four food groups (beef products; lamb, veal, and game products; finfish and shellfish products; pork products); it was almost excellent in one food group (sausages and luncheon meats); it was very good in three food groups (poultry products; fast foods; American Indian/Alaska Native foods); it was almost very good in one food group (dairy and egg products); it was good in three food groups (breakfast cereals; baby foods; restaurant foods); it was almost good in one food group (meals, entrees, and side dishes); it was acceptable in three food groups (legumes and legume products; soups, sauces, and gravies; beverages); it was almost insufficient in three food groups (sweets; snacks; baked products); it was insufficient in three food groups (fats and oils; fruits and fruit juices; vegetables and vegetable products); and it was absent in three food groups (cereal grains and pasta; nut and seed products; spices and herbs) (Figure 35). High vitamin B_12_ and vitamin B_12_ source foods based on the proposed method are given in Tables S48 and S49, respectively.

**Figure 35:**
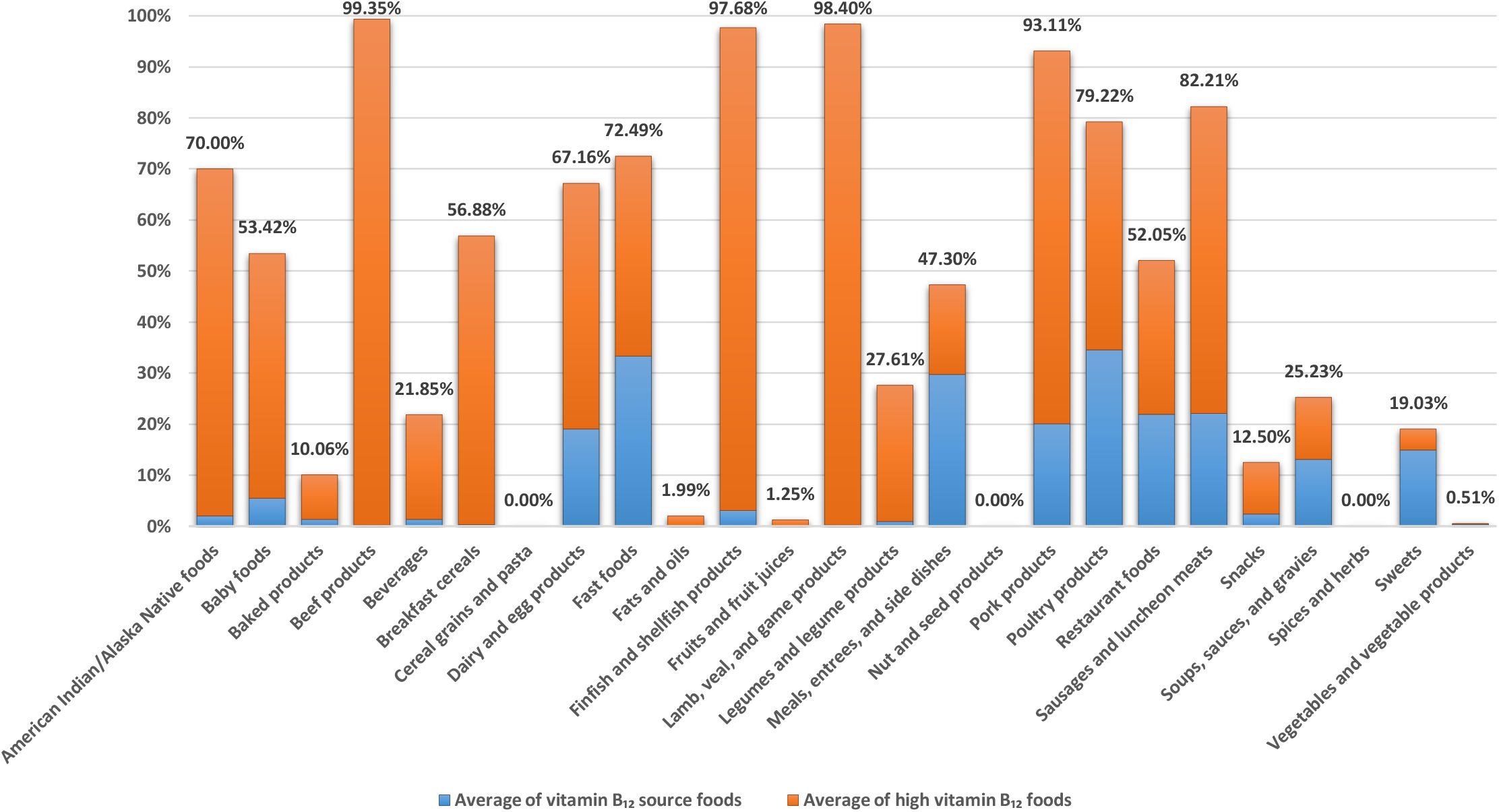
Average of vitamin B_12_ source and high vitamin B_12_ foods based on the proposed method in food groups. All vitamin B_12_ source and high vitamin B_12_ foods except the vitamin B_12_ source and high vitamin B_12_ baby foods are based on the reference energy intake of 2,000 kcal for adults and children aged 4 years and older. The vitamin B_12_ source and high vitamin B_12_ baby foods are based on the reference energy intake of 1,000 kcal for children 1 through 3 years of age.

### Vitamin C source and high vitamin C foods

Vitamin C (also known as L-ascorbic acid) functions physiologically as a water-soluble antioxidant by virtue of its high reducing power (IOM, 2000). It also plays a role in the biosynthesis of carnitine, neurotransmitters, collagen, and other components of connective tissue, and modulates the absorption, transport, and storage of iron (IOM, 2006). Almost 90 percent of vitamin C in the typical diet comes from fruits and vegetables, with citrus fruits, tomatoes and tomato juice, and potatoes being major contributors (Sinha *et al*., 1993). According to the proposed method, the average of vitamin C source and high vitamin C foods in food groups was as follows: it was very good in one food group (baby foods); it was good in two food groups (vegetables and vegetable products; fruits and fruit juices); it was satisfactory in one food group (beverages); it was acceptable in one food group (breakfast cereals); it was almost insufficient in three food groups (American Indian/Alaska Native foods; restaurant foods; snacks); it was insufficient in 14 food groups (meals, entrees, and side dishes; sausages and luncheon meats; sweets; nut and seed products; lamb, veal, and game products; pork products; soups, sauces, and gravies; legumes and legume products; fast foods; poultry products; dairy and egg products; finfish and shellfish products; beef products; baked products); and it was absent in three food groups (cereal grains and pasta; fats and oils; spices and herbs) (Figure 36). High vitamin C and vitamin C source foods based on the proposed method are given in Tables S50 and S51, respectively.

**Figure 36:**
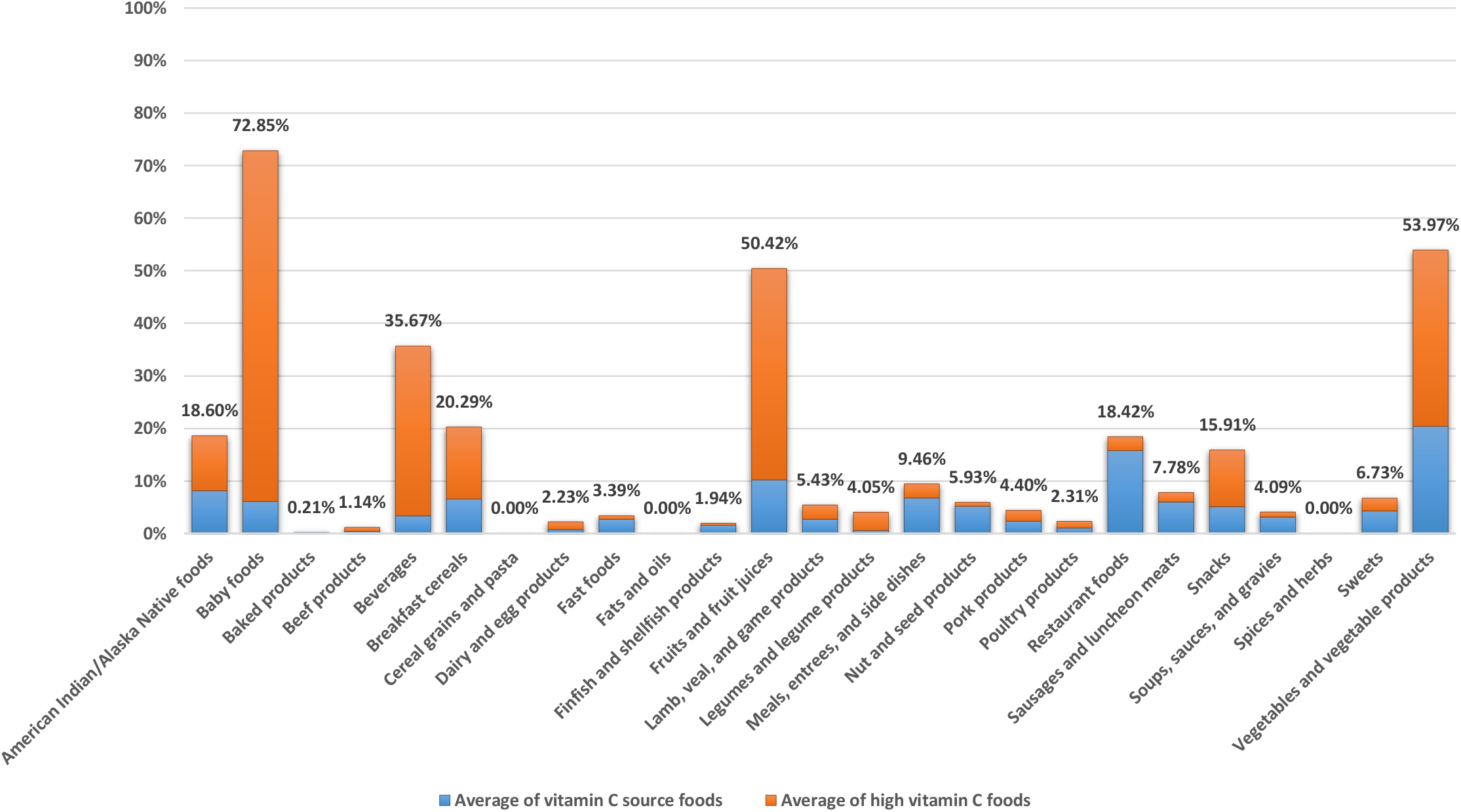
Average of vitamin C source and high vitamin C foods based on the proposed method in food groups. All vitamin C source and high vitamin C foods except the vitamin C source and high vitamin C baby foods are based on the reference energy intake of 2,000 kcal for adults and children aged 4 years and older. The vitamin C source and high vitamin C baby foods are based on the reference energy intake of 1,000 kcal for children 1 through 3 years of age.

### Vitamin D source and high vitamin D foods

Vitamin D (also known as calciferol), which comprises a group of fat soluble seco-sterols that are found in very few foods naturally, is photosynthesized in the skin of vertebrates by the action of solar ultraviolet B radiation (Holick, 1994). The primary function of vitamin D in the body is to aid in the intestinal absorption of calcium and phosphorus, thereby helping maintain normal serum levels of these minerals in the body (IOM, 2006). In nature, very few foods contain vitamin D (IOM, 1997). Those that do include some fish liver oils, the flesh of fatty fish, the liver and fat from aquatic mammals such as seals and polar bears, and eggs from hens that have been fed vitamin D (Holick, 1994; Jeans, 1950). Almost all of the human intake of vitamin D from foods comes from fortified milk products and other fortified foods such as breakfast cereals (IOM, 1997). According to the proposed method, the average of vitamin D source and high vitamin D foods in food groups was as follows: it was almost good in one food group (finfish and shellfish products); it was acceptable in one food group (baby foods); it was almost insufficient in three food groups (dairy and egg products; American Indian/Alaska Native foods; legumes and legume products); it was insufficient in eight food groups (beverages; breakfast cereals; meals, entrees, and side dishes; vegetables and vegetable products; fats and oils; fruits and fruit juices; lamb, veal, and game products; pork products); and it was absent in 12 food groups (baked products; beef products; cereal grains and pasta; fast foods; nut and seed products; poultry products; restaurant foods; sausages and luncheon meats; snacks; soups, sauces, and gravies; spices and herbs; sweets) (Figure 37). High vitamin D and vitamin D source foods based on the proposed method are given in Tables S52 and S53, respectively.

**Figure 37:**
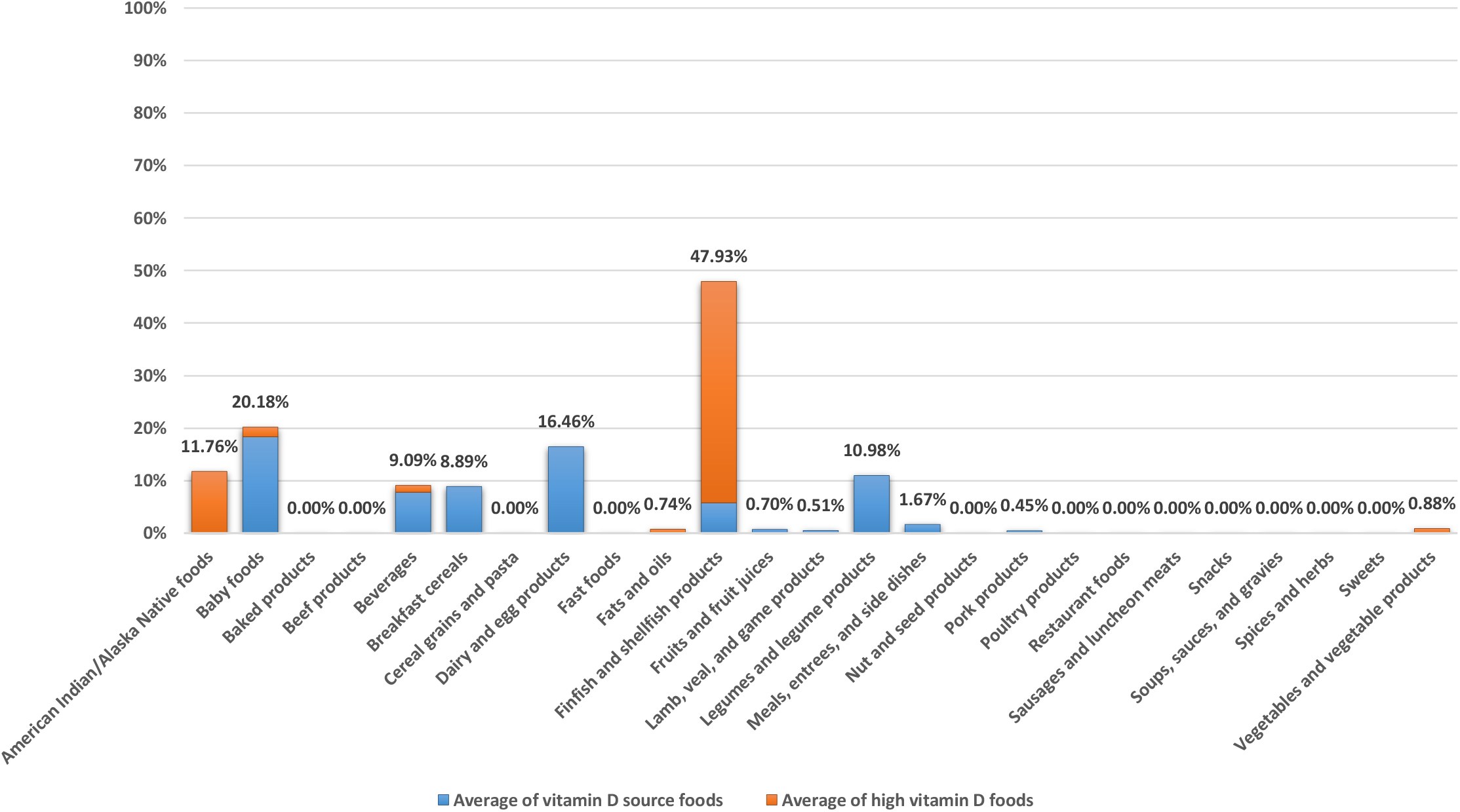
Average of vitamin D source and high vitamin D foods based on the proposed method in food groups. All vitamin D source and high vitamin D foods except the vitamin D source and high vitamin D baby foods are based on the reference energy intake of 2,000 kcal for adults and children aged 4 years and older. The vitamin D source and high vitamin D baby foods are based on the reference energy intake of 1,000 kcal for children 1 through 3 years of age.

### Vitamin E source and high vitamin E foods

Vitamin E (a fat-soluble vitamin) is thought to function primarily as a chain-breaking antioxidant that prevents the propagation of lipid peroxidation (IOM, 2000). Of the eight naturally occurring forms of vitamin E only the α-tocopherol form of the vitamin is maintained in the plasma (IOM, 2006). The main dietary sources of vitamin E are edible vegetable oils (Dial and Eitenmiller, 1995; McLaughlin and Weihrauch, 1979; Sheppard *et al*., 1993). According to the proposed method, the average of vitamin E source and high vitamin E foods in food groups was as follows: it was almost good in two food groups (baby foods; nut and seed products); it was satisfactory in one food group (snacks); it was acceptable in one food group (fats and oils); it was almost insufficient in six food groups (finfish and shellfish products; legumes and legume products; beverages; vegetables and vegetable products; American Indian/Alaska Native foods; breakfast cereals); it was insufficient in nine food groups (soups, sauces, and gravies; fruits and fruit juices; fast foods; meals, entrees, and side dishes; restaurant foods; sweets; dairy and egg products; baked products; beef products); and it was absent in six food groups (cereal grains and pasta; lamb, veal, and game products; pork products; poultry products; sausages and luncheon meats; spices and herbs) (Figure 38). High vitamin E and vitamin E source foods based on the proposed method are given in Tables S54 and S55, respectively.

**Figure 38:**
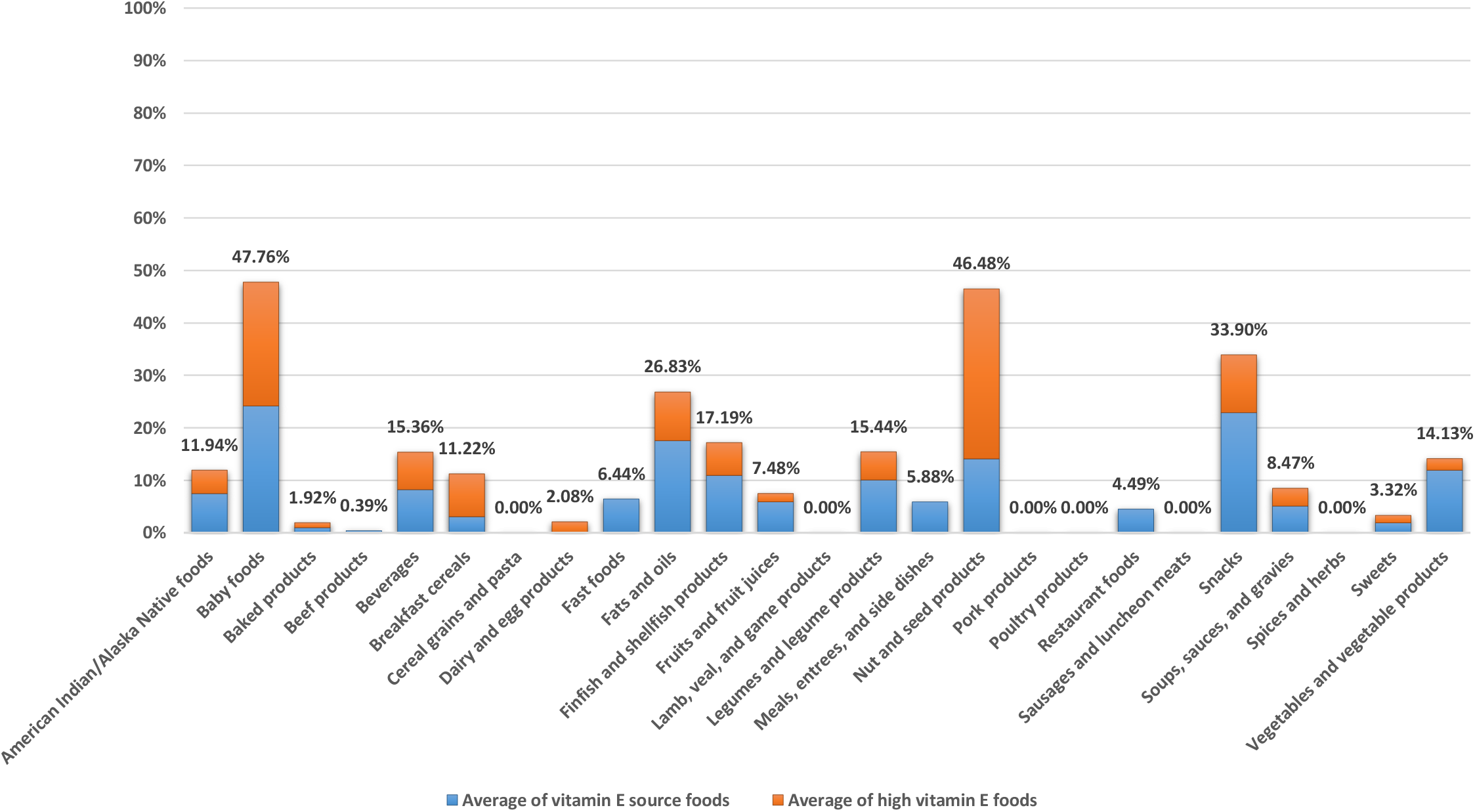
Average of vitamin E source and high vitamin E foods based on the proposed method in food groups. All vitamin E source and high vitamin E foods except the vitamin E source and high vitamin E baby foods are based on the reference energy intake of 2,000 kcal for adults and children aged 4 years and older. The vitamin E source and high vitamin E baby foods are based on the reference energy intake of 1,000 kcal for children 1 through 3 years of age.

### Vitamin K source and high vitamin K foods

Vitamin K (a fat-soluble vitamin) functions as a coenzyme during the synthesis of the biologically active form of a number of proteins involved in blood coagulation and bone metabolism (IOM, 2001). Phylloquinone, the plant form of vitamin K, is the major form in the diet (IOM, 2006). Spinach, collards, broccoli, and iceberg lettuce are the major contributors of vitamin K in the diet of U.S. adults and children (IOM, 2001; Sokol *et al*., 2006). The breast-fed infant receives vitamin K through ingestion of mother’s milk (Sokol *et al*., 2006). According to the proposed method, the average of vitamin K source and high vitamin K foods in food groups was as follows: it was almost very good in one food group (restaurant foods); it was good in one food group (baby foods); it was almost good in one food group (vegetables and vegetable products); it was acceptable in two food groups (fats and oils; meals, entrees, and side dishes); it was almost insufficient in four food groups (soups, sauces, and gravies; American Indian/Alaska Native foods; fruits and fruit juices; fast foods); it was insufficient in 11 food groups (snacks; spices and herbs; baked products; beverages; legumes and legume products; sausages and luncheon meats; nut and seed products; cereal grains and pasta; dairy and egg products; finfish and shellfish products; beef products); and it was absent in five food groups (breakfast cereals; lamb, veal, and game products; pork products; poultry products; sweets) (Figure 39). High vitamin K and vitamin K source foods based on the proposed method are given in Tables S56 and S57, respectively.

**Figure 39:**
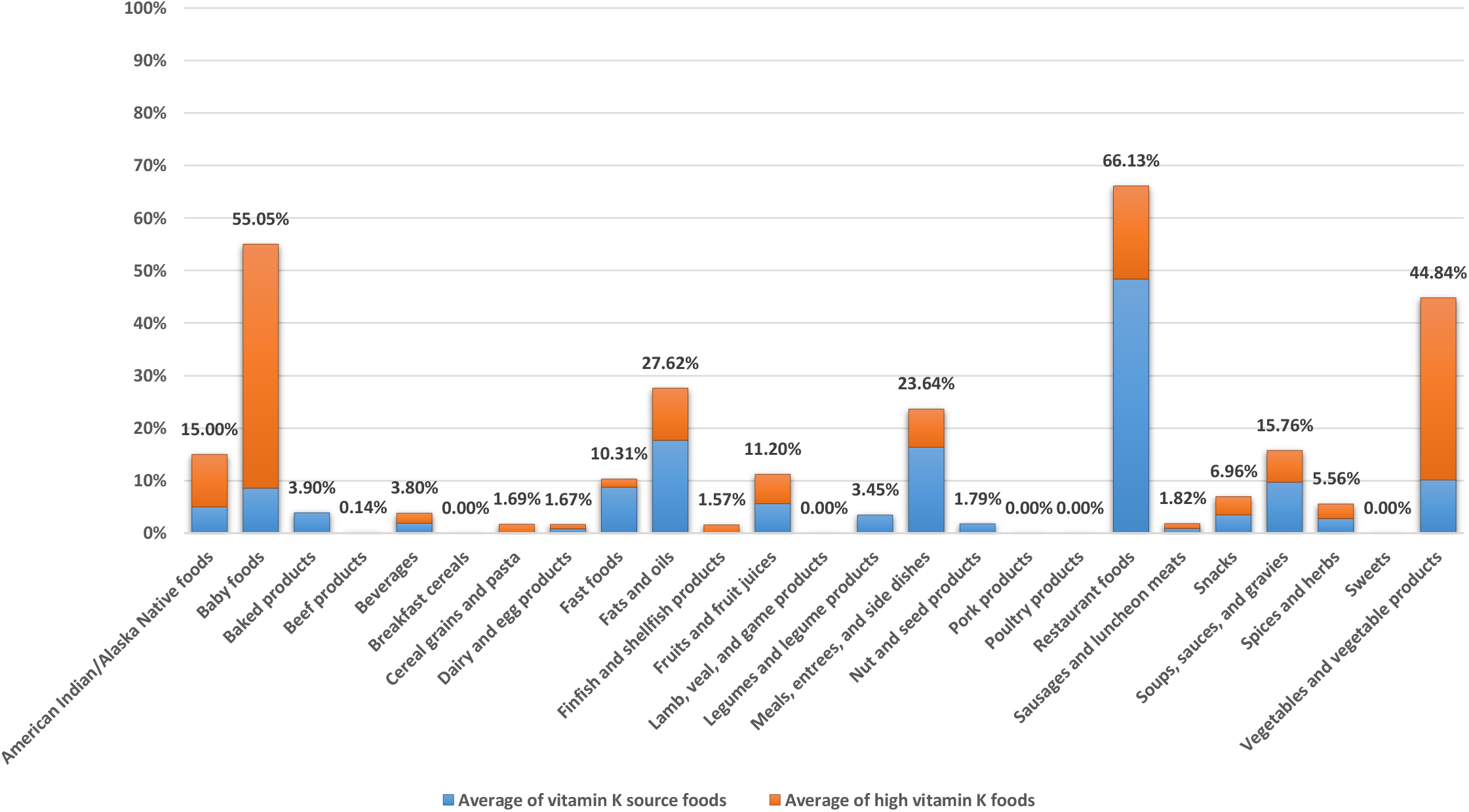
Average of vitamin K source and high vitamin K foods based on the proposed method in food groups. All vitamin K source and high vitamin K foods except the vitamin K source and high vitamin K baby foods are based on the reference energy intake of 2,000 kcal for adults and children aged 4 years and older. The vitamin K source and high vitamin K baby foods are based on the reference energy intake of 1,000 kcal for children 1 through 3 years of age.

### Zinc source and high zinc foods

Zinc functions as a component of various enzymes in the maintenance of the structural integrity of proteins and in the regulation of gene expression (IOM, 2001). Zinc is abundant in red meats, certain seafood, and whole grains (IOM, 2001). According to the proposed method, the average of zinc source and high zinc foods in food groups was as follows: it was excellent in three food groups (beef products; lamb, veal, and game products; pork products); it was almost excellent in one food group (poultry products); it was almost very good in two food groups (breakfast cereals; baby foods); it was good in one food group (sausages and luncheon meats); it was almost good in one food group (nut and seed products); it was satisfactory in four food groups (fast foods; American Indian/Alaska Native foods; legumes and legume products; restaurant foods); it was acceptable in four food groups (finfish and shellfish products; meals, entrees, and side dishes; dairy and egg products; cereal grains and pasta); it was almost insufficient in two food groups (soups, sauces, and gravies; snacks); it was insufficient in four food groups (beverages; vegetables and vegetable products; sweets; baked products); and it was absent in three food groups (fats and oils; fruits and fruit juices; spices and herbs) (Figure 40). High zinc and zinc source foods based on the proposed method are given in Tables S58 and S59, respectively.

**Figure 40:**
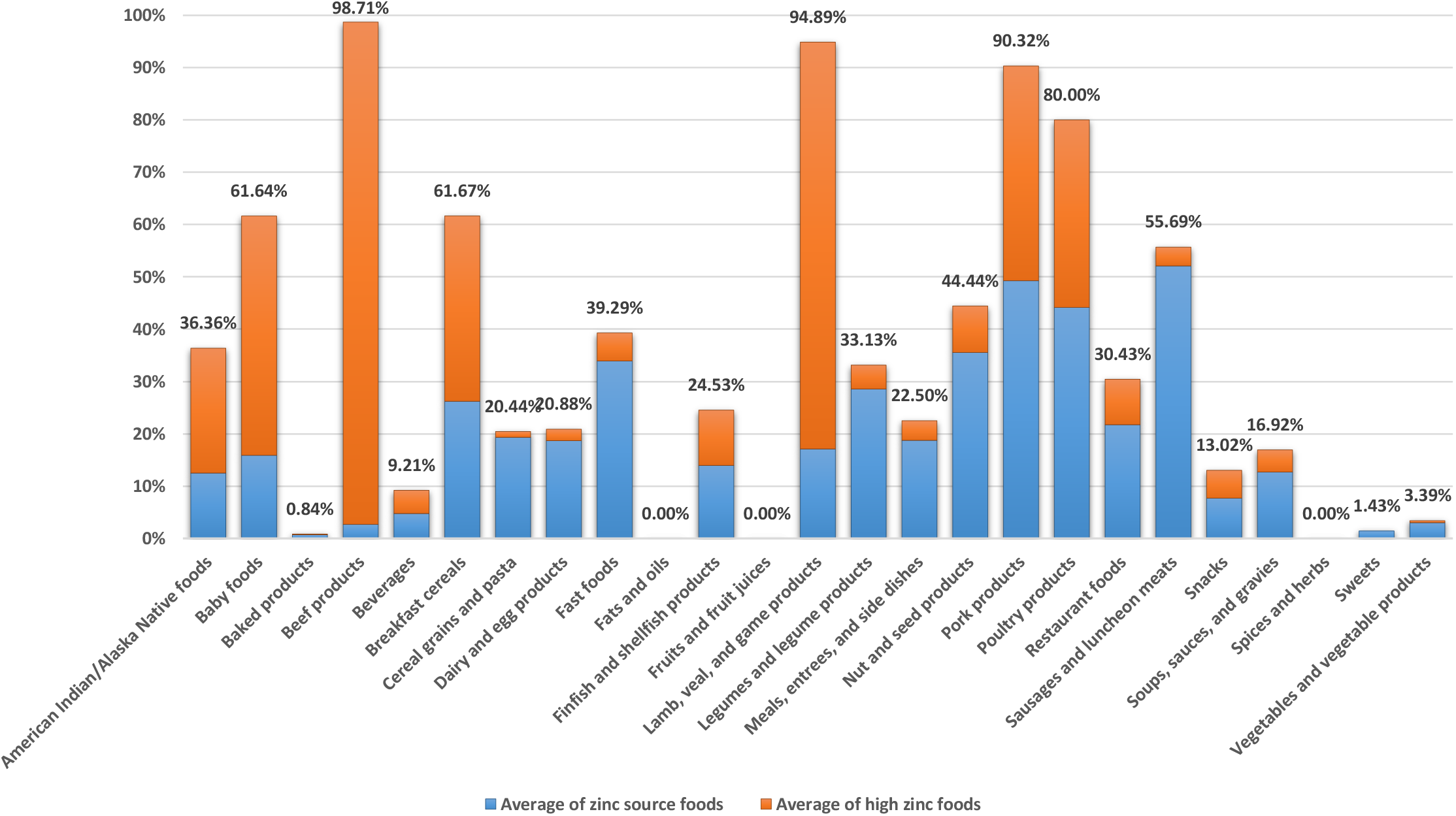
Average of zinc source and high zinc foods based on the proposed method in food groups. All zinc source and high zinc foods except the zinc source and high zinc baby foods are based on the reference energy intake of 2,000 kcal for adults and children aged 4 years and older. The zinc source and high zinc baby foods are based on the reference energy intake of 1,000 kcal for children 1 through 3 years of age.

### Average of nutrient source and high nutrient foods in food groups

The average of nutrient source and high nutrient foods based on the proposed method in food groups is given in Table S60. The average of nutrient source and high nutrient foods in American Indian/Alaska Native foods was as follows: it was almost very good to very good for protein, vitamin B_12_, and selenium; it was almost good to good for iron, copper, pantothenic acid, vitamin B_6_, riboflavin, niacin, and phosphorus; it was acceptable to satisfactory for vitamin A, thiamin, manganese, zinc, dietary fiber, and choline; and it was insufficient to almost insufficient for potassium, calcium, vitamin D, vitamin E, folate, vitamin K, magnesium, and vitamin C. The average of nutrient source and high nutrient foods in baby foods was as follows: it was almost excellent for copper; it was almost very good to very good for zinc, niacin, protein, riboflavin, vitamin B_6_, and vitamin C; it was almost good to good for calcium, phosphorus, vitamin E, folate, iron, thiamin, selenium, vitamin B_12_, magnesium, vitamin A, vitamin K, and pantothenic acid; it was acceptable for vitamin D, dietary fiber, and manganese; and it was insufficient to almost insufficient for potassium and choline.

The average of nutrient source and high nutrient foods in baked products was as follows: it was almost good to good for folate, thiamin, and selenium; it was acceptable to satisfactory for copper, niacin, riboflavin, and manganese; it was insufficient to almost insufficient for vitamin C, choline, zinc, pantothenic acid, vitamin E, magnesium, vitamin K, calcium, protein, dietary fiber, vitamin B_12_, phosphorus, vitamin B_6_, iron, and vitamin A; and it was absent for potassium and vitamin D.

The average of nutrient source and high nutrient foods in beef products was as follows: it was almost excellent to excellent for choline, phosphorus, vitamin B_6_, selenium, niacin, protein, zinc, and vitamin B_12_; it was almost very good to very good for iron and riboflavin; it was almost good for pantothenic acid; it was satisfactory for copper; it was insufficient for potassium, vitamin K, calcium, vitamin E, folate, vitamin A, manganese, vitamin C, and thiamin; and it was absent for dietary fiber, magnesium, and vitamin D. The average of nutrient source and high nutrient foods in beverages was as follows: it was acceptable to satisfactory for manganese, pantothenic acid, vitamin B_12_, riboflavin, and vitamin C; and it was insufficient to almost insufficient for choline, dietary fiber, vitamin K, potassium, iron, folate, selenium, magnesium, vitamin D, zinc, protein, vitamin A, phosphorus, calcium, thiamin, copper, vitamin E, vitamin B_6_, and niacin.

The average of nutrient source and high nutrient foods in breakfast cereals was as follows: it was almost excellent to excellent for niacin and thiamin; it was almost very good to very good for zinc, vitamin B_6_, manganese, riboflavin, folate, and iron; it was good for vitamin A, vitamin B_12_, selenium, dietary fiber, and copper; it was acceptable to satisfactory for vitamin C, protein, calcium, magnesium, and phosphorus; it was insufficient to almost insufficient for choline, potassium, vitamin D, vitamin E, and pantothenic acid; and it was absent for vitamin K.

The average of nutrient source and high nutrient foods in cereal grains and pasta was as follows: it was almost very good to very good for selenium and manganese; it was good for niacin, copper, and thiamin; it was acceptable to satisfactory for zinc, iron, riboflavin, phosphorus, magnesium, dietary fiber, protein, and folate; it was insufficient to almost insufficient for vitamin K, pantothenic acid, and vitamin B_6_; and it was absent for calcium, choline, potassium, vitamin A, vitamin B_12_, vitamin C, vitamin D, and vitamin E.

The average of nutrient source and high nutrient foods in dairy and egg products was as follows: it was almost very good for phosphorus, vitamin B_12_, and protein; it was almost good to good for selenium, pantothenic acid, riboflavin, and calcium; it was acceptable for zinc and vitamin A; and it was insufficient to almost insufficient for manganese, dietary fiber, vitamin K, vitamin E, vitamin C, niacin, iron, potassium, magnesium, thiamin, vitamin B_6_, folate, choline, copper, and vitamin D. The dairy group contributes many nutrients, including calcium, phosphorus, vitamin A, vitamin D (in products fortified with vitamin D), riboflavin, vitamin B_12_, protein, potassium, zinc, choline, magnesium, and selenium (HHS and USDA, 2015).

The average of nutrient source and high nutrient foods in fast foods was as follows: it was almost excellent to excellent for protein and selenium; it was almost very good to very good for phosphorus, thiamin, folate, vitamin B_12_, riboflavin, and niacin; it was acceptable to satisfactory for calcium, iron, vitamin B_6_, copper, manganese, pantothenic acid, and zinc; it was insufficient to almost insufficient for magnesium, potassium, vitamin C, vitamin A, vitamin E, dietary fiber, vitamin K, and choline; and it was absent for vitamin D. The average of nutrient source and high nutrient foods in fats and oils was as follows: it was acceptable for vitamin E and vitamin K; it was insufficient to almost insufficient for vitamin D, thiamin, vitamin B_6_, vitamin B_12_, and vitamin A; and it was absent for calcium, choline, copper, dietary fiber, folate, iron, magnesium, manganese, niacin, pantothenic acid, phosphorus, potassium, protein, riboflavin, selenium, vitamin C, and zinc. Oils provide essential fatty acids and vitamin E (HHS and USDA, 2015).

The average of nutrient source and high nutrient foods in finfish and shellfish products was as follows: it was almost excellent to excellent for choline, phosphorus, vitamin B_12_, protein, and selenium; it was very good for niacin; it was almost good to good for vitamin D, pantothenic acid, and vitamin B_6_; it was acceptable to satisfactory for thiamin, zinc, riboflavin, and copper; it was insufficient to almost insufficient for vitamin K, vitamin C, calcium, folate, potassium, vitamin A, iron, manganese, magnesium, and vitamin E; and it was absent for dietary fiber.

The average of nutrient source and high nutrient foods in fruits and fruit juices was as follows: it was almost good to good for copper and vitamin C; it was acceptable to satisfactory for manganese and dietary fiber; it was insufficient to almost insufficient for protein, phosphorus, vitamin D, vitamin B_12_, calcium, magnesium, pantothenic acid, niacin, vitamin A, riboflavin, iron, potassium, vitamin E, thiamin, folate, vitamin K, and vitamin B_6_; and it was absent for choline, selenium, and zinc. Among the many nutrients fruits provide are dietary fiber, potassium, and vitamin C (HHS and USDA, 2015).

The average of nutrient source and high nutrient foods in lamb, veal, and game products was as follows: it was almost excellent to excellent for phosphorus, riboflavin, choline, zinc, niacin, protein, and vitamin B_12_; it was almost very good to very good for pantothenic acid, copper, and selenium; it was good for vitamin B_6_; it was acceptable to satisfactory for thiamin and iron; it was insufficient for magnesium, potassium, vitamin D, manganese, folate, vitamin A, and vitamin C; and it was absent for calcium, dietary fiber, vitamin E, and vitamin K.

The average of nutrient source and high nutrient foods in legumes and legume products was as follows: it was almost excellent to excellent for protein, manganese, and copper; it was almost very good for folate and magnesium; it was almost good to good for iron, phosphorus, dietary fiber, and thiamin; it was acceptable to satisfactory for pantothenic acid, selenium, vitamin B_6_, vitamin B_12_, niacin, riboflavin, and zinc; and it was insufficient to almost insufficient for vitamin K, vitamin A, vitamin C, vitamin D, choline, vitamin E, calcium, and potassium. Legumes are excellent sources of protein, dietary fiber, potassium, folate, iron, and zinc (HHS and USDA, 2015).

The average of nutrient source and high nutrient foods in meals, entrees, and side dishes was as follows: it was almost excellent to excellent for niacin, protein, and selenium; it was almost very good to very good for riboflavin, thiamin, folate, and manganese; it was almost good to good for phosphorus, vitamin B_12_, and copper; it was acceptable to satisfactory for zinc, vitamin K, vitamin B_6_, iron, pantothenic acid, and dietary fiber; and it was insufficient to almost insufficient for vitamin D, vitamin E, vitamin A, magnesium, potassium, choline, vitamin C, and calcium.

The average of nutrient source and high nutrient foods in nut and seed products was as follows: it was almost excellent to excellent for manganese and copper; it was almost good to good for zinc, vitamin E, protein, phosphorus, and magnesium; it was acceptable to satisfactory for niacin, iron, vitamin B_6_, dietary fiber, selenium, and thiamin; it was insufficient to almost insufficient for vitamin K, potassium, calcium, vitamin C, pantothenic acid, folate, and riboflavin; and it was absent for choline, vitamin A, vitamin B_12_, and vitamin D.

The average of nutrient source and high nutrient foods in pork products was as follows: it was almost excellent to excellent for vitamin B_6_, phosphorus, choline, riboflavin, thiamin, zinc, niacin, vitamin B_12_, selenium, and protein; it was very good for pantothenic acid; it was satisfactory for copper; it was insufficient for calcium, vitamin D, vitamin A, manganese, folate, vitamin C, iron, and potassium; and it was absent for dietary fiber, magnesium, vitamin E, and vitamin K.

The average of nutrient source and high nutrient foods in poultry products was as follows: it was almost excellent to excellent for zinc, phosphorus, vitamin B_6_, pantothenic acid, niacin, selenium, and protein; it was almost very good to very good for choline, riboflavin, and vitamin B_12_, it was acceptable to satisfactory for iron and copper; it was insufficient to almost insufficient for calcium, magnesium, potassium, manganese, vitamin C, folate, vitamin A, and thiamin; and it was absent for dietary fiber, vitamin D, vitamin E, and vitamin K.

The average of nutrient source and high nutrient foods in restaurant foods was as follows: it was almost very good to very good for vitamin K, protein, and selenium; it was almost good to good for niacin, vitamin B_12_, and phosphorus; it was acceptable to satisfactory for copper, manganese, zinc, folate, vitamin B_6_, pantothenic acid, and riboflavin; it was insufficient to almost insufficient for potassium, magnesium, vitamin E, vitamin A, dietary fiber, choline, iron, calcium, vitamin C, and thiamin; and it was absent for vitamin D.

The average of nutrient source and high nutrient foods in sausages and luncheon meats was as follows: it was almost excellent to excellent for vitamin B_12_, selenium, and protein; it was almost very good for niacin; it was almost good to good for thiamin and zinc; it was acceptable for riboflavin and vitamin B_6_; it was insufficient to almost insufficient for calcium, potassium, folate, vitamin K, choline, manganese, vitamin A, vitamin C, iron, phosphorus, copper, and pantothenic acid; and it was absent for dietary fiber, magnesium, vitamin D, and vitamin E.

The average of nutrient source and high nutrient foods in snacks was as follows: it was almost very good for manganese; it was almost good for copper; it was acceptable to satisfactory for folate, vitamin B_6_, selenium, thiamin, niacin, and vitamin E; it was insufficient to almost insufficient for potassium, vitamin K, phosphorus, pantothenic acid, vitamin A, iron, protein, vitamin B_12_, calcium, zinc, dietary fiber, vitamin C, magnesium, and riboflavin; and it was absent for choline and vitamin D.

The average of nutrient source and high nutrient foods in soups, sauces, and gravies was as follows: it was almost good for manganese and copper; it was acceptable for dietary fiber, vitamin B_12_, protein, and selenium; it was insufficient to almost insufficient for choline, magnesium, calcium, vitamin C, folate, thiamin, iron, vitamin E, vitamin B_6_, phosphorus, pantothenic acid, vitamin K, vitamin A, potassium, zinc, niacin, and riboflavin; and it was absent for vitamin D.

The average of nutrient source and high nutrient foods in spices and herbs was as follows: it was insufficient for manganese and vitamin K, and it was absent for calcium, choline, copper, dietary fiber, folate, iron, magnesium, niacin, pantothenic acid, phosphorus, potassium, protein, riboflavin, selenium, thiamin, vitamin A, vitamin B_6_, vitamin B_12_, vitamin C, vitamin D, vitamin E, and zinc.

The average of nutrient source and high nutrient foods in sweets was as follows: it was acceptable for copper; it was insufficient to almost insufficient for thiamin, zinc, niacin, selenium, iron, protein, dietary fiber, vitamin B_6_, vitamin E, magnesium, vitamin A, pantothenic acid, vitamin C, phosphorus, manganese, calcium, vitamin B_12_, and riboflavin; and it was absent for choline, folate, potassium, vitamin D, and vitamin K.

The average of nutrient source and high nutrient foods in vegetables and vegetable products was as follows: it was almost good to good for manganese, vitamin K, copper, and vitamin C; it was acceptable to satisfactory for vitamin A, vitamin B_6_, folate, and dietary fiber; and it was insufficient to almost insufficient for vitamin B_12_, vitamin D, choline, zinc, selenium, phosphorus, protein, calcium, niacin, vitamin E, iron, magnesium, potassium, pantothenic acid, riboflavin, and thiamin. Vegetables are important sources of many nutrients, including dietary fiber, potassium, vitamin A (in the form of provitamin A carotenoids), vitamin C, vitamin K, copper, magnesium, vitamin E, vitamin B_6_, folate, iron, manganese, thiamin, niacin, and choline (HHS and USDA, 2015).

### Averages of nutrient free and low nutrient foods in food groups

Averages of nutrient free and low nutrient foods based on the proposed method in food groups are given in Tables S61 and S62, respectively. The average of nutrient free foods in American Indian/Alaska Native foods was as follows: it was acceptable for sodium; it was insufficient to almost insufficient for cholesterol, saturated fat, and total fat; and it was absent for energy. Also, the average of low nutrient foods in American Indian/Alaska Native foods was as follows: it was almost very good to very good for saturated fat and sodium; it was good for total fat; it was satisfactory for energy; and it was insufficient for cholesterol.

The average of nutrient free foods in baby foods was as follows: it was acceptable to satisfactory for saturated fat, sodium, and cholesterol; and it was insufficient to almost insufficient for energy and total fat. Also, the average of low nutrient foods in baby foods was as follows: it was excellent for sodium; it was almost good for cholesterol, saturated fat, and total fat; and it was insufficient for energy.

The average of nutrient free foods in baked products was as follows: it was acceptable for cholesterol; and it was insufficient for energy, sodium, total fat, and saturated fat. Also, the average of low nutrient foods in baked products was as follows: it was acceptable to satisfactory for sodium, total fat, cholesterol, and saturated fat, and it was insufficient for energy.

The average of nutrient free foods in beef products was as follows: it was insufficient for sodium; and it was absent for cholesterol, energy, saturated fat, and total fat. Also, the average of low nutrient foods in beef products was as follows: it was excellent for sodium; it was insufficient for energy, saturated fat, and total fat; and it was absent for cholesterol.

The average of nutrient free foods in beverages was as follows: it was almost excellent for saturated fat and cholesterol; it was very good for total fat; it was satisfactory for sodium; and it was almost insufficient for energy. Also, the average of low nutrient foods in beverages was as follows: it was almost excellent to excellent for saturated fat, cholesterol, and total fat; it was very good for sodium; and it was almost good for energy.

The average of nutrient free foods in breakfast cereals was as follows: it was excellent for cholesterol; it was insufficient to almost insufficient for total fat, saturated fat, and sodium; and it was absent for energy. Also, the average of low nutrient foods in breakfast cereals was as follows: it was almost excellent to excellent for total fat, cholesterol, and saturated fat; it was acceptable for sodium; and it was insufficient for energy.

The average of nutrient free foods in cereal grains and pasta was as follows: it was excellent for cholesterol; it was very good for sodium; it was acceptable to satisfactory for total fat and saturated fat; and it was absent for energy. Also, the average of low nutrient foods in cereal grains and pasta was as follows: it was almost excellent to excellent for sodium, cholesterol, saturated fat, and total fat; and it was insufficient for energy. The average of nutrient free foods in dairy and egg products was as follows: it was insufficient to almost insufficient for sodium, cholesterol, saturated fat, and total fat; and it was absent for energy. Also, the average of low nutrient foods in dairy and egg products was as follows: it was almost good for sodium and energy; and it was acceptable to satisfactory for saturated fat, cholesterol, and total fat.

The average of nutrient free foods in fast foods was as follows: it was insufficient for sodium, cholesterol, saturated fat, and total fat; and it was absent for energy. Also, the average of low nutrient foods in fast foods was as follows: it was insufficient for sodium, cholesterol, energy, saturated fat, and total fat. There is a growing body of evidence linking fast food intake to increased calorie consumption, body weight, and fat intake (Binkley *et al*., 2000; Bowman and Vinyard, 2004; French *et al*., 2000; Paeratakul *et al*., 2003). Fast food tends to be more energy dense, and higher in saturated fat and sodium (Isganaitis and Lustig, 2005; Nielsen and Popkin, 2003; Pereira *et al*., 2005).

The average of nutrient free foods in fats and oils was as follows: it was good for sodium; and it was insufficient to almost insufficient for energy, saturated fat, total fat, and cholesterol. Also, the average of low nutrient foods in fats and oils was as follows: it was good for sodium; it was acceptable for cholesterol and energy; and it was insufficient to almost insufficient for total fat and saturated fat.

The average of nutrient free foods in finfish and shellfish products was as follows: it was insufficient for energy, cholesterol, total fat, and saturated fat; and it was absent for sodium. Also, the average of low nutrient foods in finfish and shellfish products was as follows: it was almost very good for sodium and saturated fat; it was good for total fat; it was satisfactory for energy; and it was insufficient for cholesterol. The average of nutrient free foods in fruits and fruit juices was as follows: it was almost excellent to excellent for sodium, saturated fat, and cholesterol; it was very good for total fat; and it was insufficient for energy. Also, the average of low nutrient foods in fruits and fruit juices was as follows: it was excellent for saturated fat, total fat, sodium, and cholesterol; and it was good for energy.

The average of nutrient free foods in lamb, veal, and game products was as follows: it was absent for cholesterol, energy, saturated fat, sodium, and total fat. Also, the average of low nutrient foods in lamb, veal, and game products was as follows: it was excellent for sodium; it was insufficient to almost insufficient for energy, saturated fat, and total fat; and it was absent for cholesterol.

The average of nutrient free foods in legumes and legume products was as follows: it was very good for cholesterol; it was acceptable for saturated fat and sodium; it was almost insufficient for total fat; and it was absent for energy. Also, the average of low nutrient foods in legumes and legume products was as follows: it was very good for saturated fat and cholesterol; it was almost good to good for sodium and total fat; and it was almost insufficient for energy.

The average of nutrient free foods in meals, entrees, and side dishes was as follows: it was insufficient for sodium, saturated fat, and cholesterol; and it was absent for energy and total fat. Also, the average of low nutrient foods in meals, entrees, and side dishes was as follows: it was acceptable for cholesterol and saturated fat; it was insufficient to almost insufficient for sodium and total fat; and it was absent for energy. The average of nutrient free foods in nut and seed products was as follows: it was almost very good for sodium; it was acceptable for cholesterol; it was insufficient to almost insufficient for total fat and saturated fat; and it was absent for energy. Also, the average of low nutrient foods in nut and seed products was as follows: it was almost excellent for sodium; it was acceptable for total fat, saturated fat, and cholesterol; and it was almost insufficient for energy.

The average of nutrient free foods in pork products was as follows: it was insufficient for sodium; and it was absent for cholesterol, energy, saturated fat, and total fat. Also, the average of low nutrient foods in pork products was as follows: it was good for sodium; it was insufficient to almost insufficient for energy, saturated fat, and total fat; and it was absent for cholesterol.

The average of nutrient free foods in poultry products was as follows: it was absent for cholesterol, energy, saturated fat, sodium, and total fat. Also, the average of low nutrient foods in poultry products was as follows: it was almost very good for sodium; it was acceptable for total fat; it was insufficient to almost insufficient for energy and saturated fat; and it was absent for cholesterol.

The average of nutrient free foods in restaurant foods was as follows: it was insufficient for cholesterol; and it was absent for energy, saturated fat, sodium, and total fat. Also, the average of low nutrient foods in restaurant foods was as follows: it was insufficient for energy, total fat, cholesterol, sodium, and saturated fat.

The average of nutrient free foods in sausages and luncheon meats was as follows: it was insufficient for sodium, total fat, and saturated fat; and it was absent for cholesterol and energy. Also, the average of low nutrient foods in sausages and luncheon meats was as follows: it was acceptable for energy; and it was insufficient to almost insufficient for cholesterol, sodium, saturated fat, and total fat.

The average of nutrient free foods in snacks was as follows: it was almost good for cholesterol; it was insufficient to almost insufficient for total fat, saturated fat, and sodium; and it was absent for energy. Also, the average of low nutrient foods in snacks was as follows: it was almost good for saturated fat, cholesterol, and sodium; it was satisfactory for total fat; and it was insufficient for energy.

The average of nutrient free foods in soups, sauces, and gravies was as follows: it was acceptable to satisfactory for saturated fat and cholesterol; and it was insufficient to almost insufficient for sodium, energy, and total fat. Also, the average of low nutrient foods in soups, sauces, and gravies was as follows: it was almost very good for energy, cholesterol, and total fat; it was good for saturated fat; and it was insufficient for sodium.

The average of nutrient free foods in spices and herbs was as follows: it was almost excellent for cholesterol; it was almost good for sodium; it was satisfactory for total fat and saturated fat; and it was almost insufficient for energy. Also, the average of low nutrient foods in spices and herbs was as follows: it was almost excellent for saturated fat, energy, sodium, and cholesterol; and it was almost very good for total fat.

The average of nutrient free foods in sweets was as follows: it was acceptable to satisfactory for sodium, saturated fat, total fat, and cholesterol; and it was insufficient for energy. Also, the average of low nutrient foods in sweets was as follows: it was almost excellent for sodium; it was good for total fat; it was satisfactory for cholesterol and saturated fat; and it was almost insufficient for energy.

The average of nutrient free foods in vegetables and vegetable products was as follows: it was excellent for cholesterol; it was almost very good to very good for total fat and saturated fat; it was acceptable for sodium; and it was insufficient for energy. Also, the average of low nutrient foods in vegetables and vegetable products was as follows: it was almost excellent to excellent for energy, total fat, saturated fat, and cholesterol; and it was almost very good for sodium.

### Similarities between nutrient content claims in food groups

The number of 95% similarities or more between nutrient content claims in food groups was as follows: it was 1121 in spices and herbs; it was 1030 in fats and oils; it was 817 in beef products; it was 809 in sweets; it was 646 in sausages and luncheon meats; it was 515 in pork products; it was 482 in fruits and fruit juices; it was 466 in lamb, veal, and game products; it was 455 in baked products; it was 417 in poultry products; it was 382 in dairy and egg products; it was 374 in fast foods; it was 368 in cereal grains and pasta; it was 321 in restaurant foods; it was 292 in nut and seed products; it was 242 in soups, sauces, and gravies; it was 222 in meals, entrees, and side dishes; it was 218 in snacks; it was 200 in vegetables and vegetable products; it was 188 in finfish and shellfish products; it was 157 in beverages; it was 119 in breakfast cereals; it was 101 in legumes and legume products; it was 86 in American Indian/Alaska Native foods; and it was 20 in baby foods. Similarities between nutrient content claims under the proposed method in 25 food groups are given in Tables S63-S87.

### Food rankings based on the score for met claims of the nutrient content

Food groups were ranked based on the average of scores for source and high nutrients as follows: (1) breakfast cereals; (2) baby foods; (3) pork products; (4) lamb, veal, and game products; (5) poultry products; (6) beef products; (7) American Indian/Alaska Native foods; (8) finfish and shellfish products; (9) legumes and legume products; (10) fast foods; (11) meals, entrees, and side dishes; (12) restaurant foods; (13) nut and seed products; (14) cereal grains and pasta; (15) sausages and luncheon meats; (16) dairy and egg products; (17) snacks; (18) vegetables and vegetable products; (19) beverages; (20) soups, sauces, and gravies; (21) baked products; (22) fruits and fruit juices; (23) sweets; (24) fats and oils; and (25) spices and herbs (Figure 41). Ranked foods based on the score of source and high nutrients under the proposed method are given in Table S88. Food groups were ranked based on the average of scores for free, very low, and low nutrients as follows: (1) fruits and fruit juices; (2) beverages; (3) vegetables and vegetable products; (4) spices and herbs; (5) cereal grains and pasta; (6) breakfast cereals; (7) legumes and legume products; (8) sweets; (9) baby foods; (10) soups, sauces, and gravies; (11) American Indian/Alaska Native foods; (12) nut and seed products; (13) finfish and shellfish products; (14) snacks; (15) dairy and egg products; (16) fats and oils; (17) baked products; (18) lamb, veal, and game products; (19) beef products; (20) poultry products; (21) pork products; (22) meals, entrees, and side dishes; (23) sausages and luncheon meats; (24) fast foods; and (25) restaurant foods (Figure 41). Ranked foods based on the score of free, very low, and low nutrients under the proposed method are given in Table S89.

**Figure 41:**
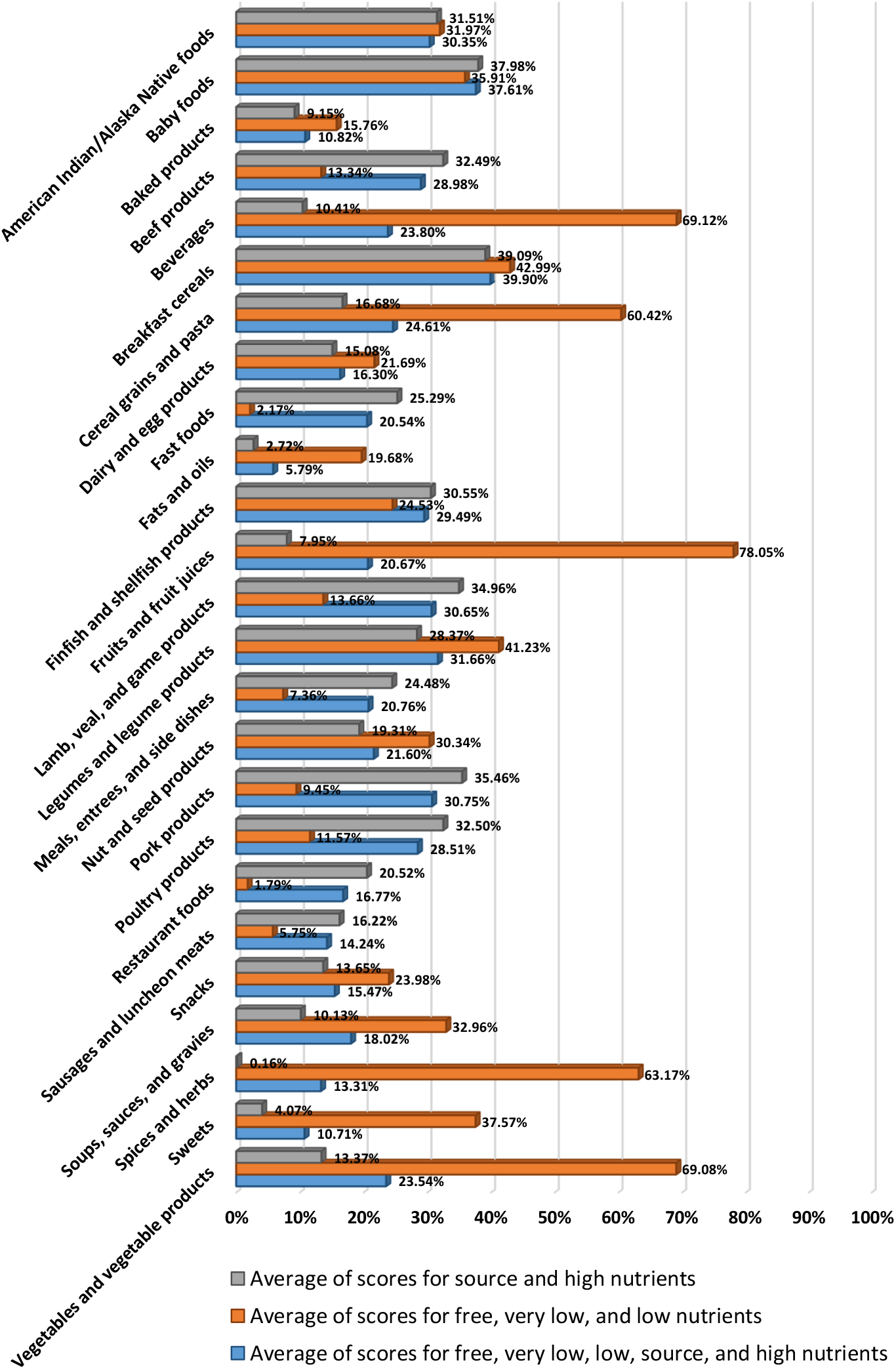
Average of scores for met claims of the nutrient content under the proposed method in food groups.

Food groups were ranked based on the average of scores for free, very low, low, source, and high nutrients as follows: (1) breakfast cereals; (2) baby foods; (3) legumes and legume products; (4) pork products; (5) lamb, veal, and game products; (6) American Indian/Alaska Native foods; (7) finfish and shellfish products; (8) beef products; (9) poultry products; (10) cereal grains and pasta; (11) beverages; (12) vegetables and vegetable products; (13) nut and seed products; (14) meals, entrees, and side dishes; (15) fruits and fruit juices; (16) fast foods; (17) soups, sauces, and gravies; (18) restaurant foods; (19) dairy and egg products; (20) snacks; (21) sausages and luncheon meats; (22) spices and herbs; (23) baked products; (24) sweets; and (25) fats and oils (Figure 41). Ranked foods based on the score of free, very low, low, source, and high nutrients under the proposed method are given in Table S90.

## Data Availability

Publicly available datasets were analyzed in this study. This data can be found here: https://fdc.nal.usda.gov/

## Abbreviations

CAC: Codex Alimentarius Commission
FDA: U.S. Food and Drug Administration
RACC: reference amounts customarily consumed
DV: daily value
NRV: nutrient reference value

## Notes

### Competing Interest Statement

The authors have declared no competing interest.

### Funding Statement

Not funded

